# How Do Nurses Make Clinical Decisions Via Remote Reviews: A Convergent Mixed-Methods Study

**DOI:** 10.64898/2026.07.15.26357946

**Authors:** Yuhan Zhang, Sheera Sutherland, Kathleen Greenway, Louise Stayt

## Abstract

**Background:** Remote clinical reviews have become an integral component of contemporary nursing practice across community and acute care settings. Nurses increasingly make autonomous clinical decisions using telephone, video, and online/digital systems, often with limited sensory information and under conditions of uncertainty. However, empirical understanding of how nurses make clinical decisions via remote reviews remains limited.

**Aim:** To explore and understand how registered nurses (RNs) make clinical decisions about patient care via remote reviews.

**Methods:** A convergent mixed-methods design was employed. Quantitative data (analytic quantitative sample N=53) were collected using validated questionnaires that measured decision-making processes, physician–nurse collaboration, decision-making stress, and perceived decision-making ability. Qualitative data (N=23) were generated through semi-structured interviews. Data collection took place between October 2024 and April 2025. Quantitative data were analysed using descriptive statistics, correlation, and multiple regression. Qualitative data were analysed using framework analysis. Integration was achieved through pillar-building and theory-driven synthesis and illustrated by joint display tables.

**Results:** Most nurses demonstrated a flexible decision-making style, integrating analytical and intuitive reasoning. Both analytical and intuitive processes were positively associated with perceived decision-making ability. Physician– nurse collaboration emerged as a strong predictor of decision-making confidence, while decision-related stress was not a significant predictor. Qualitative findings identified three themes: characteristics of remote review; making adaptive decisions shaped by both internal and external constraints and enablers; and external influencing factors. The integrated findings informed a theory-informed ICE framework to illustrate how nurses make clinical decisions via remote reviews.

**Conclusion:** Remote clinical decision-making is a dynamic cognitive–environmental process rather than a purely individual cognitive act. The ICE framework conceptualises this interaction, extending existing decision-making theories to digitally mediated care.

**Impact:** Understanding remote decision-making supports training design, clinical governance, and the development of Artificial Intelligence-enhanced decision-support tools grounded in ecological bounded rationality.

**What is already known about this topic:**

- Remote clinical review is an increasingly common part of nursing practice in digitally enabled services.
- Nurses undertaking remote reviews often make decisions under uncertainty, using incomplete or indirect clinical information.
- Empirical evidence on how nurses make clinical decisions in remote review contexts remains limited.

**What this paper adds:**

- Most nurses in this study demonstrated a flexible decision-making style, drawing on both analytical and intuitive reasoning rather than one dominant mode.
- Physician–nurse collaboration was positively associated with perceived decision-making ability, while decision-related stress was not a significant predictor in the final models.
- Integrated quantitative and qualitative findings informed a theory-informed Integrated Cognitive–Environmental (ICE) framework for understanding remote nursing decision-making.

**The implications of this paper:**

- Remote clinical decision-making should be supported through service design, training, and digital systems that strengthen collaboration, information synthesis, and cognitive flexibility.
- Artificial Intelligence-enabled or digital decision-support tools for remote care should augment, rather than replace, nurses’ judgment and should be designed around the realities of practice.

## 1. Introduction

Remote modes of clinical care have expanded rapidly worldwide, driven by digital transformation, workforce pressures, and post-pandemic service redesign. Nurses now routinely assess, triage, and manage patients through telephone, video, and digital or online systems across community services and acute virtual wards (NHS England, 2023). These remote models require nurses to make complex clinical judgements while interpreting incomplete or indirect cues, synthesising information from multiple digital sources, and often working without the benefit of physical examination or immediate in-person support (Haimi *et al.,* 2018). Despite the central role of nurses in these services, there remains limited empirical evidence exploring how nurses make clinical decisions in remote contexts and the factors that shape this process.

The rapid growth of remote reviews was further accelerated during the COVID-19 pandemic and has since become a standard part of routine healthcare delivery (Ozanne *et al.,* 2020). Globally, healthcare systems are increasingly implementing virtual wards, hospital-at-home models, and remote monitoring approaches to manage acutely unwell patients outside of hospital settings, aiming to reduce unnecessary admissions and shorter lengths of stay (NHS England & NHS Improvement, 2022). For many patients, especially frail older adults, remote care can enhance access, support ongoing care, and allow treatment in familiar environments, which may improve comfort and functional outcomes (Alderwick & Dixon, 2019; Dehours *et al.,* 2022). However, these advantages bring new clinical and cognitive challenges for nurses responsible for monitoring risk, detecting deterioration, and making timely decisions remotely.

Within remote services, registered nurses (RNs) undertake increasingly autonomous and complex clinical decision-making (CDM), often in settings characterised by reduced medical staffing and high service demand (Health Education England, 2025). CDM is a dynamic, ongoing process that involves collecting, interpreting, and integrating clinical information to determine appropriate action (Tiffen *et al.,* 2014). This process is influenced by internal and external factors, including clinical experience, workload, organisational context, cognitive load, and access to collaborative support (McIntosh *et al.,* 2016; Zhang *et al.,* 2024). Nurses may draw on intuitive pattern recognition, analytical reasoning, or a combination of both, shifting between cognitive modes in response to task demands and situational constraints (Kahneman, 2013; Wouters *et al.,* 2019).

Remote clinical reviews also pose specific risks. The lack of physical assessment increases diagnostic uncertainty and may amplify vulnerability to cognitive bias and error (Dunn Lopez *et al.,* 2017; Haimi *et al.,* 2018). Stress and time pressure may further affect decision quality, especially when nurses need to manage risk independently or escalate concerns across professional boundaries (Johansen & O’Brien, 2016; Park *et al.,* 2023). While theoretical models such as Dual Processing Theory (DPT) and Cognitive Continuum Theory (CCT) propose that reasoning adapts to environmental factors, there is limited nursing-specific research on how these processes work within digitally mediated, hospital-based remote services.

This study addresses this gap by examining nurses’ clinical decision-making during remote reviews, including exploration of experiences, with a focus on decision-making processes, physician–nurse collaboration, stress, and perceived decision-making ability. It is reported in accordance with Mixed Methods Reporting in Rehabilitation & Health Sciences (MMR-RHS) (Tovin & Wormley, 2023). By producing empirically grounded insights into how nurses reason and act in remote settings, this study aims to inform nursing education, clinical practice, workforce development, and the design of safe and supportive digital care systems.

## 2. Aim and objectives

### Aim

To explore and understand how registered nurses make clinical decisions about patient care via remote reviews.

### Objectives

● To explore RNs’ clinical decision-making process via remote reviews
● To explore potential internal and external influencers on RNs’ clinical decision-making via remote reviews
● To explore RNs’ experiences and perceptions of clinical decision-making via remote reviews
● To explore associations and correlations between the internal and external influencers on RNs’ clinical decision-making (such as the demographics, attitudes towards physician-nurse collaboration, and decision-making associated stress) and RNs’ perceived decision-making ability.

## 3. Hypothetical statistical model

The following diagram (***Fig.*** 1) serves as the hypothetical statistical framework for this study. The hypothetical statistical model was informed by our prior systematic review of remote clinical decision-making in nursing (Zhang *et al.,* 2024), which identified recurring associations between reasoning processes, collaboration, stress, and decision-making capability. Decision-making denotes nurses’ cognitive or information-processing processes when making clinical decisions. The physician-nurse collaboration refers to nurses’ attitudes towards collaboration between doctors and nurses. Decision-making stress refers to the stress nurses experience when making autonomous decisions in remote reviews. Lastly, decision-making ability refers to nurses’ perceived ability to make clinical decisions. There are three independent variables (IVs) as illustrated on the left side of ***Figure*** 1, and one dependent variable (DV) as the outcome.

**Figure 1.**
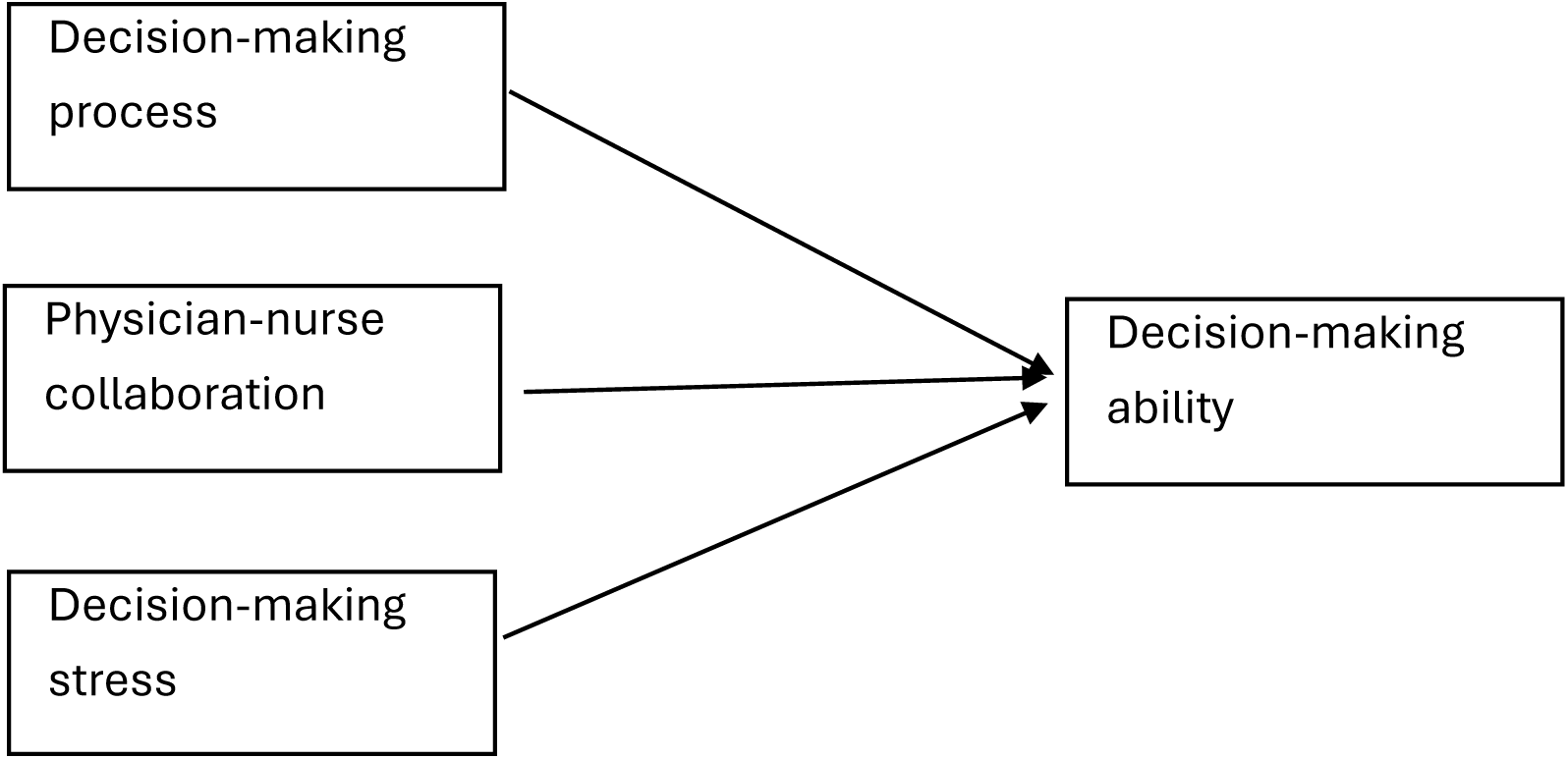
The hypothetical relationship of the variables.

While the hypothetical model (***Figure*** 1) specifies testable relationships among key variables, the broader theoretical framework (***Figure*** 2) situates these relationships within an epistemic/theoretical framing. Previous clinical decision-making models, e.g., dual-process theory, suggest that clinicians use both intuitive and analytical thinking in their decision-making. In addition, there are various internal and external influences, such as patient factors (medical and non-medical factors), clinicians’ internal system (age, gender, past experience, previous education and training), and environmental factors (e.g., workload, the use of clinical decision support system (CDSS)) and technical issues (Zhang *et al.,* 2024). To map these to the chosen ontological and epistemological approach, the theoretical framework of the study is presented in ***Figure*** 2. The study was informed by an existing theoretical framework of remote clinical decision-making, which was used to shape both the design and the analysis. The framework identified key influences on decision-making, including cognitive processes, clinical experience, contextual pressures, collaboration, and features of the remote review environment. These domains informed the selection of survey measures, the development of interview questions, and the integration of quantitative and qualitative findings.

**Figure 2.**
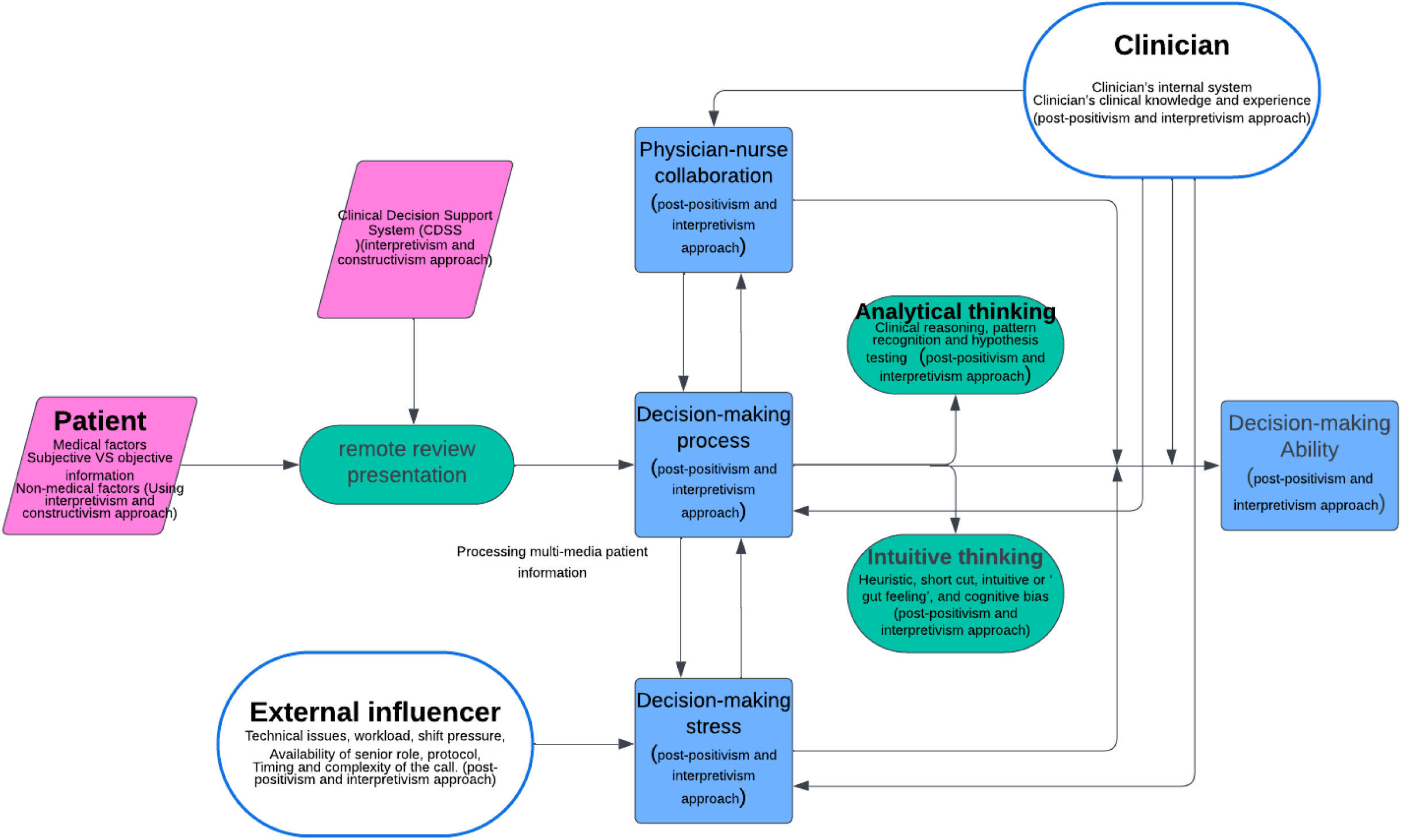
The Theoretical framework of the stud.

## 4. Methods

### 4.1 Design

This study employed a convergent mixed-methods design (Creswell & Plano Clark, 2011) (***Figure*** 3), integrating quantitative and qualitative data to provide a comprehensive understanding of nurses’ remote clinical decision-making. The design combined a descriptive cross-sectional quantitative component with a qualitative interview component. The quantitative component examined nurses’ perceived clinical decision-making ability in remote review contexts, potential internal and external influences on decision-making, and associations between these factors. Quantitative data were collected using validated scales and demographic items. A qualitative component, using semi-structured interviews, was undertaken to explore nurses’ experiences and perceptions of clinical decision-making via remote reviews in greater depth. Integrating the two strands enabled both the measurement of key relationships and a richer understanding of how decision-making was experienced in practice. The design included collecting and analysing numerical and narrative data, followed by integration through triangulation and synthesis.

**Figure 3.**
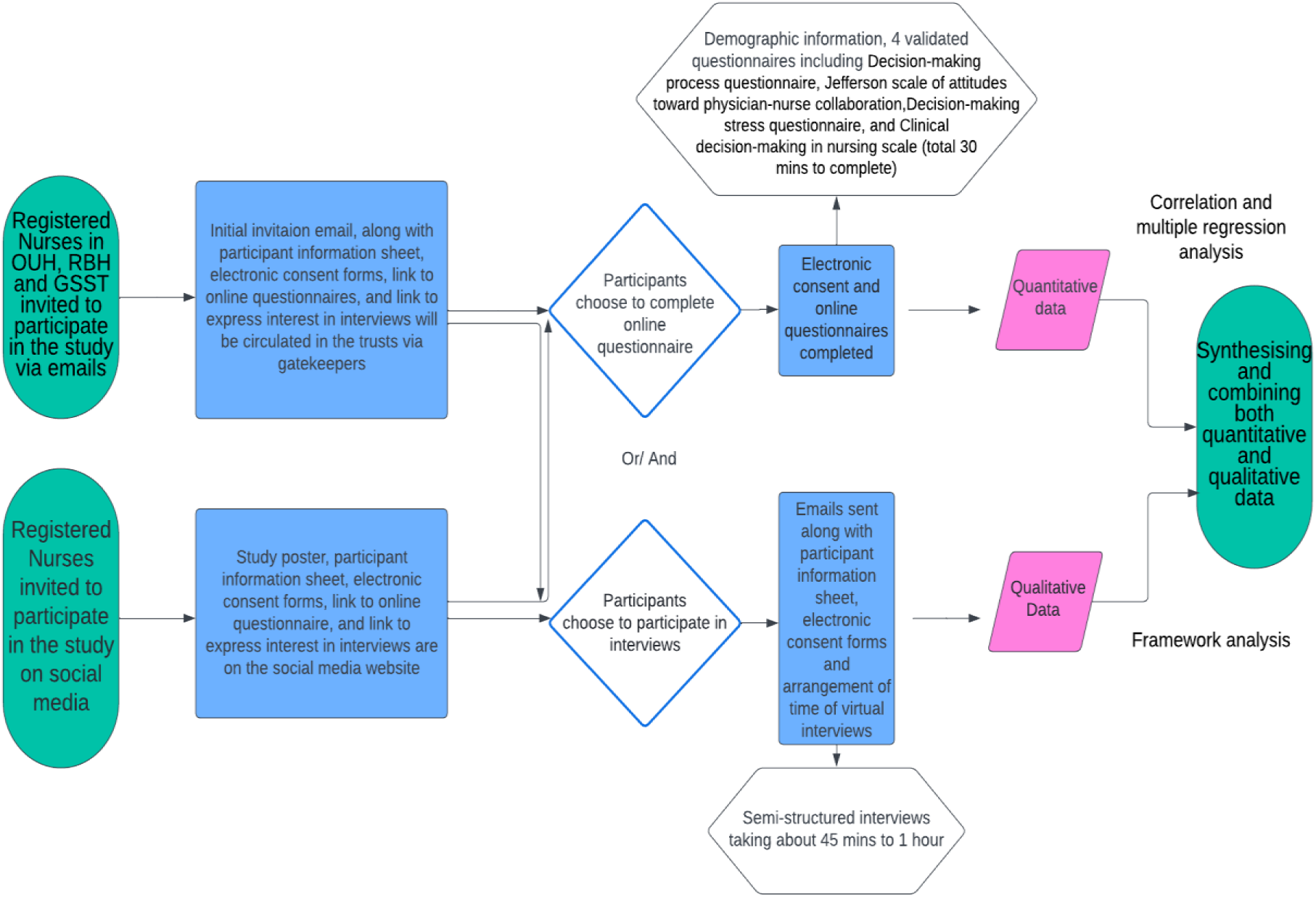
The Study Flow Chart.

### 4.2 The Study Setting

The study sites were three hospitals in three regions of the UK. In addition to recruiting registered nurses directly through the three NHS Trusts, recruitment was supplemented by advertising using a range of social media platforms (Facebook, X, and LinkedIn).

### 4.3 SAMPLE AND RECRUITMENT

#### 4.3.1 Inclusion and Exclusion Criteria

The study population was RNs, who conduct remote reviews at least once a month in the UK.

Inclusion criteria were as follows:

– Registered nurses with the Nursing and Midwifery Council (NMC), including clinic/outpatient nurses, specialist nurses, specialist nurse practitioners, advanced nurse practitioners, and advanced clinical practitioners who are registered nurses, in acute hospitals or primary settings
– RNs who are actively conducting remote reviews on patients (conducting at least one remote review per month), either via video, telephone, or other online platforms
– RNs who conduct remote reviews in outpatient clinics or inpatient settings (such as virtual wards) for initial consultations or follow-up reviews.
– RNs who work in the virtual ward at the hospital where the author works and are colleagues of the researcher will be excluded from participating in the interviews. These colleagues, however, were able to complete the anonymous online questionnaire.

The study excluded student nurses and registered nurse associates.

#### 4.3.2 Sampling

A mixed-method approach, involving two different sampling techniques, was used. Convenience sampling was employed for the quantitative component, while purposive sampling was utilised for the qualitative approach. RNs with varied years of experience, age, gender, ethnicity, education level, and speciality were selected for interviews to gain a broad spectrum of experiences and opinions.

#### 4.3.3 Recruitment

Registered nurses undertaking remote reviews were recruited through three NHS hospital trusts in England and through professional social media platforms. Within the hospital trusts, study invitations were distributed by local gatekeepers, including Advanced Clinical Practitioner leads and Divisional Nurse Leads, to relevant nursing staff.

The researcher is a registered nurse with experience in remote clinical practice and doctoral training in mixed-methods research; to minimise role conflict, nurses working directly with the researcher in the local virtual ward were excluded from interview participation.

### 4.4 Data collection

Data collection comprised a quantitative questionnaire and qualitative semi-structured interviews (see ***Appendix*** 1). Quantitative data were collected using an anonymous online questionnaire hosted on Qualtrics^XM^ software (Version 2024, Copyright © 2024, Qualtrics, Provo, UT). The questionnaire included demographic items and validated measures relating to clinical decision-making, physician–nurse collaboration, and decision-making-associated stress. No identifiable personal information or URL data were collected. Before accessing the questionnaire, potential participants were asked to confirm that they had read the Participant Information Sheet, met the eligibility criteria, and provided informed consent electronically. Participants who undertook both face-to-face and remote reviews were reminded to answer in relation to remote reviews only.

Qualitative data were collected through semi-structured interviews. Nurses who wished to take part in an interview could indicate their interest either at the end of the questionnaire or via a separate online expression-of-interest form. Interested participants provided their contact details and preferred communication method, after which the researcher contacted them directly to arrange an interview at a mutually convenient time. Interviews were conducted by video call. At the start of each interview, participants were asked to confirm that they had read and understood the Participant Information Sheet, met the eligibility criteria, and consented to participate. The interviews explored nurses’ experiences of clinical decision-making during remote reviews in greater depth, using reflection on clinical scenarios from practice.

#### 4.4.1 Data Collection Instruments

Validated measurement tools were selected to measure the key variables identified for this doctoral study.

##### 4.4.1.1 Decision-Making Ability

Decision making ability was measured by Clinical Decision-Making in Nursing Scale (CDMNS) (Jenkins, 1985), which is a 40-item questionnaire having four subscales (Search for alternatives or options; Canvassing of objectives and values; Evaluation and re-evaluation of consequences; Search for information and Unbiased assimilation of new information) with five-point Likert Scale ranging from “strongly agree” (=5) to “strongly disagree” (=1). Pooled alpha across imputations indicated low reliability for most subscales: α = 0.293 for the CDMNS_option subscale, α = 0.527 for the CDMNS_objectives subscale, α = 0.539 for the CDMNS_evaluation subscale, and α = 0.112 for the CDMNS_assimilation subscale.

##### 4.4.1.2 Decision-Making Process

The decision-making process (DMP) was measured by a 5-point Likert scale of 24 items (Lauri & Salanterä, 2002; Bjørk & Hamilton, 2011; Abate *et al.,* 2022). The instrument has five response options: ‘never’, ‘rarely’, ‘sometimes’, ‘often’, and ‘always’. This instrument was developed by incorporating analytical information processing and intuitive decision-making, and is based on several theories, including Hammond’s Cognitive Continuum Theory (1996). Pooled Cronbach’s alpha across imputations indicated acceptable reliability: α = 0.762 for the analytical subscale, α = 0.779 for the intuitive subscale, and good reliability of α = 0.87 for the DMP total scale.

##### 4.4.1.3 Jefferson Scale of Attitudes Toward Physician-Nurse Collaboration (JSAPNC)

JSAPNC (Hojat & Herman, 1985; Hojat *et al.,* 1997 & 1999) is a validated tool to measure physician-nurse collaboration (PNC). The questionnaire consists of 15 items measuring four factors, including shared education and teamwork, caring as opposed to curing, nurses’ autonomy, and physicians’ authority (Hojat *et al.,* 1999). The participants need to choose a scale from ‘strongly disagree’ (=1) to ‘strongly agree’ (=4). Pooled Cronbach’s alpha across imputations indicated acceptable reliability: α = 0.718 for the shared education and collaboration subscale, α = 0.420 for the nurses’ autonomy, α = 0.311 for the caring vs curing subscale, α = 0.774 for the physicians’ authority subscale, and α = 0.812 for the PNC_total scale.

##### 4.4.1.4 Decision-making Stress

Decision-making Stress (Stress) was measured by a 5-item questionnaire (Appendix 7) developed by Bucknall and Thomas (1996). These questions were developed by investigating nurses’ reflections in critical care settings (Bucknall & Thomas, 1996) and have been used to measure decision-making stress (Park *et al.,* 2023). Pooled Cronbach’s alpha across imputations indicated acceptable reliability: α = 0.851 for the stress scale.

As previously discussed, the current literature indicates that the stress of facing uncertainty and the fear of making mistakes can affect nurses’ decision-making. Therefore, two more questions (***Appendix*** 2) were adapted from Nevalainen et al. (2012) to investigate tolerance of uncertainty and fears of making mistakes (the words ‘diagnosis’ and ‘medical’ were removed from the question items to accommodate nurses’ decision-making).

#### 4.4.2 Qualitative method

Generic qualitative research method was employed to be compatible with mixed-method study design. An interview topic guide was developed to inform interviews. The topic guide and the theoretical model (***Figure*** 2) were developed by the systematic review (Zhang *et al.,* 2024), stakeholder discussion, and interactions with patient and public representatives. Interviews took place via Zoom video calls, each lasting up to one hour. The interviews were recorded after obtaining the participant’s agreement throughout the entire interview session. The audio transcripts were downloaded after the interview. Only the audio output from a video call was recorded and transcribed. Reflective notes were taken immediately after the interview and included in the data analysis.

### 4.5 Data analysis

#### 4.5.1 Quantitative data analysis

Data were analysed using SPSS (IBM Version 29.0.1.1). Descriptive statistics (means, standard deviations, medians and interquartile ranges, frequencies, and percentages) were calculated for demographic variables and main study response measures (CDMNS, PNC, DMP, and Stress). Internal consistency of the CDMNS, DMP, PNC and stress scales was assessed using Cronbach’s alpha.

Associations between decision-making model scores and demographic variables were tested using both Pearson’s and the Spearman test. Pearson correlations were used when variables met parametric assumptions. Spearman correlations were used for ordinal scales, non-normally distributed variables, or when assumptions for Pearson correlation were violated. Group comparisons across categorical demographics (such as gender, nursing band, qualification) were performed via independent *t*-tests and one-way ANOVA, with post hoc tests if required.

Missing data were evaluated before the main analyses. The percentage of missing values across variables ranged from 0% to 13.2%, with an overall missingness rate of 4.48%. Little’s MCAR test was non-significant (χ² (35) = 1.317, p = 1.000), suggesting that missingness was consistent with data missing completely at random. Given that missingness was not trivial across several variables, multiple imputation was used rather than listwise deletion. Twenty imputed datasets were created, consistent with recommendations that the number of imputations should reflect the proportion of incomplete cases. Each dataset was analysed separately, and pooled estimates were calculated using Rubin’s rules.

#### 4.5.2 Framework analysis as the qualitative data analysis

Steps of the framework analysis were followed: 1. Transcription of the interview verbatim (word for word); 2. Familiarisation with the interview; 3. Coding (labelling); 4. Developing a working analytical framework; 5. Applying the analytical framework; 6. Charting data into the framework matrix, and finally, 7. Interpreting the data (Gale *et al.,* 2013). The participants’ names have been de-identified and pseudo-named.

### 4.6 Integration of Data

The integration of quantitative and qualitative findings was conducted using a combined pillar-building and theory-building approach, designed to produce both empirical and conceptual syntheses. This dual strategy followed established mixed-methods integration guidance (Fetters & Freshwater, 2015; Johnson *et al.,* 2017), ensuring that findings were not only merged at the data level but also interpreted through relevant theoretical frameworks of clinical decision-making.

The findings were then integrated with previous theories to produce both empirical and conceptual syntheses (Fetters & Freshwater, 2015; Johnson *et al.,* 2017). The Pillar Integration Process (PIP) was employed to integrate quantitative and qualitative findings into higher-order meta-inferences, referred to as ‘pillars’. Stages of PIP, including listing (listing raw data from QUAN and QUAL), matching (aligning similar data from QUAN and QUAL), checking (cross-checking completeness), and pillar building (Comparing and contrasting the findings to build inferences), were followed. Quantitative results (e.g., correlations between decision-making process, decision-making ability, age, qualification, experience, stress, and collaboration) and qualitative themes (e.g., making adaptive decisions, external influencing factors) were displayed side by side in an integration matrix. The open comment section in the survey was incorporated into the QUAL section of the display for illustration and integration, as it provides further explanation or comments on the quantitative data (these are highlighted in the survey comments on the right side of the table). Findings were compared for convergence, divergence, and complementarity.

### 4.7 Ethical Considerations

Ethical approval was obtained from the local university ethics committee (Sponsorship review reference: HLS.NHSS.24.4), University Research Ethics Committee (UREC) and the Health Research Authority (HRA) (proportionate review) (IRAS ID: 335816) for the study protocol, electronic consent forms, participant information sheet, and poster. Approval from the local Research and Development (R&D) department of each participating NHS Hospital Trust was sought prior to recruitment.

### 4.8 Methodological rigour

Strategies to enhance methodological quality were considered across both study strands. In the quantitative component, rigour was supported through the use of validated instruments, standardised data collection procedures, and transparent statistical analysis. In the qualitative component, trustworthiness was enhanced through reflexive interviewing, systematic analysis, and attention to credibility, dependability, and confirmability (Lincoln & Guba, 1985; Plano Clark & Ivankova, 2016).

## 5. Results

A total of 81 registered nurses participated in the quantitative phase by completing the questionnaire, and 23 RNs took part in the qualitative interviews. Findings from the quantitative and qualitative strands are presented in turn below.

### 5.1 Quantitative Findings

#### 5.1.1 Demographics

From the 81 participants who completed the questionnaire, ages ranged from 26 to 70 years (M = 44.83, SD = 10.15), with the distribution appearing approximately normal (Shapiro–Wilk p = 0.487, Skewness = 0.20). Most participants were female (n = 70, 86.4%), and 11 (13.6%) were male nurses. The majority of the participants identified as White British (57.0%), followed by Asian (22.8%). Years of nursing experience ranged from 3 to 47 years, with a median of 15.5 years (IQR 10.0–25.0). More than half of participants were employed at UK Agenda for Change Band 6 or 7 level (64.5%), broadly corresponding to experienced specialist nurses, senior nurses, or advanced practice roles. Among them, nearly half (45.6%) had master’s degrees. Most participants were clinical specialist nurses (31.6%) or advanced nurse/clinical practitioners (26.6%), and they conducted remote reviews on a daily (50.6%) or weekly (38.0%) basis. Participants came from almost all specialities in healthcare settings, including primary care, ambulatory/emergency, cardiology, neurology, and virtual wards (including hospital-at-home). The sample comprised nurses with substantial clinical experience and many were working in senior or specialist roles, suggesting that participants were likely to have a high level of autonomy and responsibility in remote clinical decision-making. Detailed demographic information is in ***Table*** 4.

**Table 4.**
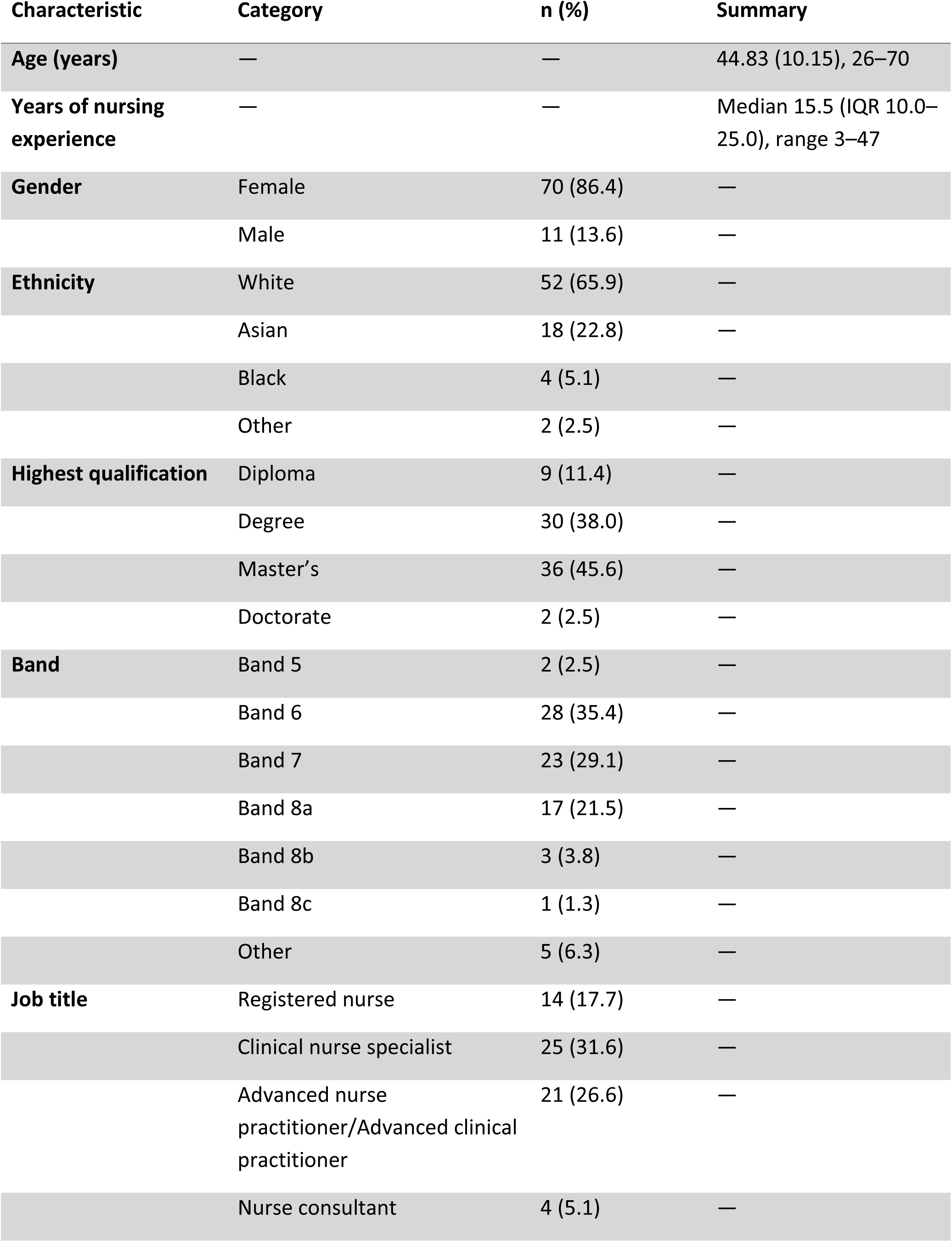
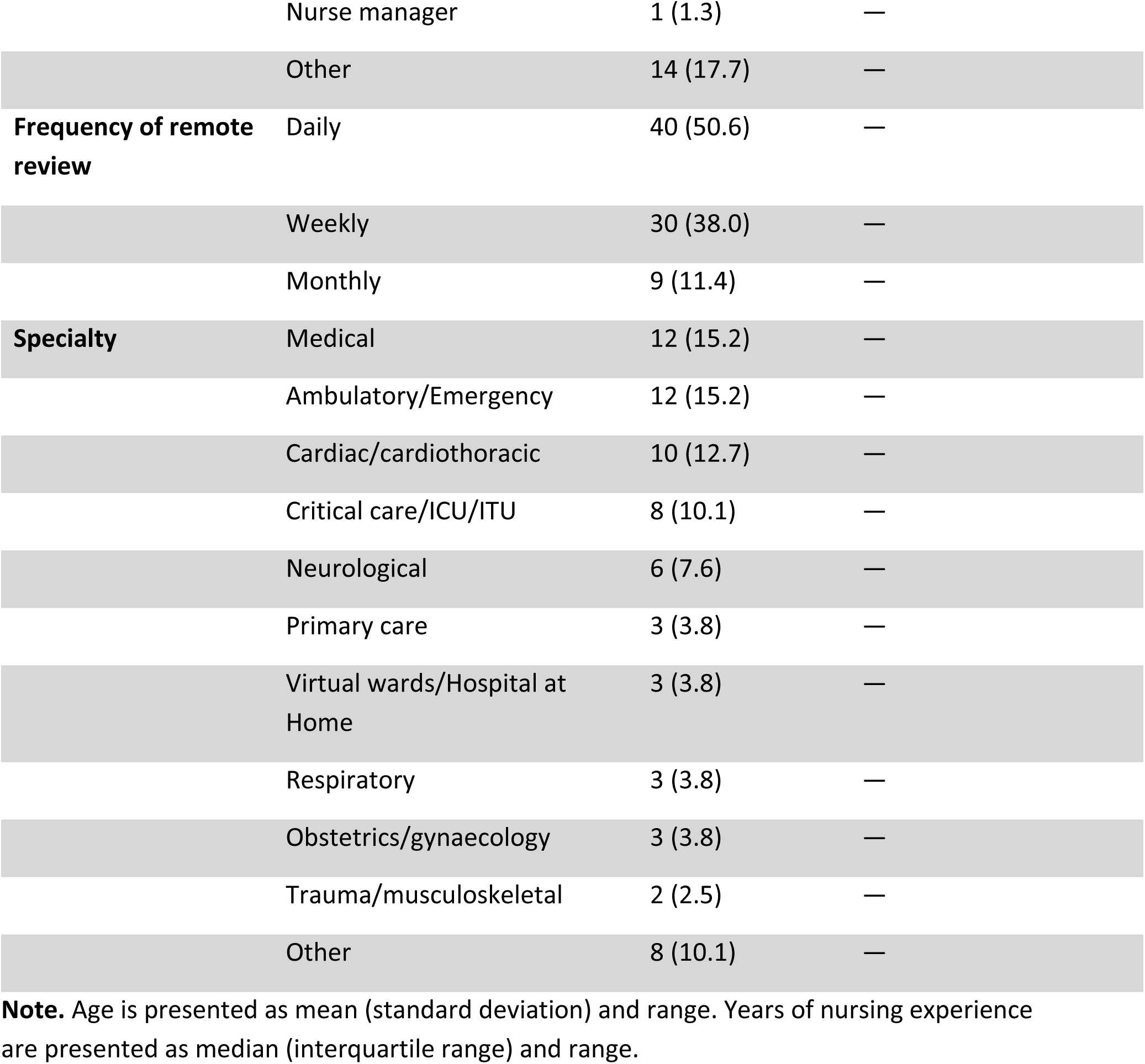
Participant demographic and professional characteristics (n = 81)

#### 5.1.2 Descriptive statistics for the main study variables

Overall, 81 nurses completed the survey. Participant characteristics are reported for the full sample (n = 81). For analyses involving composite scale scores, the sample reduced to 53 because only these participants had sufficient item completion to allow reliable scoring of the relevant measures. Remaining item-level missingness within this analytic sample was handled using multiple imputation.

Descriptive statistics for the main study variables are shown in ***Table*** 5. Participants demonstrated generally high levels of perceived clinical decision-making ability, moderate-to-high physician–nurse collaboration, and moderate stress. Most nurses were classified as flexible decision-makers (73.6%), with fewer classified as analytical (22.6%) or intuitive (3.8%).

**Table 5.** Descriptive statistics and reliability for study variables.

| *Variable / scale* | *n* | *Observed*<br>*range* | *Mean*<br>*(SD)* | *Skewness* | *Kurtosis* | *Cronbach's*<br>*$\alpha$* |
| --- | --- | --- | --- | --- | --- | --- |
| ***Decision-Making***<br>***Process (DMP)*** |  |  |  |  |  |  |
| *DMP total* | 53 | 64–80 | 70.25<br>(3.55) | 0.55 | 0.22 | 0.870 |
| *DMP analytical* | 53 | 39–59 | 48.96<br>(5.13) | 0.05 | -0.72 | 0.779 |
| *DMP intuitive* | 53 | 34–58 | 47.21<br>(5.12) | -0.31 | -0.33 | 0.762 |
| ***Clinical Decision-***<br>***Making in Nursing***<br>***Scale (CDMNS)*** |  |  |  |  |  |  |
| *CDMNS total* | 53 | 123–167 | 148.12<br>(11.42) | -0.42 | -0.59 | 0.758 |
| *CDMNS options* | 53 | 31–44 | 37.00<br>(3.12) | 0.02 | -0.36 | 0.293 |
| *CDMNS objectives* | 53 | 25–44 | 37.15<br>(4.25) | -0.48 | -0.04 | 0.527 |
| *CDMNS evaluation* | 53 | 27–45 | 37.76<br>(4.24) | -0.19 | -0.46 | 0.539 |
| *CDMNS assimilation* | 53 | 31–41 | 36.18<br>(2.73) | 0.09 | -0.74 | 0.112 |
| ***Physician–Nurse***<br>***Collaboration (PNC)*** |  |  |  |  |  |  |
| *PNC total* | 53 | 40–60 | 55.21<br>(4.17) | — | — | 0.812 |
| *Shared education and teamwork* | 53 | 19–28 | 26.29<br>(1.97) | — | — | 0.718 |
| *Caring versus curing* | 53 | 7–12 | 10.97<br>(1.14) | — | — | 0.311 |
| *Nurse autonomy* | 53 | 9–12 | 11.52<br>(0.68) | — | — | 0.420 |
| *Physician authority* | 53 | 3–8 | 5.58<br>(1.56) | — | — | 0.774 |
| ***Stress*** |  |  |  |  |  |  |
| *Total stress score* | 53 | 7–30 | 18.32<br>(6.04) | 0.19 | -0.65 | 0.851 |

**Note:** Values for SD, skewness, kurtosis, and Cronbach's alpha are pooled across 20 imputed datasets where applicable. Dashes indicate values not reported or not applicable. For PNC, skewness was approximately -1.4 to -1.7 for most subscales, and kurtosis was approximately 3–4.

#### 5.1.3 Decision-making process scale

Nurses were classified according to their dominant decision-making model based on their total score (out of a range of 24 to 96). Nurses were categorised as analytical (<67), flexible (68–78), or intuitive (>78) decision-makers. The majority of nurses demonstrated a flexible decision-making style (73.6%), followed by an analytical style (22.6%) and an intuitive style (3.8%).

Analytical decision-making was positively correlated with years of experience (Pearson r = 0.324, p = 0.023). There was no difference observed between female nurses (M = 70.30, SD = 3.653) and male nurses (M = 69.8, SD = 2.950) in DMP analytical (t(51) = 0.658, p = 0.511), Intuitive (t(51) = 0.452, p = 0.651) or total score (t(51) =-0.296, p = 0.767). One-way ANOVA showed there was a difference in analytical mean scores between the different job bands, and further post hoc tests showed differences between Band 7 and Band 6 (mean difference=4.799, p = 0.044) and between Band 7 and Band 8a (mean difference=5.582, p = 0.039). Multiple regression indicated that years of experience were a significant predictor of analytical decision-making (R² = 0.15, p = 0.024). No demographic variable significantly predicted intuitive decision-making or total score in the regression model.

#### 5.1.4 CDMNS

Pooled descriptive statistics across 20 imputed datasets indicated that the mean total CDMNS score was 148.12 (SD = 11.421) (out of a maximum 200), with scores ranging from 123 to 167. The pooled mean CDMNS score represented approximately three-quarters of the maximum possible score, indicating generally well-developed clinical decision-making ability within the sample. The pooled skewness (–0.416) and kurtosis (–0.591) indicated an approximately normal distribution. Similar patterns were observed across the four subscales (options, objectives, evaluation, and assimilation), with means ranging from 36.16 to 37.76 and standard deviations between 2.729 and 4.252.

Spearman’s rho correlations were computed across 20 imputed datasets to examine associations between demographic variables, collaboration, stress, decision-making processes, and nurses’ clinical decision-making ability. Age was positively correlated with the CDMNS_evaluation subscale (ρ = 0.347, p = 0.011; moderate correlation), while years of experience correlated with both CDMNS_options (ρ = 0.334, p = 0.041; moderate correlation) and CDMNS_evaluation (ρ = 0.317, p = 0.021; moderate correlation). Qualification level showed a significant association with CDMNS_objectives (ρ = 0.311, p = 0.024; moderate correlation), although this should be interpreted cautiously given the modest internal consistency of some subscales.

The total physician–nurse collaboration score (PNC_total) was positively related to CDMNS_objectives (ρ = 0.392, p = 0.004; moderate correlation) and assimilation (ρ = 0.295, p = 0.032; weak correlation). PNC_total was not significantly related to CDMNS_options (ρ = 0.115, p = 0.174), and CDMNS_evaluation (ρ = 0.081, p = 0.686). Similarly, the PNC_shared education subscale correlated positively with CDMNS_objectives (ρ = 0.343, p = 0.012) and CDMNS_total (ρ = 0.285, p = 0.039), whereas physician authority demonstrated negative correlations with CDMNS_objectives (ρ = –0.284, p = 0.039) and assimilation (ρ = –0.290, p = 0.035).

Both analytical and intuitive decision-making styles (DMP) were significantly associated with overall clinical decision-making ability. Analytical style was strongly correlated with CDMNS_total (ρ = 0.442, p = 0.001) and multiple subscales (ρ = 0.309–0.388, p < 0.05), while intuitive style was also moderately related to CDMNS_total (ρ = 0.384, p = 0.005). No significant correlations were observed between stress levels and decision-making scores.

Overall, the results suggest that both analytical and intuitive reasoning styles contribute to nurses’ clinical decision-making and that higher collaboration scores may reinforce confidence in the decision-making ability.

#### 5.1.5 PNC

The PNC Scale demonstrated moderate-to-high levels of perceived collaboration among participants (M = 55.21, SD = 4.17). Subscale analysis showed particularly high scores for Shared Education and Teamwork (M = 26.29, SD = 1.97) and Nurse Autonomy (M = 11.52, SD = 0.68). The lowest subscale mean was observed in Physician Authority (M = 5.58, SD = 1.56). Skewness values (–1.3 to –1.7) indicate that the majority of participants reported high collaboration. Missing data were minimal (FMI < 0.15), and pooled estimates from multiple imputation demonstrated high relative efficiency (≥0.99).

Qualification was significantly and positively correlated with total PNC (ρ = 0.307, p = 0.028) and the shared education subscale (ρ = 0.338, p = 0.014). Additionally, qualification was negatively correlated with PNC_physician authority (ρ = −0.266, p = 0.047). Job banding also showed a significant positive correlation with *shared education* (ρ= 0.316, p = 0.026). Age was significantly correlated with nurses’ autonomy (ρ= 0.296, p = 0.044). No significant correlations were found between stress and any of the PNC dimensions, or between years of experience and PNC measures.

#### 5.1.6 Stress and tolerance of uncertainty

The total stress scores (n = 53) ranged from 7 to 30 (out of a possible 30), with a mean of 18.32 (SD = 6.04).

Regarding the tolerance of uncertainty, 14 (29.8%) participants reported difficulties tolerating uncertainty in decision-making, 27 (57.4%) reported tolerating it quite well, and 6 (12.8%) reported tolerating it very well.

Correlation analyses were conducted to examine the relationships between total stress scores and demographic variables. Stress scores showed weak correlations with gender (ρ = 0.166), age (ρ = −0.165), qualification (ρ = −0.142), years of experience (ρ = −0.119), and job band (ρ = −0.232). None of these correlations reached statistical significance (all p > 0.05), indicating no significant association between stress levels and the demographic variables examined.

The level of tolerance of uncertainty is noted to be associated with PNC_total (ρ = 0.378, p = 0.009).

#### 5.1.7 Regression models predicting perceived clinical decision-making ability

Initial inspection of Pearson correlations suggested a high positive correlation between the variables of DMP_intuitive and DMP_analytical (Pearson r > 0.7). Two separate models were tested: one including DMP_analytical alongside PNC_total and stress, and one including DMP_intuitive with the same predictors.

##### 5.1.7.1 Model 1: Analytical reasoning

Assumptions of linearity, independence, normality, homoscedasticity, outliers, and multicollinearity were checked and met. Regression analyses were conducted across 20 multiply imputed datasets. Pooled regression coefficients, standard errors, t-values, and p-values are reported (See ***Table*** 6). The mean R² across imputations was 0.314 with a mean adjusted R² of 0.311, indicating that the model explained approximately 31% of the variance in the outcome. Across the 20 imputations, the regression model was statistically significant, with F-values ranging from 6.37 to 10.46 (mean F = 7.40, SD = 0.92). For reference, the model F-test in the original dataset was F (3, 49) = 10.455, p < 0.001; however, all inferential conclusions are based on the pooled estimates from the imputed datasets. In this model, perceived clinical decision-making ability was positively associated with physician–nurse collaboration and analytical decision-making, while stress was not a statistically significant independent predictor. For each one-unit increase in physician–nurse collaboration, CDMNS_total increases by approximately 1.08 points. Greater use of analytical reasoning is associated with higher perceived decision-making ability.

**Table 6.** Pooled regression coefficients, standard errors, t-values, p-values, and 95% confidence intervals for the analytical and intuitive regression models.

| Predictor | B | Se | T | p | 95%CI |
| --- | --- | --- | --- | --- | --- |
| **Analytical Regression Model** |  |  |  |  |  |
| (Constant) | 55.139 | 22.215 | 2.482 | 0.013 | [11.595, 98.683] |
| PNC_total | 1.080 | 0.321 | 3.369 | <0.001 | [0.452,1.708] |
| Stress | -0.340 | 0.226 | -1.500 | 0.134 | [-0.785,0.106] |
| DMP_analytical | 0.809 | 0.257 | 3.143 | 0.002 | [0.305,1.314] |
| **Intuitive Regression Model** |  |  |  |  |  |
| (Constant) | 68.723 | 22.130 | 3.105 | 0.002 | [25.346, 112.100] |
| PNC_total | 0.991 | 0.335 | 2.955 | 0.003 | [0.334,1.648] |
| Stress | -0.282 | 0.236 | -1.196 | 0.233 | [-0.746,0.182] |
| DMP_intuitive | 0.633 | 0.271 | 2.240 | 0.019 | [0.103,1.164] |

##### 5.1.7.2 Model 2: Intuitive reasoning

Pooled regression coefficients, standard errors, t-values, and p-values are reported (See ***Table*** 6). The mean adjusted R² across imputations was 0.26, explaining approximately 26% of the variance in the outcome. Across the 20 imputations, the regression model was statistically significant, with F-values ranging from 5.324 to 8.450 (mean F = 6.40, SD = 0.74). The model F-test in the original dataset was F(3, 43) = 8.450, p < 0.001.

### 5.2 Qualitative Findings

Twenty-three nurses with diverse backgrounds participated in the interview, and their demographics are outlined in ***Appendix*** 3. Three overarching themes were identified: (1) ***characteristics of remote reviews;*** (2) ***making adaptive decisions shaped by both internal and external constraints and enablers;*** and (3) ***external influencing factors***.

#### 5.2.1 Theme 1: Characteristics of remote review

Participants portrayed remote review as more than a technical adjustment to care delivery; rather, it represented a fundamental reconfiguration of clinical reasoning, communication, and relational practice. Two interrelated subthemes were identified: (1) remote review as a hybrid invention driven by practical need, and (2) the perceived benefits and challenges of conducting assessments at a distance.

##### 5.2.1.1 Remote Review as a Hybrid Invention

Participants consistently described remote review as an integrated and adaptive component of contemporary nursing practice rather than a replacement for face-to-face care. Remote and in-person encounters were blended within hybrid care pathways, shaped by patient need, clinical risk, and service pressures. Monica (Fracture Prevention Specialist Nurse) explained that this blended approach had become routine: *“A lot of services have moved to a blend—even if it starts face-to-face, we’ll follow up on the phone.”*

Remote reviews were used flexibly for triage, follow-up, monitoring, reassurance, and early intervention. Echo (IBD Specialist Nurse) described the role of remote advice lines in maintaining continuity and preventing unnecessary admissions: *“One of the major parts of this job is running an advice line… when they’re having a relapse, we try to keep them out of A&E.”* These accounts highlighted how remote review was embedded within broader care pathways rather than operating as a standalone activity.

Participants described using multiple modalities, including telephone, video, secure email, and review of the electronic patient record. Telephone consultations remained the most common due to reliability and patient familiarity. Monica noted, *“Very few people phone in to ask for a video call… we do have them, but it’s rare.”* However, video review was valued where visual assessment or relational presence enhanced safety. Lynn (Parkinson’s Nurse Specialist) explained, *“With video, I can check tremor, facial expression, even get them to walk up the hallway—it adds a safety element.”* These accounts illustrated how modality choice was situational and clinically driven.

Hybrid practice also extended to team-based care. Kate (Heart Failure Nurse Consultant) described how remote review supported multidisciplinary working in hospital-at-home services: *“We’ve developed the MDT [multidisciplinary team] further—there’s more remote review now because of hospital-at-home pathways.”*

##### 5.2.1.2 Benefits and Challenges of Remote Review

Participants highlighted several benefits of remote review, particularly improved accessibility and patient convenience. Nurses described patients as more relaxed when consulting from home. Rose (Hypertension Nurse Specialist) reflected, *“You can ring them at home while they’re having a cup of tea—they seem more relaxed because they’re in their own environment.”* Remote review was also viewed as supporting efficiency and structured working practices. Emma explained, *“I type everything as I speak, and the letter goes out the same day—I don’t tend to run behind.”*

Despite these benefits, participants described significant challenges. A central concern was the loss of non-verbal cues and physical examination, which increased uncertainty and cognitive effort. Faith (Stroke Nurse Specialist) noted, *“You’ve just not got those non-verbal clues… you can’t see their body language or responses.”* Similarly, Iris (Respiratory Nurse Specialist) described sensory deprivation during assessment: *“If they say their sputum’s green, you can’t see if they’re pale or breathless.”*

Participants also reflected on heightened accountability when making autonomous decisions remotely. One nurse described the weight of responsibility: *“You’re making decisions on your own a lot more, and that responsibility feels heavier when you haven’t physically seen the patient.”* (Dawn, Clinical Nurse Specialist).

Overall, participants portrayed remote review as a multifaceted and situational practice. While it enhanced accessibility, efficiency, and continuity, it also introduced new forms of uncertainty, responsibility, and relational complexity.

#### 5.2.2 Theme 2: Making adaptive decisions shaped by both internal and external constraints and enablers

Participants described decision-making as a dynamic and iterative cognitive process that integrates analytical reasoning, intuitive judgement, and contextual awareness. Remote decision-making was characterised by continuous cycles of information gathering, hypothesis testing, risk appraisal, and reflection, shaped by professional experience and system constraints. Nurses demonstrated bounded yet adaptive rationality, making defensible, patient-centred decisions despite uncertainty and limited sensory information. Three interrelated subthemes were identified: (1) decision-making process, (2) decision-making ability, and (3) knowledge and experience.

##### 5.2.2.1 Decision-Making Process

Participants consistently described decision-making as beginning with “building up the picture”—constructing a holistic mental representation of the patient in the absence of physical presence. This involved synthesising electronic health records, laboratory results, referral information, and conversational cues to form an initial working hypothesis. Faith described this as forming “a picture of what you think the initial diagnosis is,” while recognising that “it might turn out completely different.”

Context building was central to this process. Nurses reviewed prior history, medication use, and social circumstances before initiating contact, enabling them to anticipate risks and tailor questioning. Jayne described integrating “multifactorial information” into “a larger picture for that particular patient,” while Lee emphasised nursing’s holistic orientation: “Doctors focus on the leg; we care about the whole person.” Accurate history-taking was viewed as foundational, with Rose estimating that “probably 80, if not 90%” of diagnoses of angina were based on history alone.

Skilled questioning compensated for the lack of visual and non-verbal cues. Participants described balancing structured frameworks with conversational flexibility, adapting questions to patient presentation, urgency, and familiarity. Knowing the patient enhanced safety; as Lynn noted, subtle changes in tone or behaviour often signalled deterioration. Some nurses also incorporated multimedia information, such as patient-uploaded videos, to strengthen assessment.

Exploring patients’ hopes, fears, and expectations contextualised clinical decisions and supported person-centred care, though these conversations were often more challenging remotely. Emma described patients withholding information due to fear of consequences, while Jason noted the difficulty of discussing emotionally charged issues without visual cues: “You’re throwing these milestone bombs into a conversation… without the tools to support it.”

Risk assessment was embedded throughout the process. Participants relied on red flags, structured scoring systems, and mental checklists to guide escalation and safety-netting. Rose described routinely providing clear guidance on when patients should seek urgent care, while Echo explained that some red flags were recognised intuitively rather than formally listed.

Nurses also described “zooming in and zooming out”—oscillating between detailed symptom analysis and broader contextual understanding. Faith articulated this process as constantly asking whether “one thing explains everything, or multiple things explain that one thing”. System-level factors, such as local service availability or prescribing practices, further shaped decisions, reinforcing the contextual nature of remote reasoning.

Clinical reasoning in remote contexts was described as layered, iterative, and adaptive. Without physical examination, nurses relied on analytical reasoning, probabilistic thinking, pattern recognition, and intuition to manage uncertainty. Decision-making was rarely linear; instead, nurses continuously generated, tested, and revised hypotheses based on emerging information.

Participants explicitly described working within bounded rationality—making the best possible decision with incomplete information rather than striving for certainty. Sam explained, “I’m concentrating on the information in front of me and making the best decision I can at that time.” Kate reflected on the emotional complexity of such decisions, recognising that both action and inaction carry risk: “Treating and not treating are both risky in their own way.”

Critical thinking was central to safe practice. Nurses described filtering relevant from irrelevant information, balancing thoroughness with efficiency, and weighing risks and benefits proportionately. Experience enabled clinicians to prioritise effectively; as June noted, she learned to “pick out the information that I know I need, rather than getting bogged down.” Over time, reliance shifted from exhaustive data gathering to selective synthesis and narrative coherence.

Hypothesis forming and testing resembled scientific reasoning under time pressure. Claire described ranking differential diagnoses by likelihood, while emphasising the need to remain objective and avoid premature closure. Pattern recognition supported efficiency, with Rose noting that connections “become faster and more automatic the more you see,” though participants stressed that intuition must be continually checked against evidence.

Heuristics and intuition were widely used, particularly in high-risk or ambiguous situations. Jayne described intuition as intrinsic to nursing: “Part of it is just intuition… using your gut and your senses.” Others framed intuition as expert pattern recognition rather than guesswork.

Dan stated, “Instinct is pattern recognition,” grounded in repeated exposure and relational familiarity. However, participants acknowledged that remote contexts constrained intuition due to loss of visual cues, reinforcing the need to corroborate instinct with structured questioning and investigation.

##### 5.2.2.2 Decision-Making Ability: Autonomy, Accountability, and Relationships

Decision-making ability was closely linked to professional autonomy. Nurses described independently managing referrals, prescribing, and complex patient pathways, particularly in specialist and advanced roles. Lynn stated, “We are quite autonomous… it’s quicker if we do them ourselves,” while Kathy linked autonomy to role seniority and confidence.

Autonomy, however, carried significant accountability. Participants described heightened responsibility when making decisions remotely, especially when prescribing or operating near professional boundaries. Ella explained that while responsibility could be shared, “if I’m making a prescribing decision… I do have ultimate responsibility.” Jayne described the “weight of responsibility” as increasing under service pressure and limited follow-up capacity.

Maintaining professional credibility required careful justification and documentation of decisions. Monica described producing “watertight” rationales to support interprofessional trust, while Rose emphasised clearly articulating reasoning to acknowledge uncertainty: “We’re never going to be right 100% of the time.”

Building and sustaining relationships were mentioned as fundamental to effective decision-making. Nurses reported using rapport, conversational engagement, and relational continuity to encourage disclosure and detect subtle cues. Iris described using “chitchat” to foster honesty, while Lynn linked relational familiarity to intuitive insight. Confidence developed through experience but was noted to be tempered by reflexivity. Participants described self-checking, humility, and ongoing learning as safeguards against overconfidence.

Decision-making remained strongly patient-centred. Nurses integrated guidelines, risk tools, and evidence with patient preferences, social context, and feasibility. Jayne described balancing risk scores with patient choice, while Monica noted that guidelines were “a guide, not an absolute.” Shared decision-making was valued but acknowledged as challenging under uncertainty.

##### 5.2.2.3 Knowledge and Experience

Knowledge and experience underpinned all aspects of remote decision-making. Participants viewed expertise as developmental rather than fixed, emerging through education, reflection, mentorship, and exposure. Formal education enhanced confidence and autonomy, but experience was seen as the cornerstone of sound judgement. Jayne highlighted the role of emotional intelligence, while Faith described learning through hindsight and reflection.

Participants reported that mentorship, peer discussion, and psychologically safe learning environments were critical for developing expertise. Participants emphasised that learning often occurred through shared practice rather than formal training, particularly in remote roles.

Personal experiences and intrinsic motivation further shaped decision-making. Nurses described empathy, resilience, and a strong moral drive to “do the right thing” as central to their practice. Annie framed decision-making as requiring courage — “being brave enough to take that step into accountability”— while Jason described moving from feeling like a “glorified answer machine” to an autonomous decision-maker.

Overall, nurses described remote clinical decision-making as a complex, adaptive process that integrates analytical reasoning, intuition, contextual awareness, and relational skills. Decisions were shaped by bounded rationality, requiring nurses to act decisively yet reflectively within uncertainty and system constraints. Experience, confidence, and reflexivity enabled clinicians to balance autonomy with accountability, evidence with empathy, and efficiency with safety. Expertise was portrayed not as certainty, but as the capacity to navigate ambiguity thoughtfully, maintaining person-centred, safe care in the evolving landscape of remote healthcare.

#### 5.2.3 Theme 3: External influencing factors

Nurses described remote clinical decision-making as shaped by a complex interplay of external influences, including clinical decision support resources, teamwork and shared decision-making, workplace culture, stressors, and patient-related factors. These influences did not replace professional judgement but interacted dynamically with nurses’ cognitive processes, shaping how uncertainty, risk, and responsibility were managed in remote care contexts.

##### 5.2.3.1 Clinical Decision Support Resources

Participants consistently described using guidelines, standard operating procedures (SOPs), and digital decision aids as essential scaffolds for safe remote practice. These resources provided structure, reassurance, and consistency, particularly when working autonomously or in uncertain circumstances. However, nurses emphasised that such tools were interpretive rather than prescriptive and required contextual judgement.

As Claire explained, *“the protocol is a guide, not an absolute,”* highlighting the need to adapt recommendations to patient complexity. Echo similarly noted that algorithmic outputs could not replace clinical interpretation: *“It’s a helpful support tool, but it doesn’t replace clinical decision-making or assessment… it’s not the numbers that are wrong; it’s the interpretation that can’t be replaced.”*

Several participants described overriding guideline thresholds when they conflicted with the clinical narrative. Sam reflected on the impracticality of rigid cut-offs: *“Blood results fluctuate—you’ve got to look at trends and what’s feasible in real life.”* Emerging technologies, including artificial intelligence (AI), were viewed cautiously but positively as potential efficiency aids rather than substitutes for professional judgement.

##### 5.2.3.2 Teamwork, Shared Decision-Making, and Peer Support

Team-based working was central to safe remote decision-making. Nurses frequently escalated complex or uncertain cases to multidisciplinary teams (MDTs) for shared reflection and reassurance. Sarah described this collective approach during acute presentations: *“When a patient calls with chest pain, we work as a team—there’s always a doctor within the team.”*

Shared decision-making was portrayed as iterative rather than linear. Tara described remote decisions as evolving over time: *“A remote consultation doesn’t have to be one-stop; sometimes you go back and reflect. Patients appreciate that transparency.”* Nurses valued collaboration not only for patient care but also for distributing responsibility and reducing professional isolation.

Informal peer support was particularly important. Monica explained how post-clinic discussions supported learning and confidence: *“We talk through each patient we’ve had— the decision we made—more as a learning thing.”* Lynn echoed this reassurance-seeking: *“I’ll talk it through with somebody—it’s the right thing to do, but I’m just doubting myself a little.”*

However, interprofessional collaboration was not always seamless. Echo described variable medical support: *“Some doctors are supportive, others dismissive—it’s hit and miss.”* In such cases, nurses relied on documentation and professional assertiveness. Lynn stated clearly, *“If I’m not happy to prescribe something, I’m not doing it—it’s my registration, my NMC pin.”*

##### 5.2.3.3 Stress, Workload, and Workplace Culture

Stress emerged as a pervasive feature of remote decision-making, driven by uncertainty, workload, and fear of error. Ronald described the emotional weight of remote responsibility: *“It can be very stressful… particularly when you know someone’s in real trouble and possibly more trouble than they realise.”*

Nurses experience moderated stress over time. Sam reflected, *“Having seen so many scenarios over the years probably gives that extra bit of assurance.”* Clear protocols and access to senior support were also protective. Dawn explained, *“As long as I can see the bloods are stable and follow algorithms, I don’t feel that concerned about not seeing them face to face.”*

Workplace culture strongly shaped how stress and autonomy were experienced. Tara described her team as *“non-hierarchical and equal,”* attributing confidence to supportive leadership. In contrast, Dan noted how limited organisational recognition constrained autonomy: *“The service doesn’t empower nurses in terms of authority or decision-making.”*

##### 5.2.3.4 Patient Factors

Patient-related factors added further complexity to remote decision-making. Nurses actively triangulated objective data with subjective reports, family input, and contextual cues. Claire distinguished between protocolised and interpretive information: *“If it’s a neutrophil count below one, that’s straightforward… but if it’s something subjective like a rash, I’d want to see them face to face.”*

Family involvement often provided valuable collateral information but could also complicate communication. Jayne noted, *“We’re reliant on carers or family members for a lot of information.”* Interpreting verbal and auditory cues became a critical compensatory skill. Jason captured this uncertainty: *“I’m only going on words and tone of voice… I have to make decisions from the ten per cent I’ve got.”*

Overall, external influencing factors functioned as an interconnected ecosystem surrounding nurses’ cognitive work. Guidelines, teamwork, workplace culture, and patient context interacted continuously with professional judgement. Effective remote decision-making depended not on rigid adherence to frameworks, but on nurses’ ability to interpret, negotiate, and adapt these influences—maintaining safe, person-centred care despite uncertainty.

### 5.3 Integrated Findings

Through iterative synthesis, four integrated pillars were generated:

1. Pillar 1 – Flexibility as the dominant mode: Nurses dynamically shift between intuitive and analytical reasoning to manage uncertainty, illustrating cognitive adaptability within iterative decision processes.
2. Pillar 2 – Education, experience and other internal cognitive shapers: refines analytical reasoning, enhances decision-making capacity, and strengthens reflective judgment in remote settings.
3. Pillar 3 – Environmental enablers and restraints: Team communication and interdisciplinary support influence nurses’ decision-making ability, providing external scaffolding for clinical decision-making.
4. Pillar 4 – Stress as a moderated external influencer: While stress is present in remote work, its effects are buffered by experience, team support, and role autonomy, limiting its impact on decision-making ability.

These pillars represented unified empirical insights that explained how nurses make decisions in remote contexts, integrating both statistical patterns and lived experiences.

#### 5.3.1 Integrated Results: Pillar-Building and Theory-Driven Interpretation

##### 5.3.1.1 Pillar 1 – Flexibility as the Dominant Cognitive Mode

Across the quantitative data, most nurses (73.6%) demonstrated a *flexible* decision-making style, integrating both intuitive and analytical reasoning. Analytical reasoning was significantly associated with decision-making confidence (CDMNS_total: ρ = 0.442, *p* = 0.001), while intuitive reasoning also contributed meaningfully (ρ = 0.384, *p* = 0.005). Qualitative findings converged strongly with this pattern: participants described dynamically shifting between structured and intuitive thought processes, depending on clinical uncertainty and contextual urgency (See ***Appendix*** 4).

##### 5.3.1.2 Pillar 2 – Education, Experience, and Other Cognitive Shapers

Experience, age, and qualification consistently enhanced nurses’ decision-making ability and reflective judgement. Quantitative analysis showed that analytical decision-making increased with years of experience (R² = 0.15, *p* = 0.024), and that age and qualification correlated with evaluation and goal setting in decision-making (ρ = 0.311–0.347, *p* < 0.05). These findings were reinforced by qualitative accounts highlighting the developmental nature of professional reasoning (See ***Appendix*** 5).

##### 5.3.1.3 Pillar 3 – Environmental Enablers and Restraints

Interprofessional collaboration emerged as a key external enabler. Quantitatively, physician– nurse collaboration (PNC_total) significantly predicted decision-making ability in both regression models (B = 1.08, *p* < 0.001; B = 0.99, *p* = 0.003) and correlated with CDMNS subscales related to objective setting and information assimilation. PNC_total is also positively associated with tolerance of uncertainty (ρ = 0.378, p = 0.009), suggesting that supportive interprofessional relationships may be linked to nurses’ comfort in managing clinical uncertainty. Conversely, perceptions of physician authority correlated negatively with decision-making ability (ρ = –0.284, *p* = 0.039), suggesting that hierarchical dominance may constrain autonomy (See ***Appendix*** 6).

##### 5.3.1.4 Pillar 4 – Stress as a Moderated External Influencer

Although stress levels were negatively correlated with decision-making ability, these relationships were not statistically significant. Qualitative data provided insight into this apparent divergence and convergence: participants acknowledged emotional strain and responsibility inherent in remote practice but described mechanisms that buffered stress through experience, team support, and focused coping strategies (See ***Appendix*** 7).

#### 5.3.2 Theory Building (Conceptual Integration)

In the second stage, these empirical pillars were interpreted primarily through the lens of established theories of decision-making: Dual Process Theory (Evans, 2008), Cognitive Continuum Theory (Hammond, 1996; Standing, 2008), Novice to Expert (Benner, 1984), Bounded Rationality (Simon, 1957), and Ecological Bounded Rationality (Gigerenzer, 2008; Todd & Gigerenzer, 2003). Each pillar was aligned with theoretical constructs to explore how empirical evidence supported, extended, or challenged existing models of clinical cognition (***Appendix*** 8). For instance, the coexistence of intuitive and analytical reasoning across datasets reflected DPT and CCT’s principles of cognitive duality and continuity, while the contextual adaptability and satisficing (justifiable) strategies evident in the data aligned with bounded and ecological rationality.

Through this theory-building phase, the integrated findings were elevated from descriptive convergence to conceptual explanation, demonstrating that nurses’ remote clinical decision-making is both bounded by cognitive and environmental constraints and adaptively rational within context. This dual integration process thus generated theory-informed meta-inferences that bridge empirical evidence with established frameworks, contributing to the refinement of decision-making theory within contemporary, technology-mediated clinical practice.

This integration aligns with CCT and DPT, illustrating how analytical (System 2) and intuitive (System 1) reasoning operate interwoven rather than in isolation. In remote decision-making contexts, flexibility represents an adaptive balance between heuristic recognition and evidence-based evaluation. Quantitative findings demonstrated that both analytical and intuitive reasoning significantly predicted nurses’ clinical decision-making ability, and most participants adopted a flexible reasoning style. These results align with CCT’s assertion that effective decision-making depends on adaptive movement along the continuum rather than exclusive reliance on either pole. The qualitative findings reinforced this view: nurses described shifting between intuitive recognition and analytical verification depending on task demands, information availability, and time pressure. In remote care contexts, where uncertainty and incomplete data are common, this cognitive flexibility allowed nurses to integrate experiential knowledge with structured reasoning.

The positive association between analytical and intuitive reasoning (r = 0.762, p < 0.001) suggests that nurses with stronger analytical skills also demonstrate more effective intuitive reasoning. Rather than representing opposing processes, these modes appear to function synergistically in clinical decision-making. This aligns with evidence that experienced clinicians integrate analytical knowledge into intuitive judgments, supporting a flexible, dual-process model and a bidirectional relationship between reasoning styles (Hammond, 2000; Croskerry, 2009).

The concept of bounded rationality is reflected in the quantitative findings, which indicate that either the analytical or the intuitive model explains at most 31% of the variance in decision-making ability, suggesting that other factors, including clinicians’ internal and external contextual factors, are not accounted for in the model. In qualitative data, nurses reported limited time, information and resources that restrained them from an ideal decision but a ‘justifiable’ decision (Jason), or a ‘best decision at the time’ (Sam). It is also noted that expertise refines decision heuristics over time, reducing cognitive load through pattern recognition and experiential learning. Nurses’ growing clinical knowledge and academic learning expanded their decision boundaries, fostering autonomy and confidence in complex remote environments.

The overall findings show convergence with the ecological rationality process, where decision-making quality is not solely an individual cognitive product but a function of environmental adaptation. Effective collaboration, shared education, and professional trust extend nurses’ cognitive capacity through collective reasoning, while hierarchical constraints may narrow the decision space.

The findings also suggest that stress functions as a moderated rather than direct influence. When autonomy, competence, and support are present, stress does not undermine nurses’ decision-making ability in remote reviews but may instead promote heightened attentional control and situational awareness.

#### 5.3.3 Conceptualising a framework for CDM via remote reviews

A conceptual framework for CDM in remote reviews, the Integrated Cognitive-Environmental Framework of Remote Clinical Decision-Making (ICE—later may also be referred to as the ICE framework), was developed to summarise insights from decision-making theories and to integrate findings. This framework proposes two systems involved in CDM through remote reviews: the internal system within the clinician’s mind and the external system in which the clinician operates. The data indicate that the clinician employs both intuitive and analytical thinking to process information and uses a cyclical and iterative searching strategy (anchoring and sufficient/insufficient adjusting) to make decisions about the patient management plan. The external environmental system encompasses the environment in which decisions occur, including clinical decision support resources (e.g., guidelines, protocols, computer algorithms, peer/doctor/multiple disciplinary support), patients or their proxies, and external influencers. These external elements collectively present as the ‘task’ that activates and shapes the clinician’s internal decision-making process, which integrates both intuitive and analytical thinking, and involves a cyclical, iterative search strategy where initial judgments (‘anchors’) are refined (‘adjusted’) as new information emerges, capable of moving both forward and backwards.

In this study, a good remote clinical decision was conceptualised as a “decision match”: a decision that was safe, professionally justifiable, and contextually appropriate because the clinician’s internal reasoning was sufficiently aligned with external realities such as patient factors (“I’d be thinking about what’s most likely based on history, presentation, and risk factors” (Claire).), patient preferences (“they agree on a compromise: a six-week trial of lifestyle changes followed by a review,” and any future decision is described as “collaborative, and data driven.” (Jason)), available evidence and protocols (“…I’ve got protocols to back me up and senior colleagues to speak to.” (Claire)), collaborative professional input (“…that’s part of your credibility building—you feel certain of your ground with the doctor.”), and service conditions (“Now we’ve got to be adaptive and figure out the safety netting around that system.” (Ella)). This interpretation arose from the integrated findings, in which participants repeatedly described making “the best decision… at that time” or “the safe one with the information I had at the time.” This also includes respecting patient autonomy (see code of “shared decision-making”), engaging with appropriate clinical guidelines (“…as long as I can see the bloods are stable and follow algorithms, I don’t feel that concerned about not seeing them face to face.” (Dawn)), ethical and legal principles (“…if I’m not happy to prescribe something, I won’t do it. I’ll document why—it’s my accountability.” (Lynn)), and research evidence, and navigating the practicalities of the care environment. The framework’s content is in ***Appendix*** 9. The definitions of terms can be found in ***Appendix*** 10, following the diagram of the ICE framework in ***Figure*** 4. Further supporting evidence can be found in ***Appendix*** 11.

**Figure 4.**
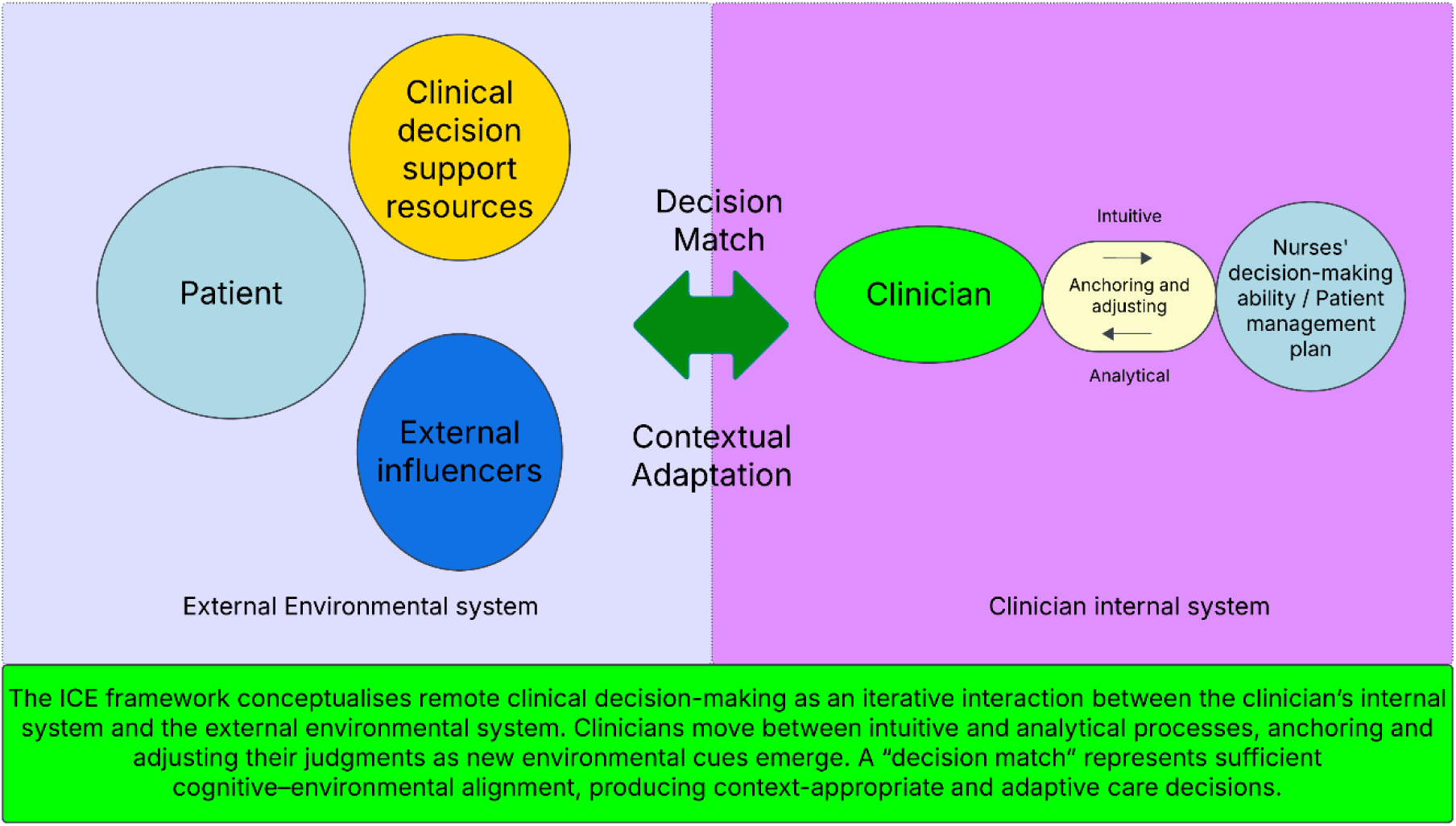
A Conceptualised Integrated Cognitive-Environmental (ICE) Framework for CDM during Remote Review.

## 6. Discussion

This mixed-methods study examined how registered nurses make clinical decisions in remote care environments and how cognitive processes interact with digital, social, and organisational contexts. The findings demonstrate that remote clinical decision-making is neither purely analytical nor purely intuitive, but emerges from a dynamic interaction between internal reasoning processes and external environmental structures. This interaction is conceptualised through the Integrated Cognitive–Environmental (ICE) Framework, which positions decision-making as an adaptive system rather than a sole individual cognitive act.

Consistent with bounded rationality (Simon, 1957), nurses made decisions under conditions of incomplete information, time pressure, and cognitive limits. Participants described using intuition—often framed as pattern recognition or a “sense” of concern—as an initial anchor, followed by analytical validation through investigations, protocols, or multidisciplinary discussion. This anchoring-and-adjustment process aligns with Tversky and Kahneman’s (1974) model and reflects hypothetico-deductive reasoning that integrates intuitive and analytical modes (Banning, 2008). One nurse explained: *“I do use intuition a lot… but I do not rely on it; I base my decisions on clinical investigations”* (Lee), illustrating deliberate cross-checking rather than uncritical reliance on intuition.

Quantitative findings showed that most nurses employed a flexible, quasi-rational decision-making style, supporting Hammond’s (1996) Cognitive Continuum Theory. However, years of experience were significantly associated with greater analytical reasoning. This contrasts with earlier studies suggesting that experience promotes intuitive thinking (Pretz & Folse, 2011) and may reflect the nature of remote practice, where limited sensory input reduces the reliability of intuitive cues. As nurses gain experience in digital environments, they appear to compensate for perceptual loss by engaging more deliberately in analytical reasoning, consistent with dual-process theory (Evans, 2008).

Importantly, decision-making in remote care was not confined to the individual nurse. Cognitive work was distributed across electronic patient records, decision-support tools, multidisciplinary teams, and relational memory of patients, reflecting distributed cognition (Hutchins, 1995). Quasi-rationality was therefore also externally scaffolded rather than embodied solely through direct sensory experience.

Environmental structure strongly influenced cognitive mode selection. When digital systems provided clear, structured data, analytical reasoning predominated; when information was ambiguous or fragmented, nurses relied more on experiential heuristics. This adaptive calibration aligns with ecological bounded rationality, which proposes that reasoning strategies are tuned to the structure of available information rather than universally optimal (Gigerenzer, 2008). Within the ICE framework, decision quality depends on the degree of alignment between internal cognition and environmental affordances.

Physician–nurse collaboration emerged as a strong predictor of perceived decision-making ability, corroborating previous findings that interprofessional collaboration enhances clinical judgment and patient outcomes (Manojlovich & DeCicco, 2007; Bulut *et al.,* 2024). Nurses described collaboration as providing reassurance, shared accountability, and access to broader expertise. In remote contexts, collaboration also functioned as a form of distributed cognition, supporting sense-making across professional and technological networks.

Contrary to much literature linking stress to impaired clinical performance (Croskerry *et al.,* 2013; Russ *et al.,* 2018; Tam *et al.,* 2025), stress was not a significant predictor of decision-making ability in this study. Instead, experienced nurses appeared to regulate stress adaptively, using reflective caution and analytical checking rather than impulsive decision-making. This finding aligns with evidence that experienced clinicians can maintain decision quality under moderate stress (LeBlanc, 2009) and supports the Yerkes–Dodson law, which suggests optimal performance occurs under moderate arousal (Yerkes & Dodson, 1908). The structured collaborative nature of remote systems and access to protocols and support may buffer the cognitive effects of stress in remote settings.

The findings also reframe expertise as contextually calibrated cognition. Drawing on Benner’s (1984) Novice-to-Expert framework, experienced nurses demonstrated adaptive expertise— the ability to flexibly integrate intuition and analysis in response to environmental demands (Hatano & Inagaki, 1986). Within the ICE framework, expertise is not solely a function of accumulated knowledge but also of ecological attunement to patients’ presentations and preferences, digital systems, organisational structures, and collaborative networks.

These findings have important implications for practice, education, and policy. Remote decision-making should be recognised as a specialist skill requiring explicit training in digital cognition, communication, and metacognitive regulation. Educational approaches should move beyond procedural reasoning to foster cognitive flexibility, ecological awareness, and reflective practice (Hammond, 1996; Croskerry, 2009). Organisationally, decision quality is strengthened when there is a good fit between the cognitive demands of remote clinical decision-making and the work environment in practice. For example, nurses are better able to make safe and appropriate decisions when digital systems support rapid access to relevant patient information, workloads allow time for assessment and critical thinking, and team cultures enable collaboration and escalation in situations of uncertainty.

These findings are particularly relevant to digital health because they demonstrate that technology-enabled care is not just about adopting remote monitoring platforms or virtual care pathways. Instead, digital health systems influence how clinicians observe, interpret, prioritise, and respond to patient information. The study indicates that the success of digitally enabled care models depends on more than just technological functionality; it also relies on whether these systems support clinical judgment, interdisciplinary communication, and safe escalation in uncertain situations. This is especially crucial in remote review settings, where nurses may need to make decisions without direct physical assessment and must depend on digitally mediated information, patient narratives, and collaborative support.

Although this study was not explicitly designed around the Fundamentals of Care framework, several findings are conceptually relevant to it. Participants described constructing a holistic view of the patient, taking into account social context and practicality, exploring patients’ hopes, fears, and expectations, and utilising rapport and relational continuity to support assessment and disclosure. These findings imply that remote nursing decision-making extends beyond biomedical judgement alone, involving relational and person-centred work that may underpin assessment of fundamental care needs. However, since the study tools and analytic framework primarily focused on clinical decision-making, we were unable to systematically determine how nurses identify, prioritise, and respond to patients’ fundamental care needs remotely. Future research should therefore explicitly apply the Fundamentals of Care framework to investigate how physical, psychosocial, and relational care needs are recognised, negotiated, and addressed during remote nursing encounters.

## 7. Implications

### 7.1 Implications for Clinical Practice, Education, Policy, and Research

This study demonstrates that safe and effective remote clinical decision-making depends on alignment between nurses’ cognitive flexibility and the digital, organisational, and relational environments in which care is delivered. Nurses must integrate analytical reasoning, intuition, and communication within digitally mediated systems while maintaining professional autonomy and collaborative working.

### 7.2 Clinical Practice and Interprofessional Collaboration

Remote decision-making was consistently framed as collaborative rather than prescriptive. Even when objective data were available, nurses emphasised shared decision-making with patients and colleagues. As Tara noted, *“It is always shared decision making… there are very few situations where we will say you really need to take this tablet.”* Quantitative findings showed that nurses’ perceived autonomy was positively associated with decision-making ability, while overall physician–nurse collaboration supported system-level coordination rather than individual cognition. These findings highlight the importance of collaborative autonomy, whereby nurses exercise independent judgement within supportive interprofessional structures, rather than working in isolation.

This study advances understanding of remote nursing clinical decision-making, but it also highlights an important area requiring further attention: how nurses assess and address patients’ fundamental care needs in remote encounters. Future research should examine remote decision-making through an explicit fundamentals-of-care lens, making person-centred fundamental care, as well as clinical reasoning, more visible in digitally mediated practice.

### 7.3 Communication, Uncertainty, and Cognitive Load

The loss of non-verbal cues in remote reviews altered how nurses balanced intuition and analysis. Participants described relying heavily on tone, phrasing, and narrative coherence to interpret patient status. Jason reflected, *“I’m only going on words and tone of voice… I have to make decisions from the ten percent I’ve got.”* Rather than viewing uncertainty as a deficit, nurses employed adaptive strategies such as iterative validation, prioritisation, and heuristic simplification. These practices reflect bounded rationality as a safety mechanism, supporting resilience and decision quality under constrained conditions.

### 7.4 Digital Systems and Cognitive–Environmental Alignment

Findings underscore the role of digital systems as active components of nurses’ cognitive environments. Well-designed dashboards, structured templates, and remote monitoring data supported analytical reasoning, whereas fragmented systems increased cognitive load and reliance on intuition. The ICE framework highlights the need for co-design between clinicians and digital developers to ensure cognitive–environmental coherence and reduce avoidable decision strain.

### 7.5 Implications for Education and Workforce Development

Remote consultation was described as a learned skill requiring deliberate training. Tara emphasised, *“Remote consultations… is a skill that I’ve had to learn on top of all my other skills.”* Education enhanced analytical reasoning, while experience supported intuitive, context-sensitive judgement, aligning with Benner’s novice-to-expert trajectory. Nursing education should therefore prioritise cognitive adaptability, teaching students how to move flexibly along the cognitive continuum. Simulation-based learning, reflective supervision, and mentorship are critical for developing tolerance of uncertainty and metacognitive awareness in remote contexts.

### 7.6 Policy, AI, and Organisational Implications

Remote decision-making should be recognised as a distinct area of professional expertise within policy and competency frameworks. The ICE framework offers a conceptual tool for designing governance structures that balance standardisation with professional discretion. As AI and decision-support systems become embedded in care, nurses must be supported to engage with them as cognitive partners rather than prescriptive authorities. As Sam noted, *“AI can help with the simple stuff… but not for complex decisions where you need to understand the person.”*

Overall, the findings affirm that effective remote clinical decision-making is an ecologically embedded process shaped by cognition, technology, collaboration, and context. Investment in nurse education, digital system design, and psychologically safe interprofessional cultures is essential to sustain adaptive expertise and patient safety in increasingly remote and data-driven healthcare systems.

## 8. Limitations, Future Directions, and Conclusion

This study provides new theoretical and empirical insights into nurses’ clinical decision-making in remote care environments; however, several limitations should be acknowledged. First, participants were recruited from UK NHS settings with established digital infrastructures, and most nurses worked in hybrid roles combining remote and face-to-face care. The relatively small quantitative sample (n = 53) and the contextual specificity of the setting may limit generalisability to healthcare systems with different levels of digital maturity, professional autonomy, or organisational support. Future research should therefore test the Integrated Cognitive–Environmental (ICE) framework across diverse international and organisational contexts.

Second, the cross-sectional design of the quantitative phase limits causal inference. Although associations between reasoning styles, collaboration, stress, and perceived decision-making ability were identified, longitudinal research is required to examine how these relationships evolve with increasing experience in remote practice. The qualitative component, while offering rich insight, relied on self-reported accounts and may not fully capture subconscious or automatic cognitive processes. Future studies could strengthen ecological validity by incorporating observational methods, real-time decision logs, or digital trace data.

Although 81 nurses completed the survey, only 53 had sufficient item completion for composite-scale analyses, which may have reduced statistical power and introduced response-related selection bias. Some questionnaire subscales demonstrated low internal consistency, particularly within the CDMNS and selected PNC subscales, and these subscale-level findings should therefore be interpreted cautiously. Greater interpretive weight should be placed on total-scale findings, regression models, and the integrated mixed-methods inferences. In addition, the quantitative outcome measured perceived decision-making ability through self-report rather than directly observed decision quality or patient outcomes, which may limit conclusions about actual performance in practice.

Despite these limitations, the mixed-methods design enabled robust triangulation, integrating quantitative modelling with qualitative interpretation to develop a theoretically grounded framework. The ICE framework advances understanding of clinical decision-making by conceptualising it as a dynamic cognitive–environmental system rather than an individual mental process. Findings demonstrate that remote decision-making is distributed across clinicians, technologies, organisational structures, and patients, challenging reductionist views of intuition and analysis.

The study has implications for education, practice, research and policy. Remote clinical decision-making should be recognised as a specialist competence requiring cognitive flexibility, digital literacy, and collaborative skills. By foregrounding ecological bounded rationality and human–technology alignment, the ICE framework offers a foundation for designing training programmes, digital systems, and governance structures that support adaptive, safe, and reflective nursing practice in increasingly remote and data-driven healthcare environments.

## Data Availability

All data produced in the present study are available upon reasonable request to the authors

## Patient or Public Contribution

Patient and public representatives contributed to stakeholder discussions that informed the development of the interview topic guide and the theoretical model. Patients or members of the public were not involved in recruitment, data collection, analysis, interpretation of findings, or preparation of the manuscript.

## Appendix 1

### Table Data collection

| Data type | Data and its measure |
| --- | --- |
| **Demographic Data** | Gender, age, ethnicity, professional qualification, years of experience in nursing, frequencies of conducting the remote review, job title, job banding, and speciality |
| **Quantitative data** | Independent variable 1: Nurses’ clinical decision-making process---Decision-making process questionnaire (Lauri & Salanterä, 2002)<br>Independent variable 2: physician-nurse collaboration --- Jefferson scale of attitudes toward physician-nurse collaboration (JSAPNC) (Hojat & Herman, 1985)<br>Independent variable 3: Decision-making stress --- Decision-making stress questionnaire (Bucknall & Thomas, 1997); |
|  | one item question measuring tolerance of uncertainty and fear of making mistakes.<br>Dependent variable/outcome: Nurses' decision-making ability --- clinical decision-making in nursing scale (CDMNS) (Jenkins,1985) |
| **Qualitative data** | Semi-structured interviews with key questions:<br>- How do you make clinical decisions in remote reviews? Can you give me a recent example of how you make clinical decisions in remote reviews?<br>- How do you interact with doctors when making a clinical decision, or how are doctors involved in the discussions about your clinical decision?<br>- How do you find making clinical decisions? How stressful do you find it?<br>- What affects your ability to make clinical decisions in remote review? |

## Appendix 2

### Table Two questions relating to uncertainty and fear of making mistakes

Appendix 2 **Table Two questions relating to uncertainty and fear of making mistakes**
| Questions relating to facing uncertainty and fear of making mistakes | Answering options |
| --- | --- |
| How do you evaluate your tolerance of uncertainty when making decisions? | a. I have difficulties in tolerating uncertainty in decision making.<br>b. I tolerate uncertainty quite well in decision making.<br>c. I tolerate uncertainty very well in medical decision making. |
| How often do you fear making mistakes in decision-making (or making decisions) | Several daily daily At least once a week At least once a month At least once a year Never |

## Appendix 3

### Table of Demographics for the Interview Participants

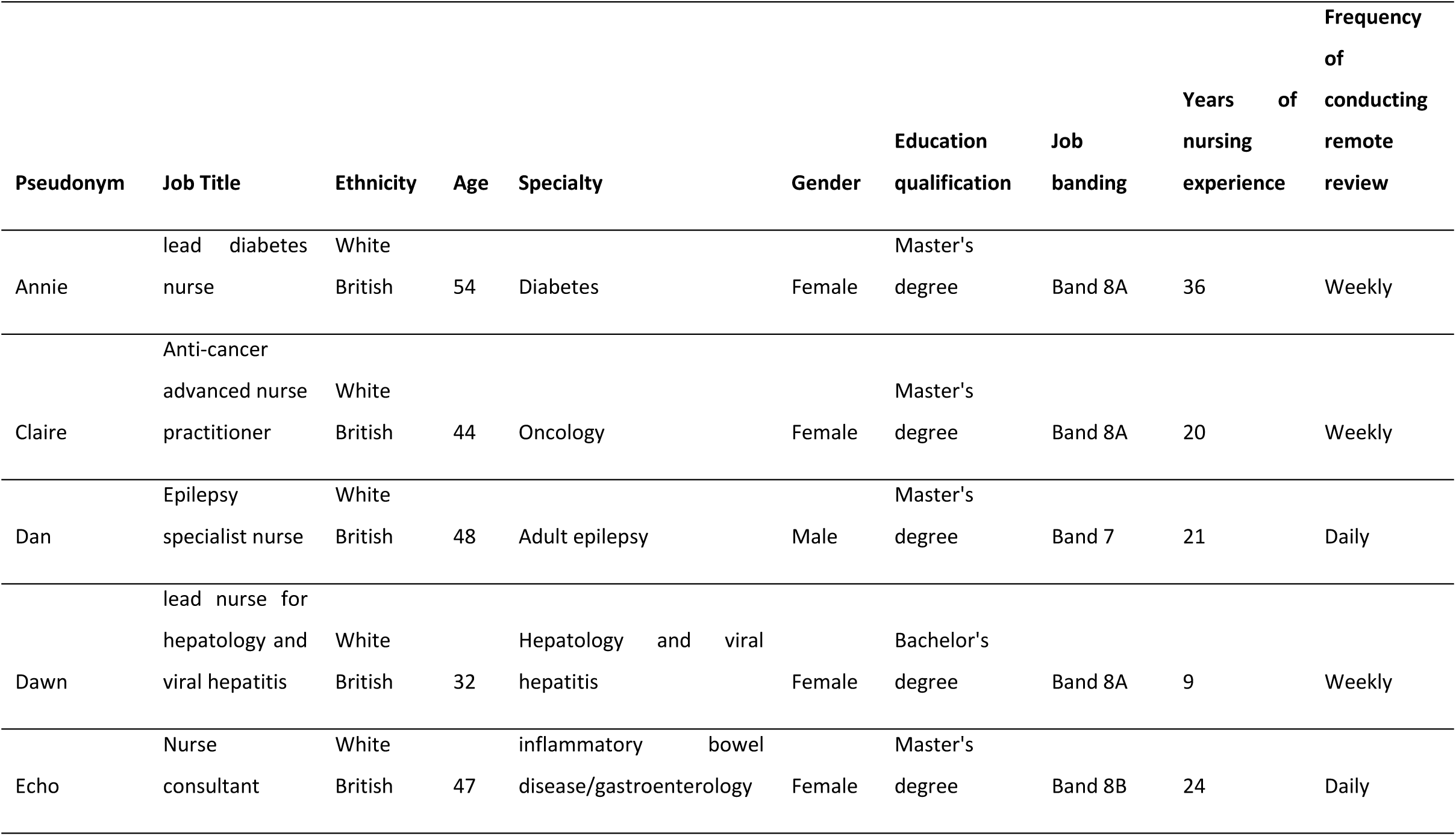
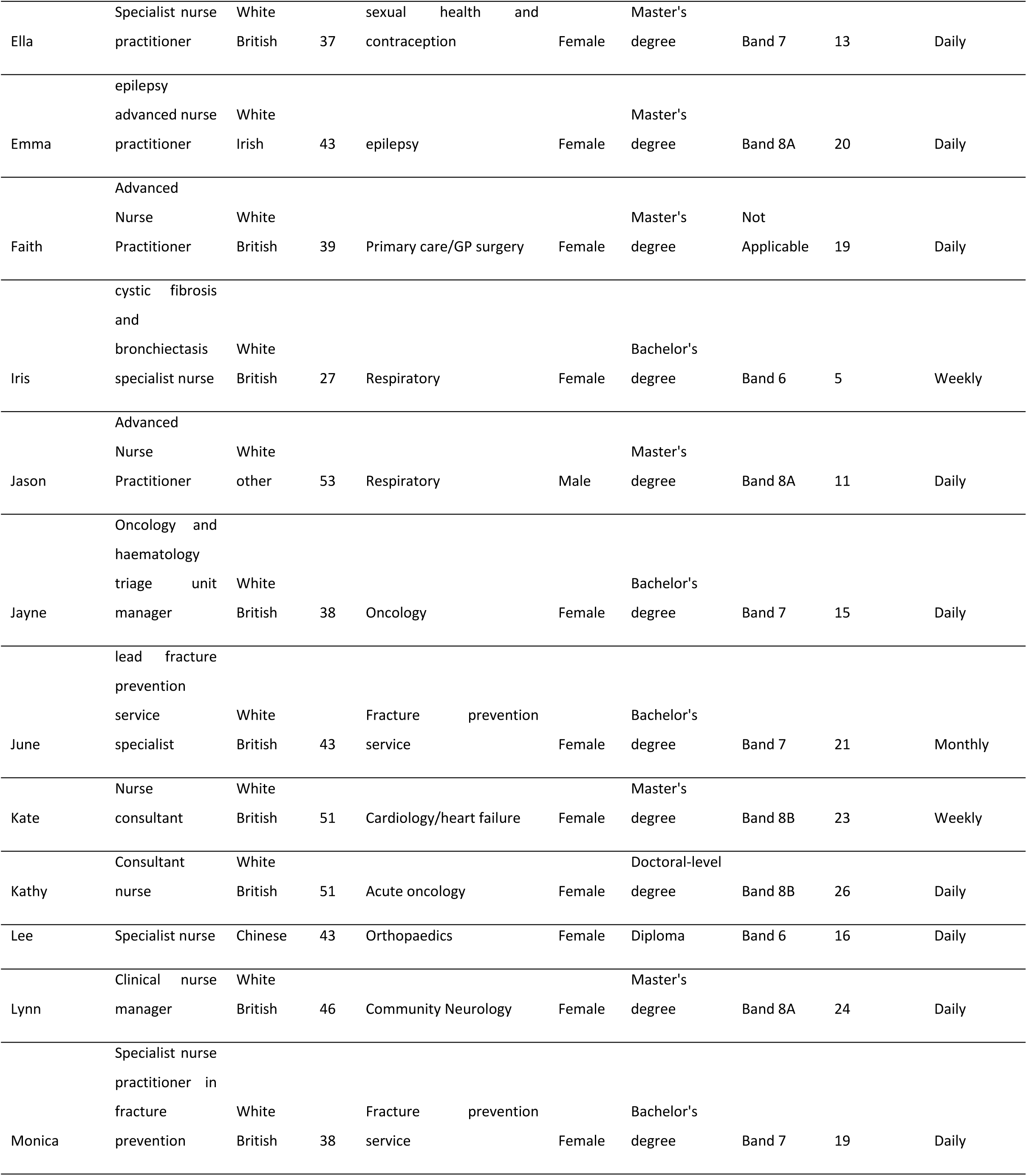
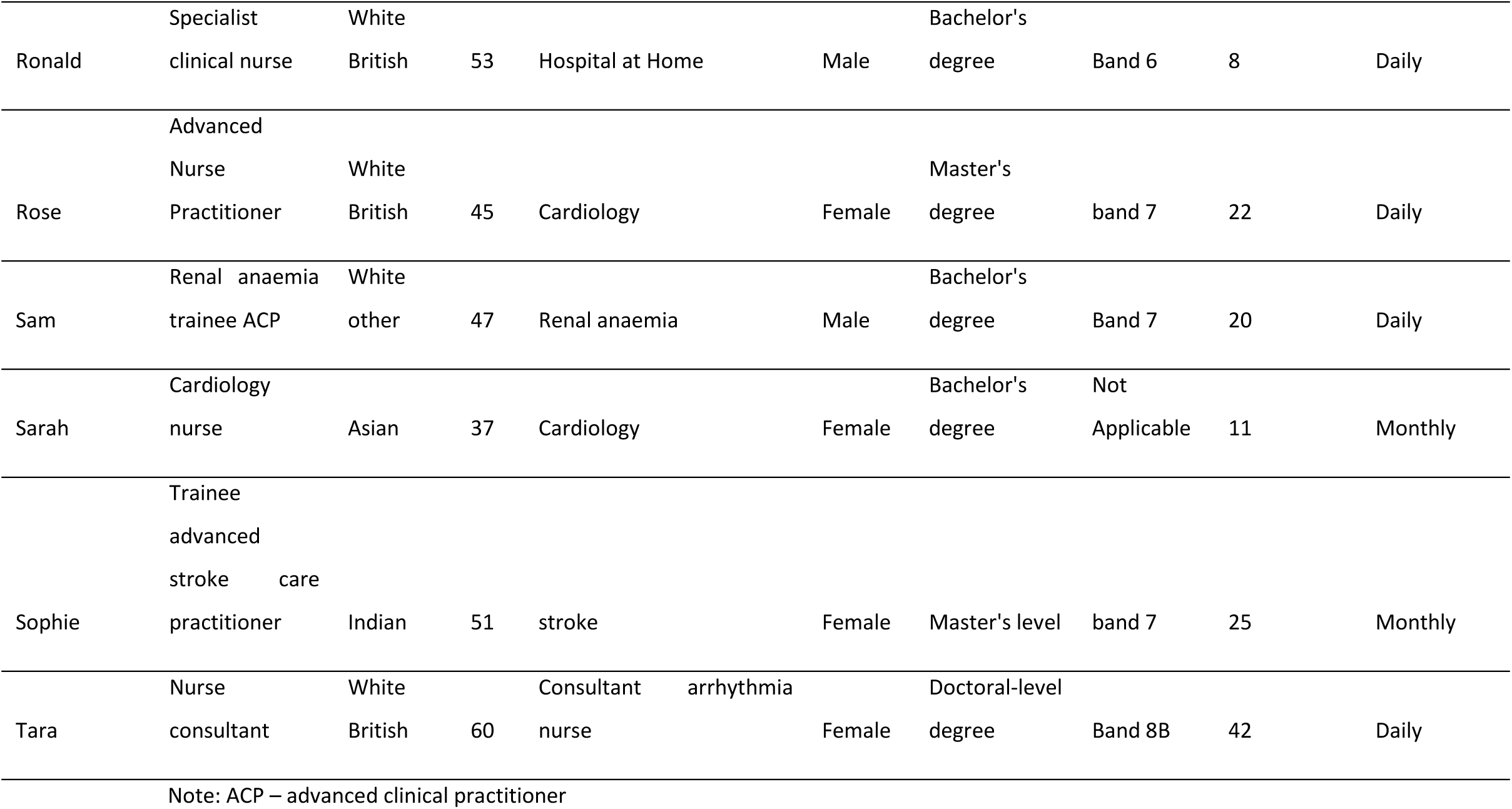

## Appendix 4

### Table Pillar Building for Mixed -method Evidence Synthesis (Pillar One)

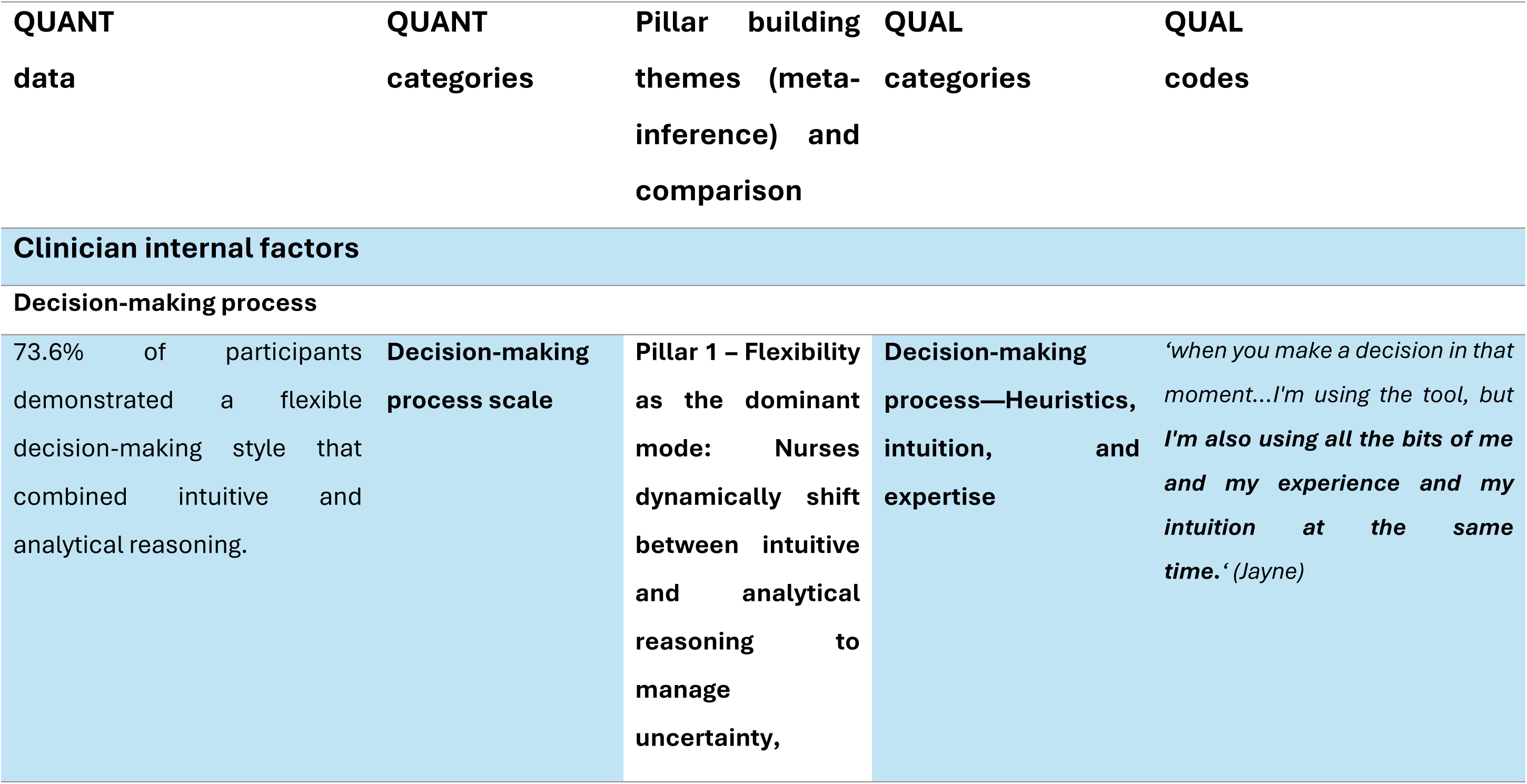
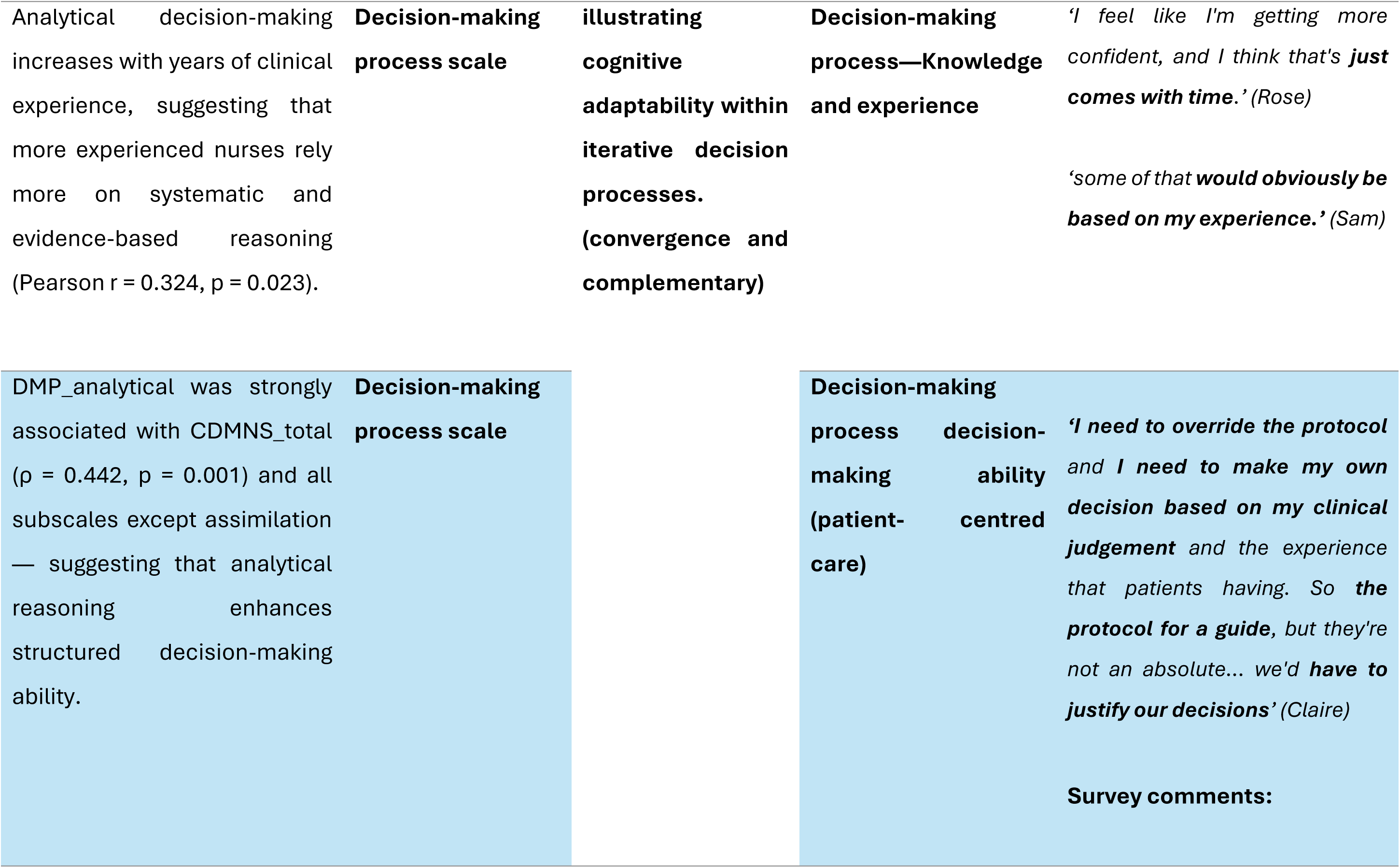
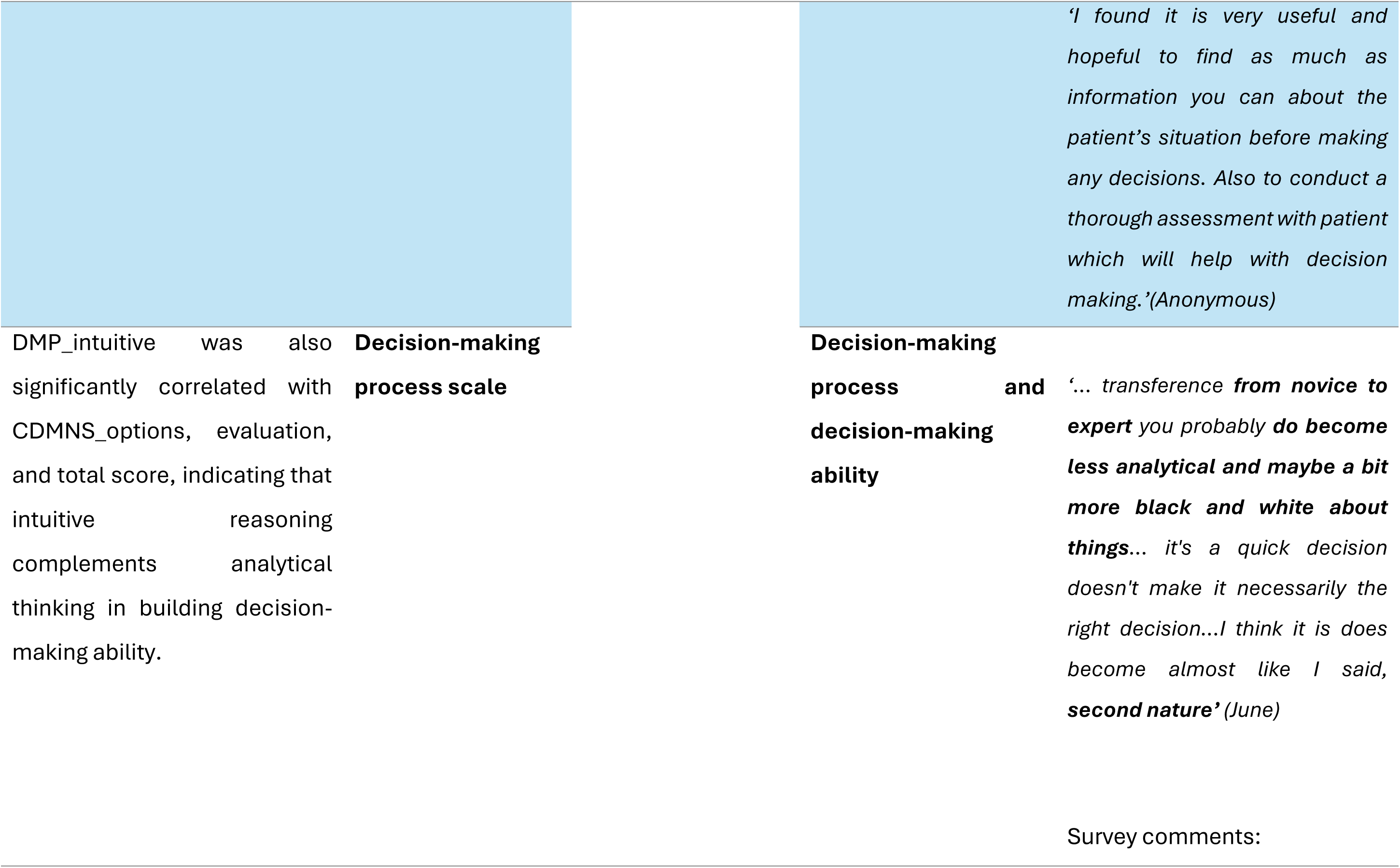
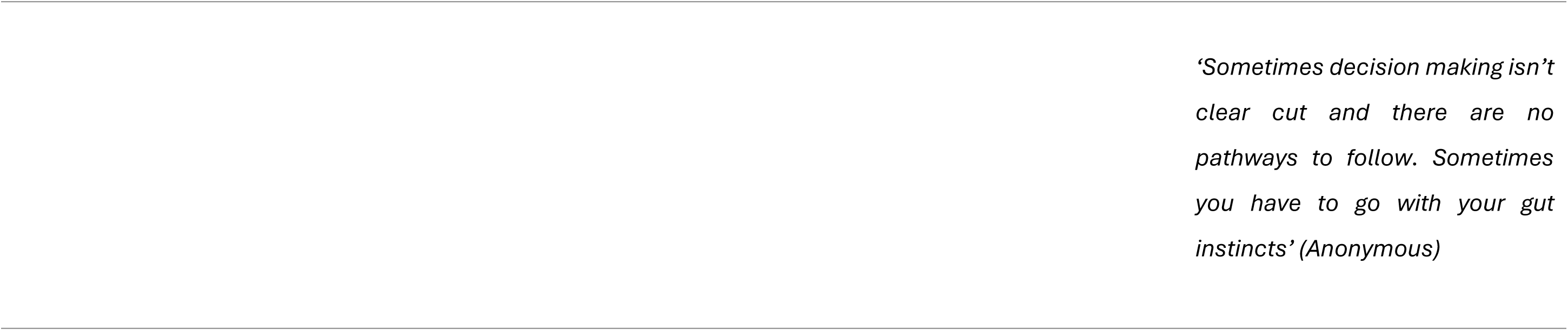

## Appendix 5

### Table Pillar Building for Mixed-method Evidence Synthesis (Pillar Two)

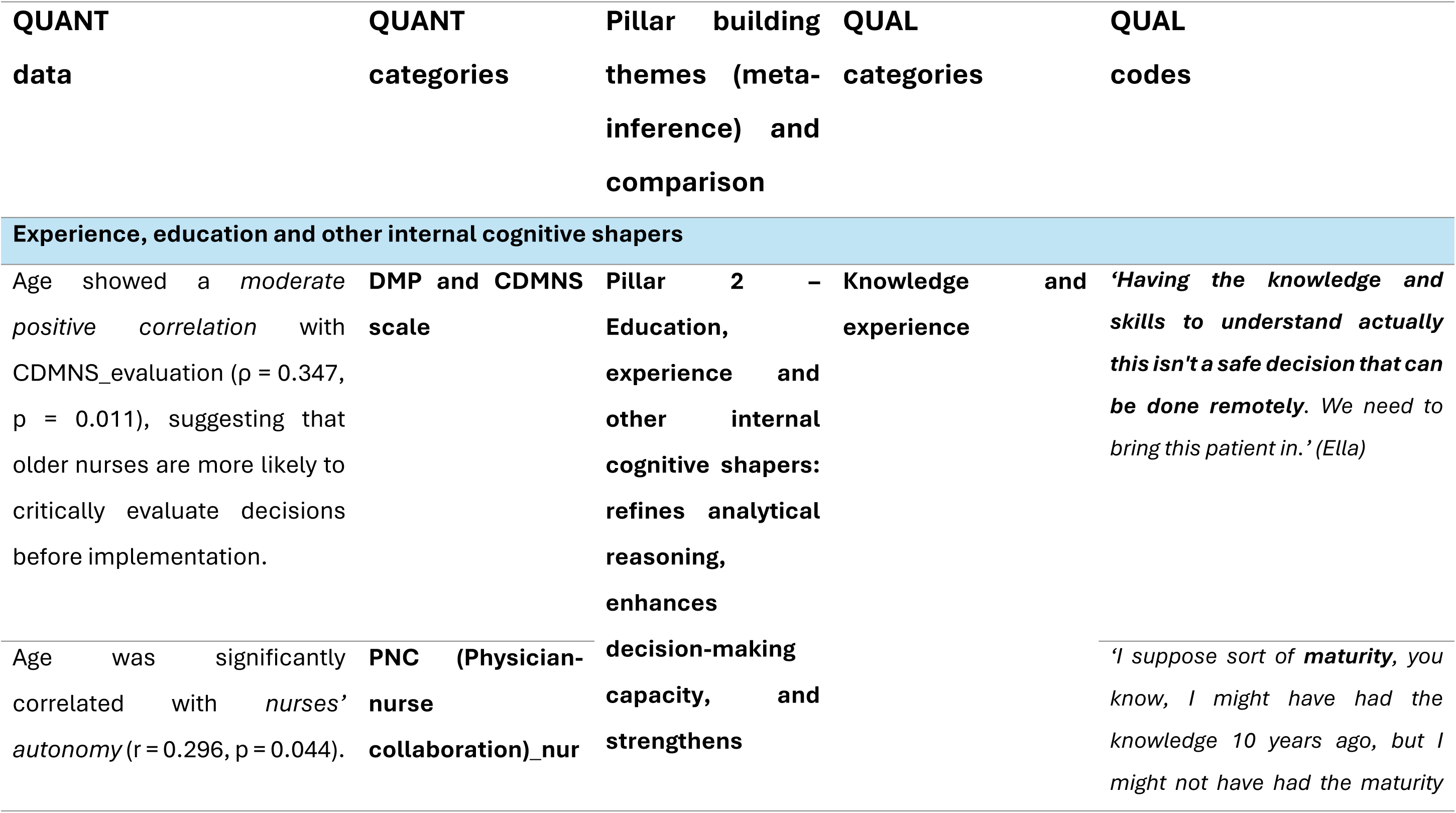
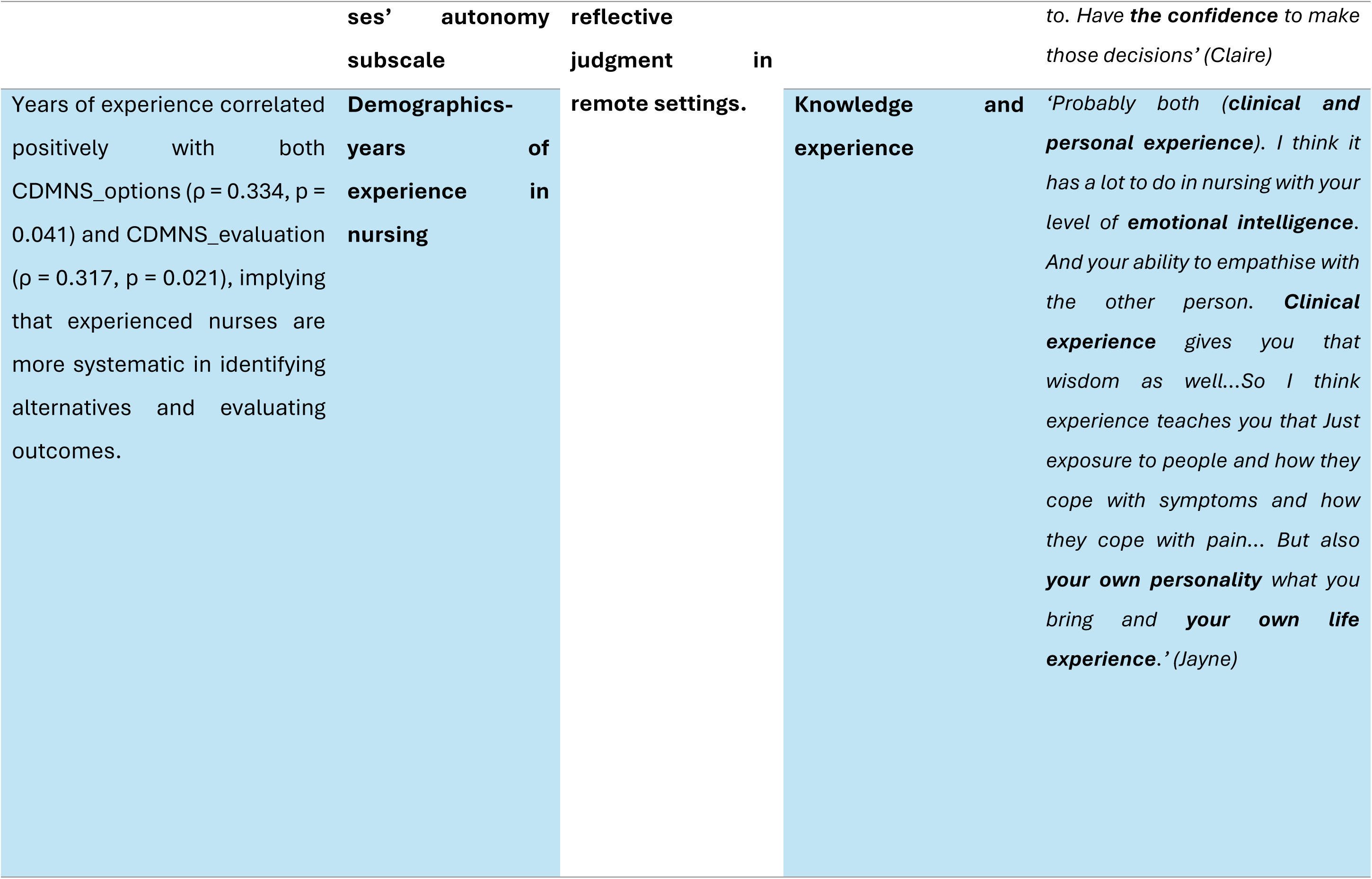
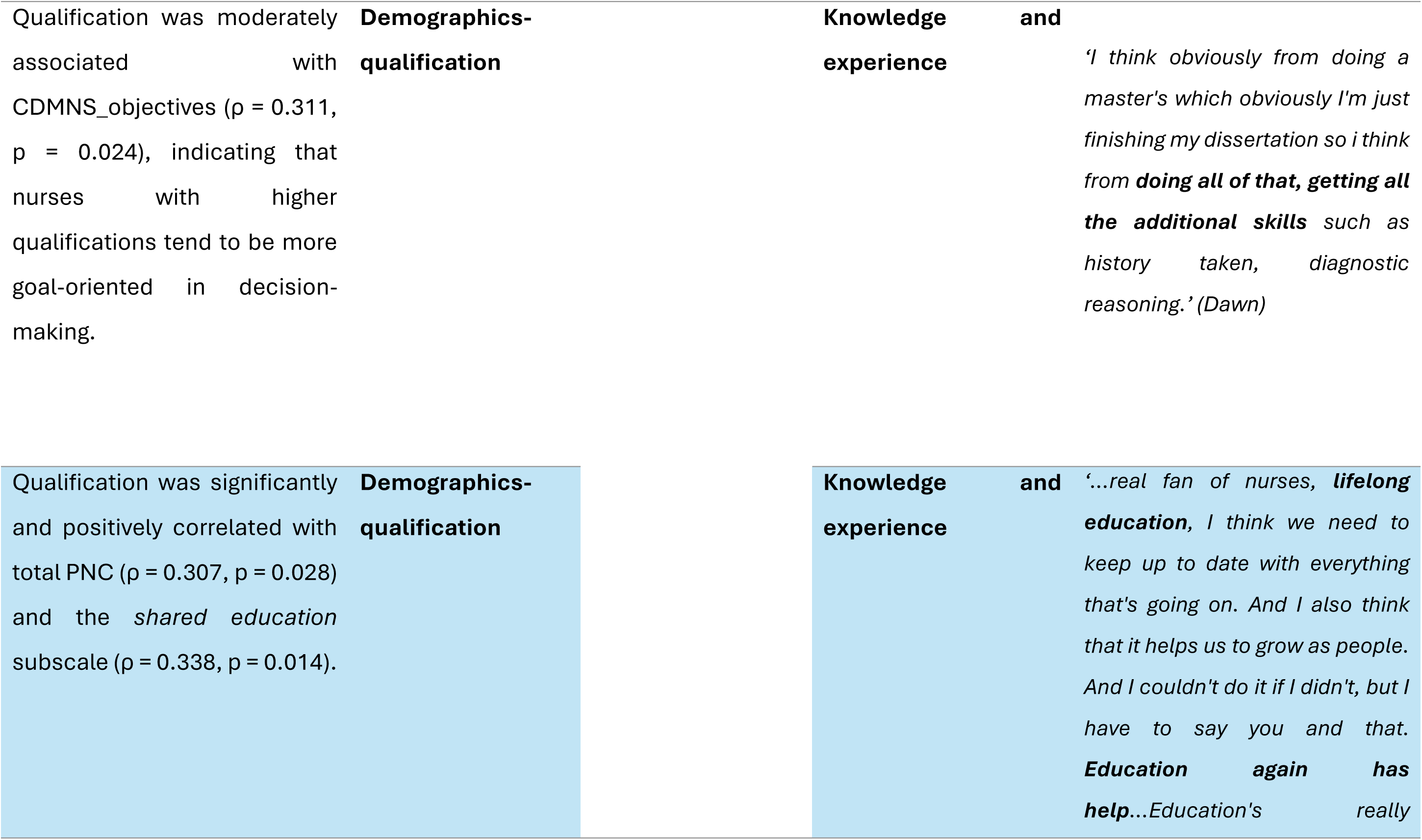
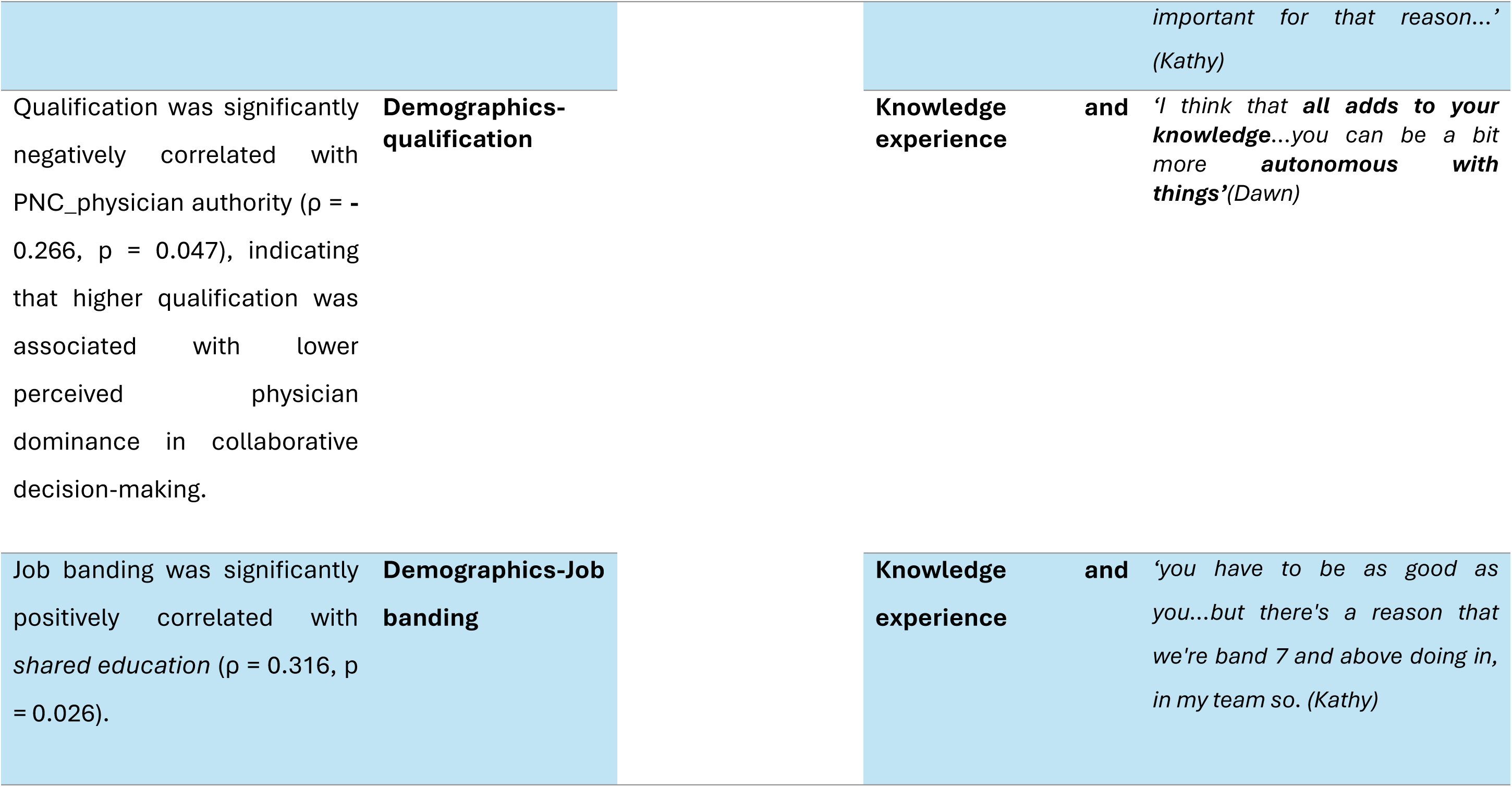

## Appendix 6

### Table Pillar Building for Mixed -method Evidence Synthesis (Pillar Three)

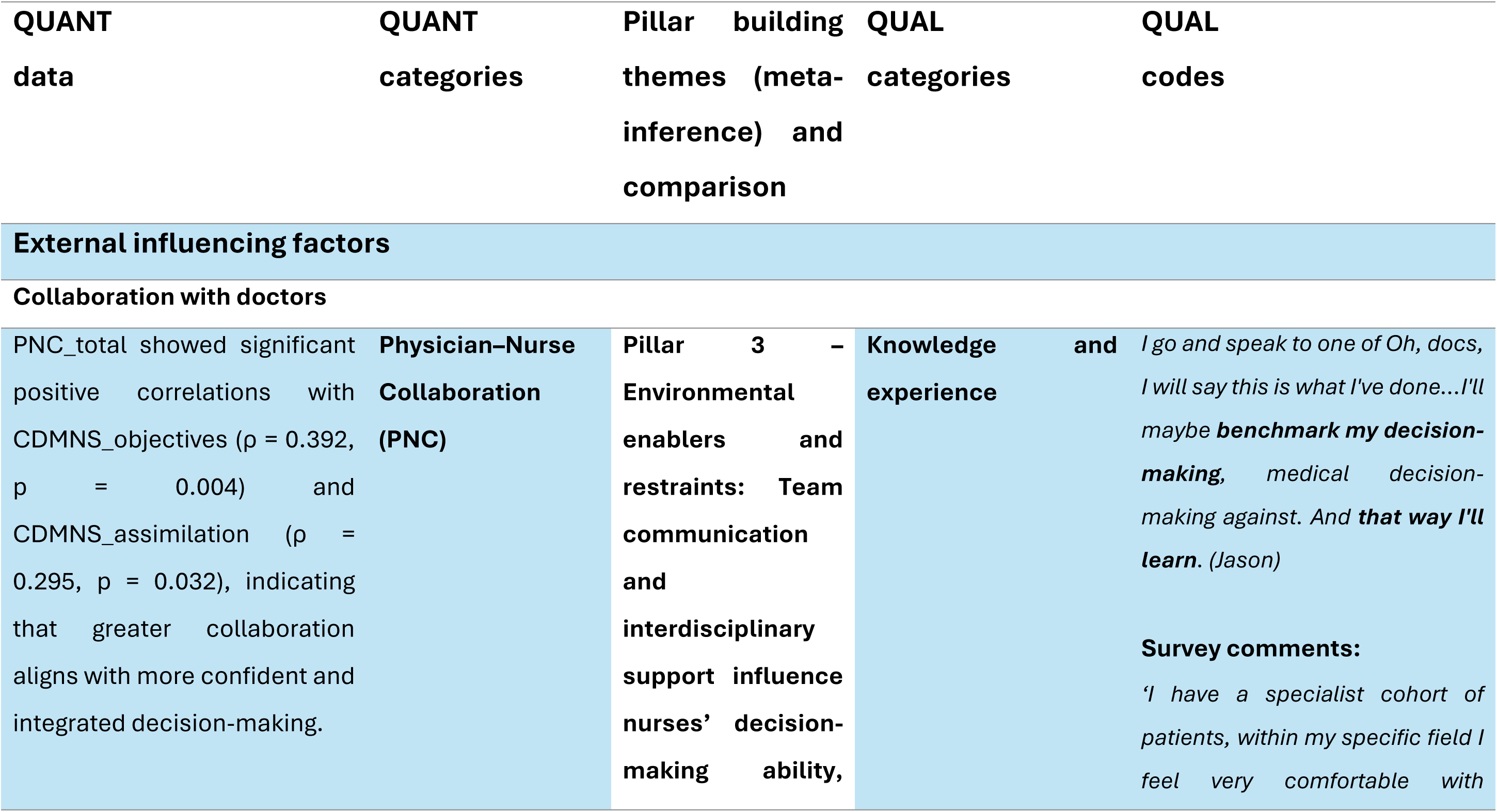
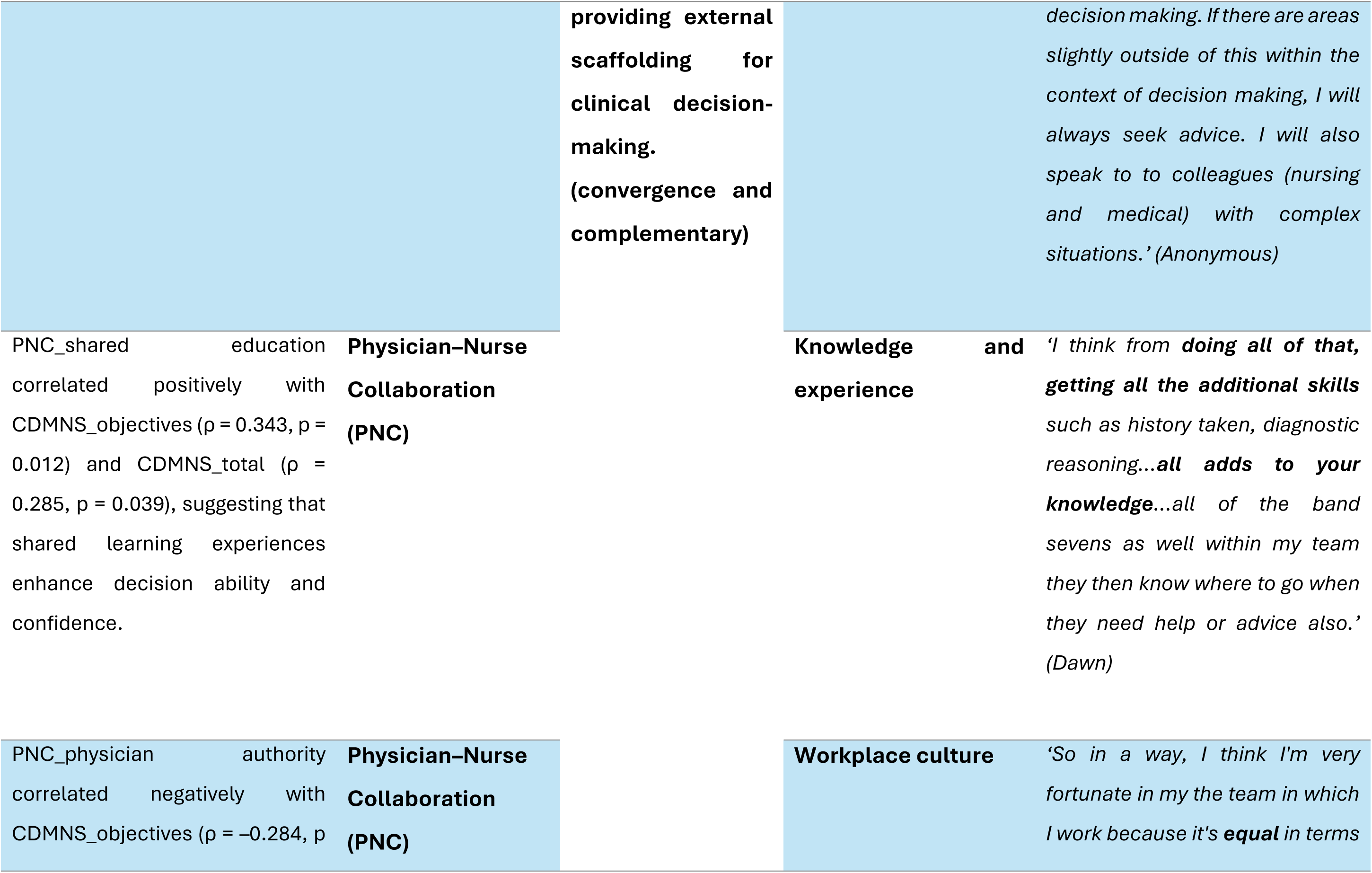
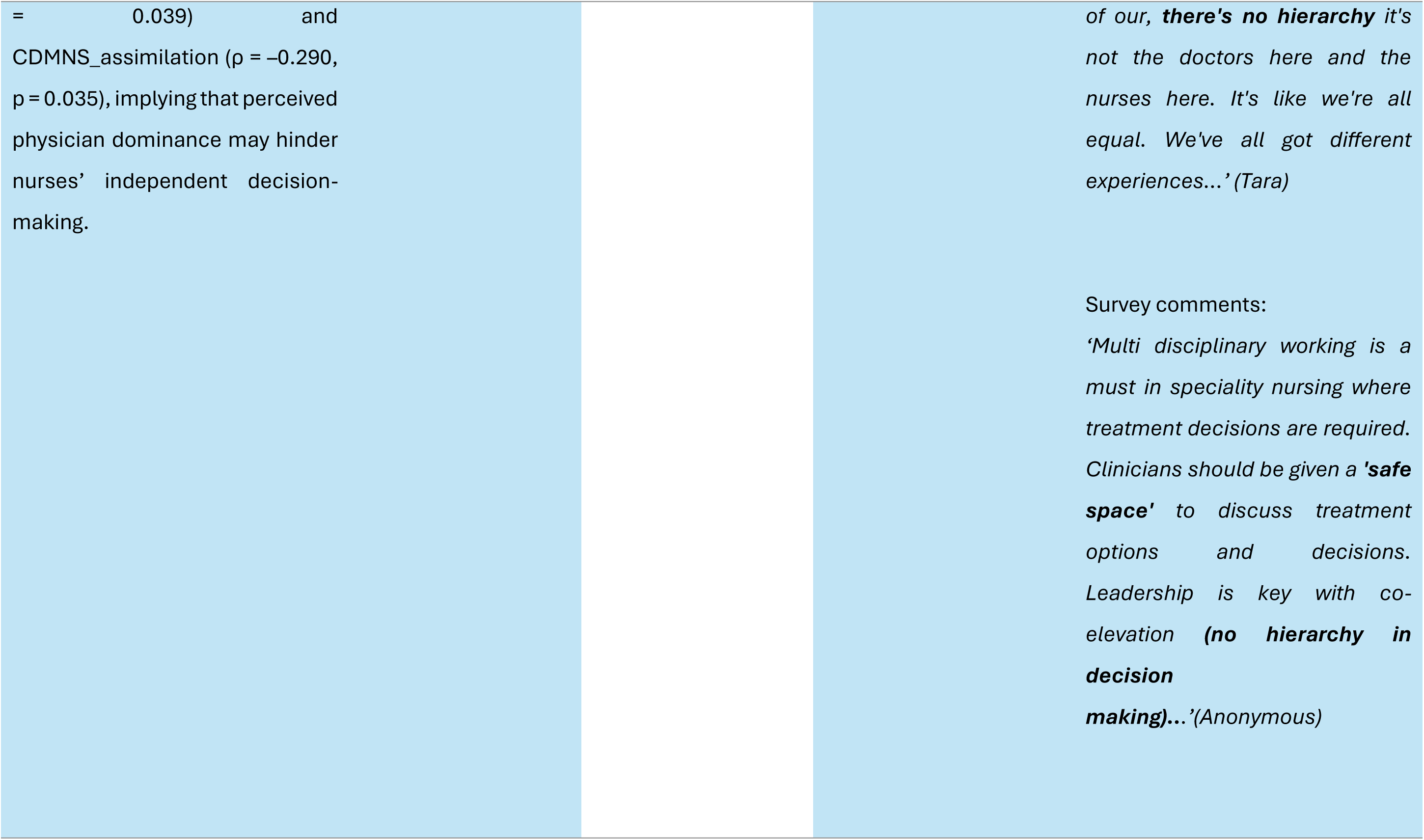
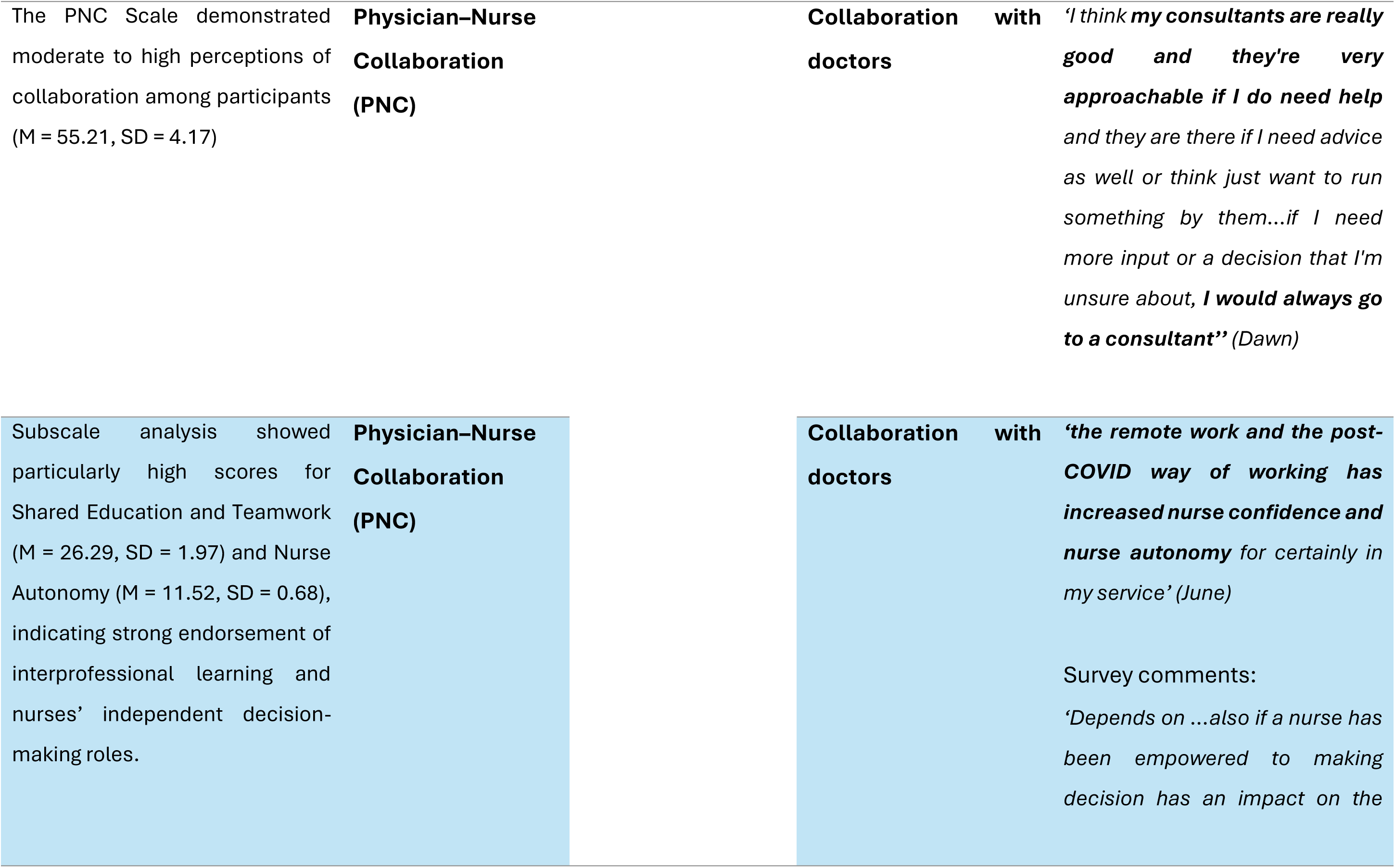
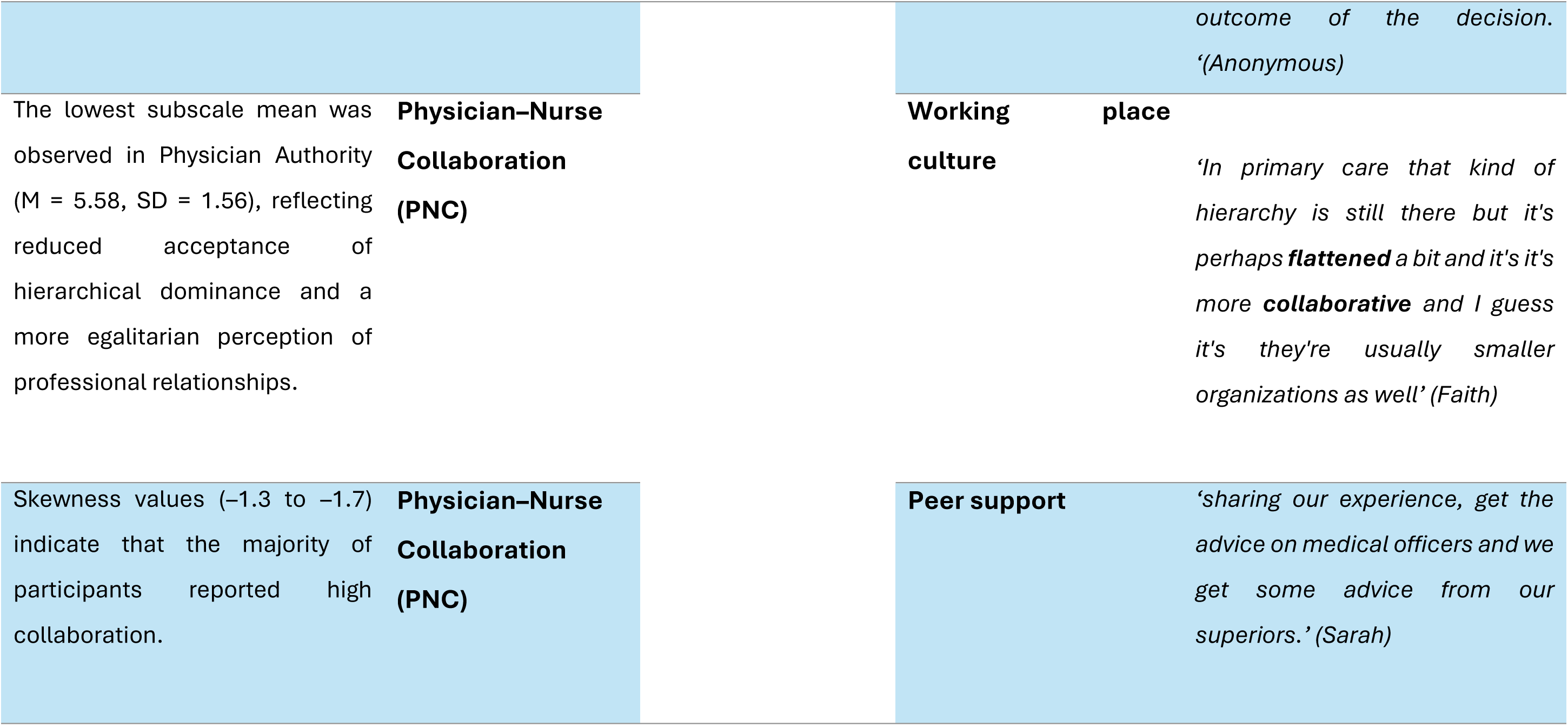

## Appendix 7

### Table Pillar Building for Mixed-method Evidence Synthesis (Pillar Four)

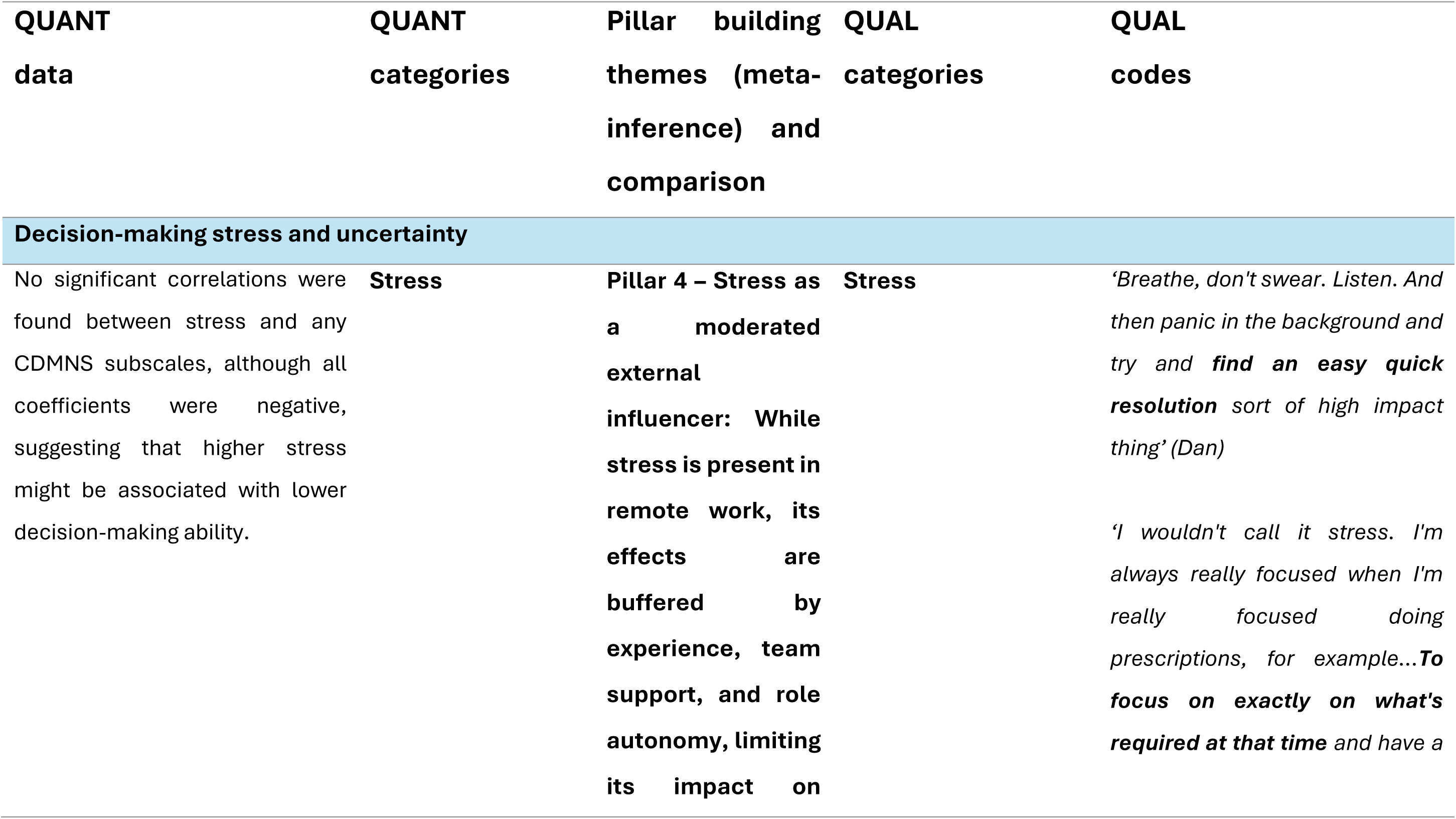
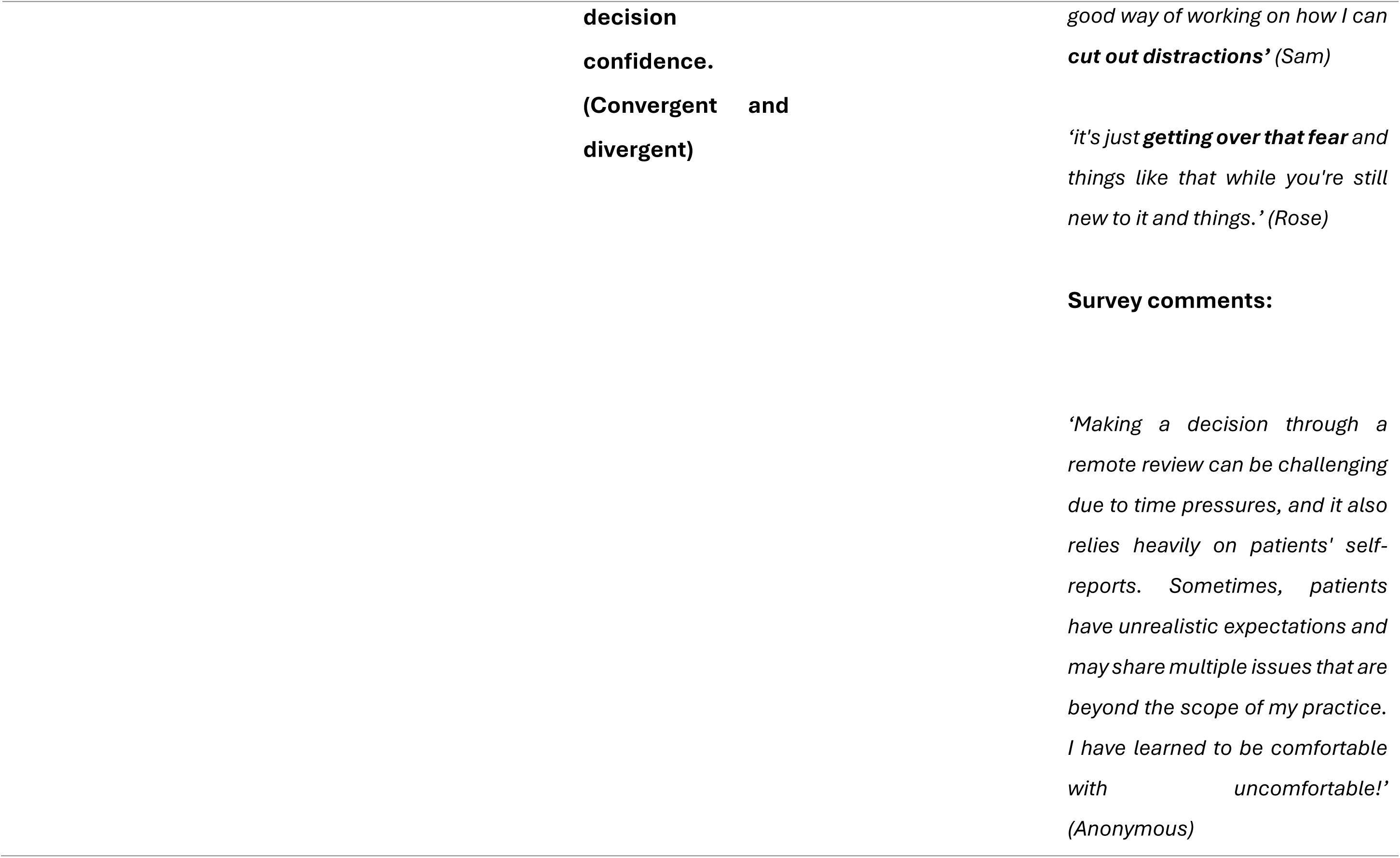
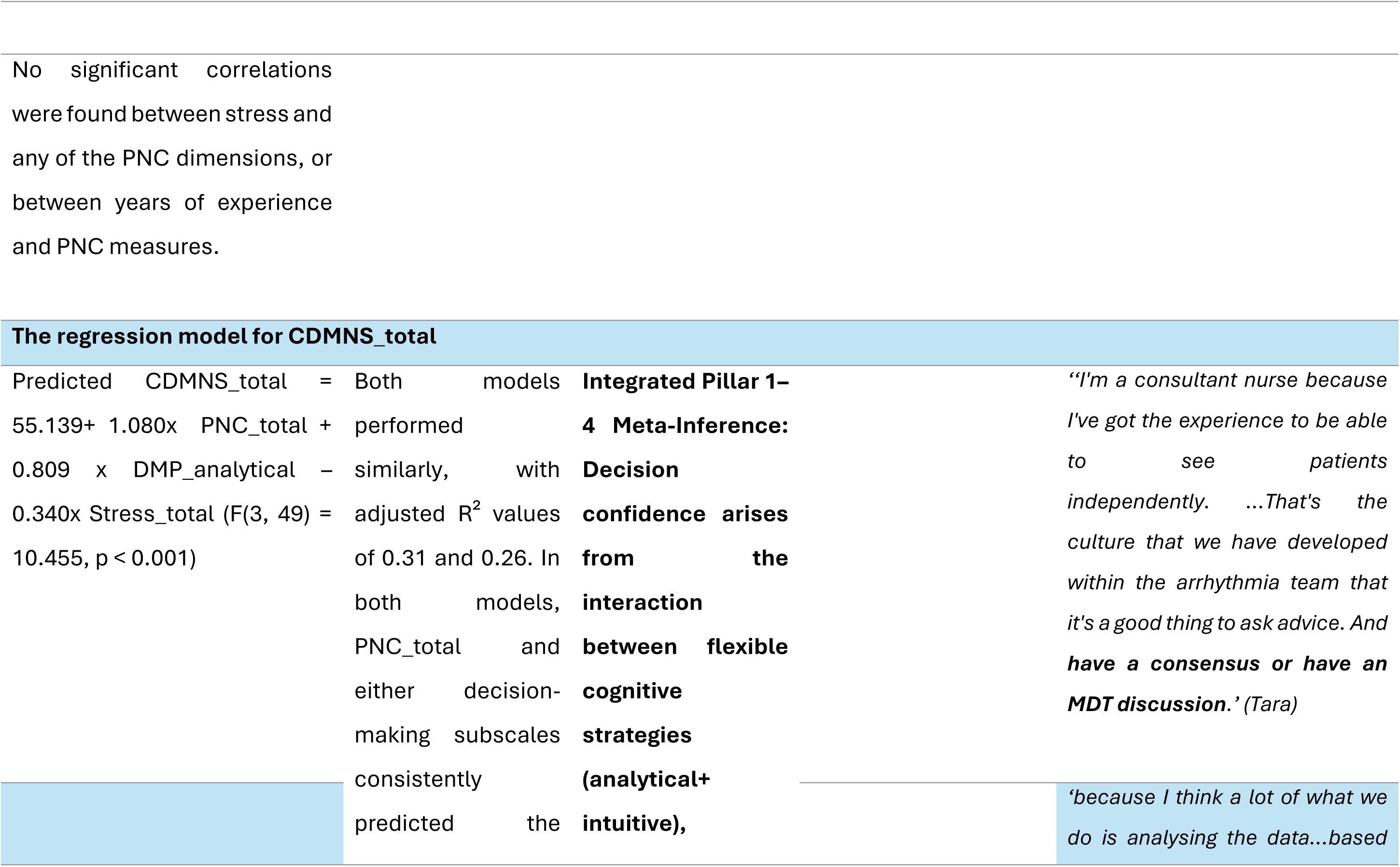
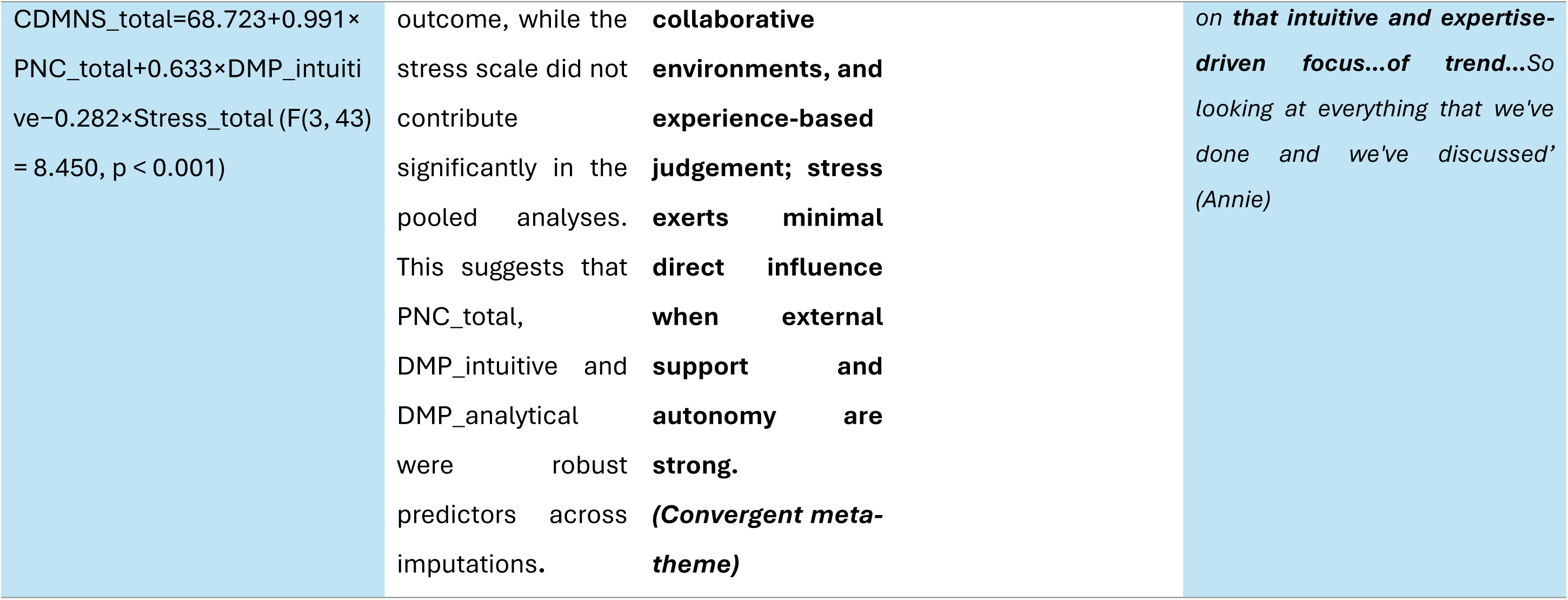

## Appendix 8

### Table Theory-Data Integration Matrix: Linking Empirical Findings to Established Decision-making Theories

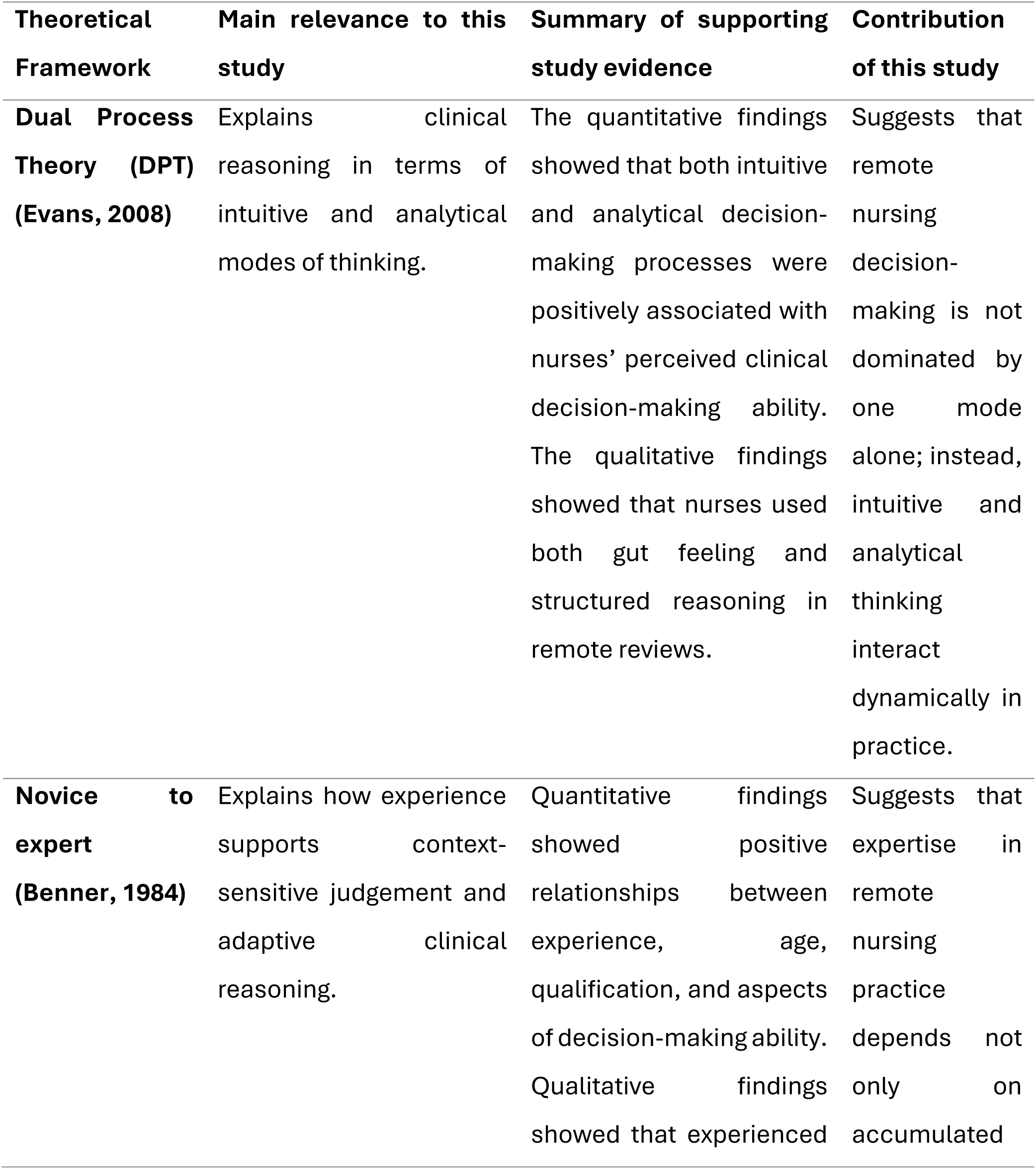
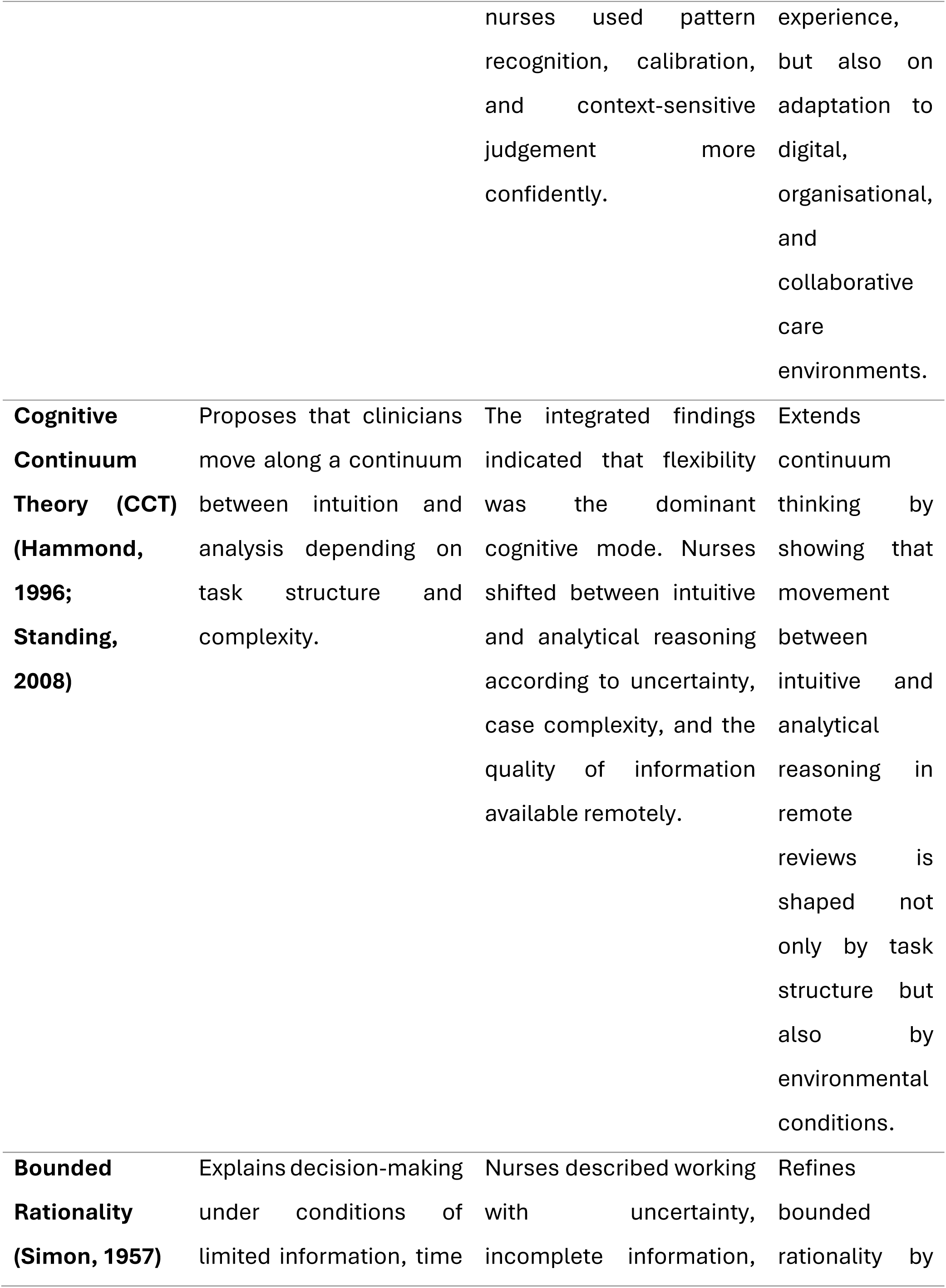
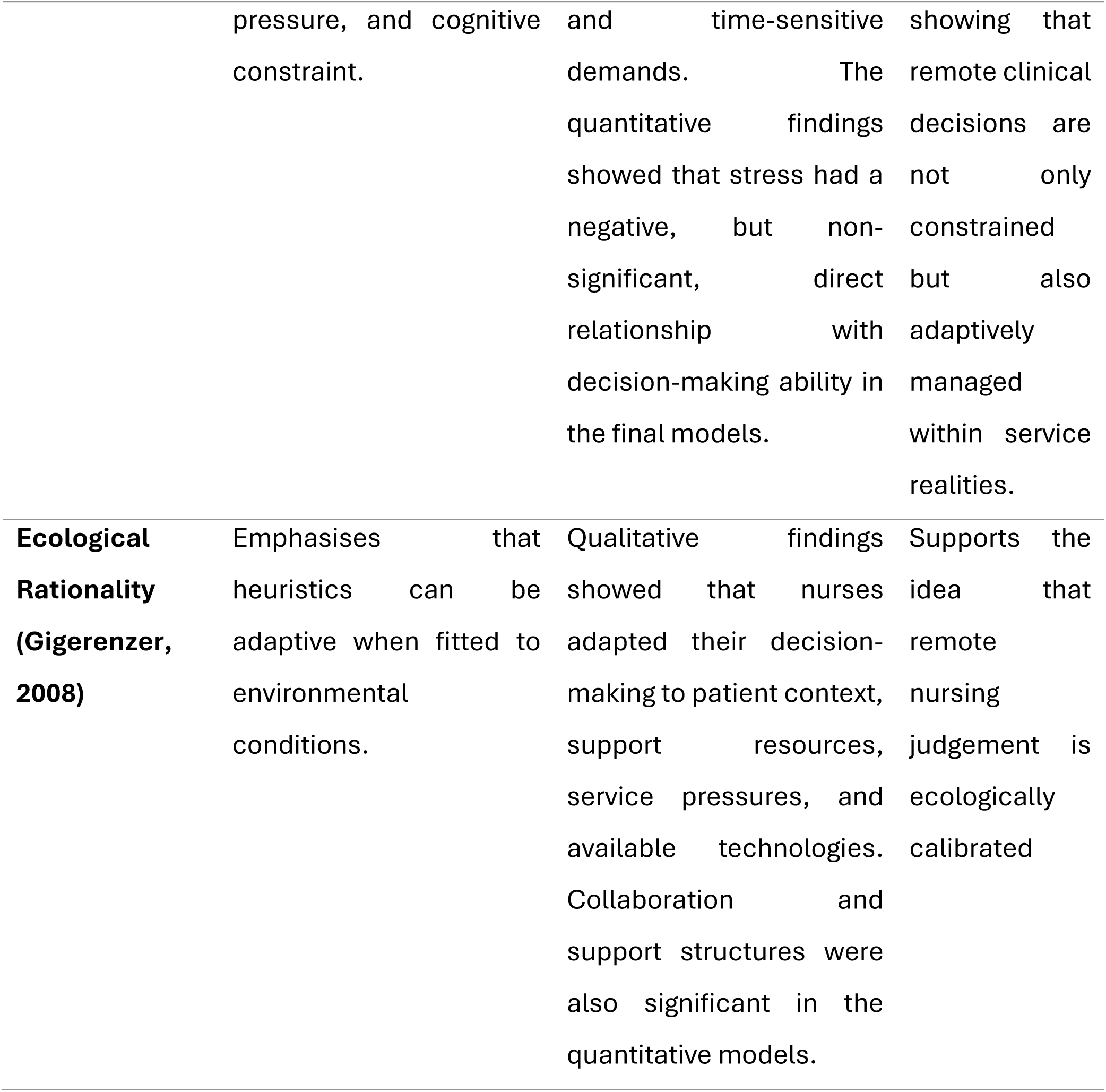

## Appendix 9

### Table Integrated Cognitive-Environment Framework of Remote Clinical Decision-making (ICE)

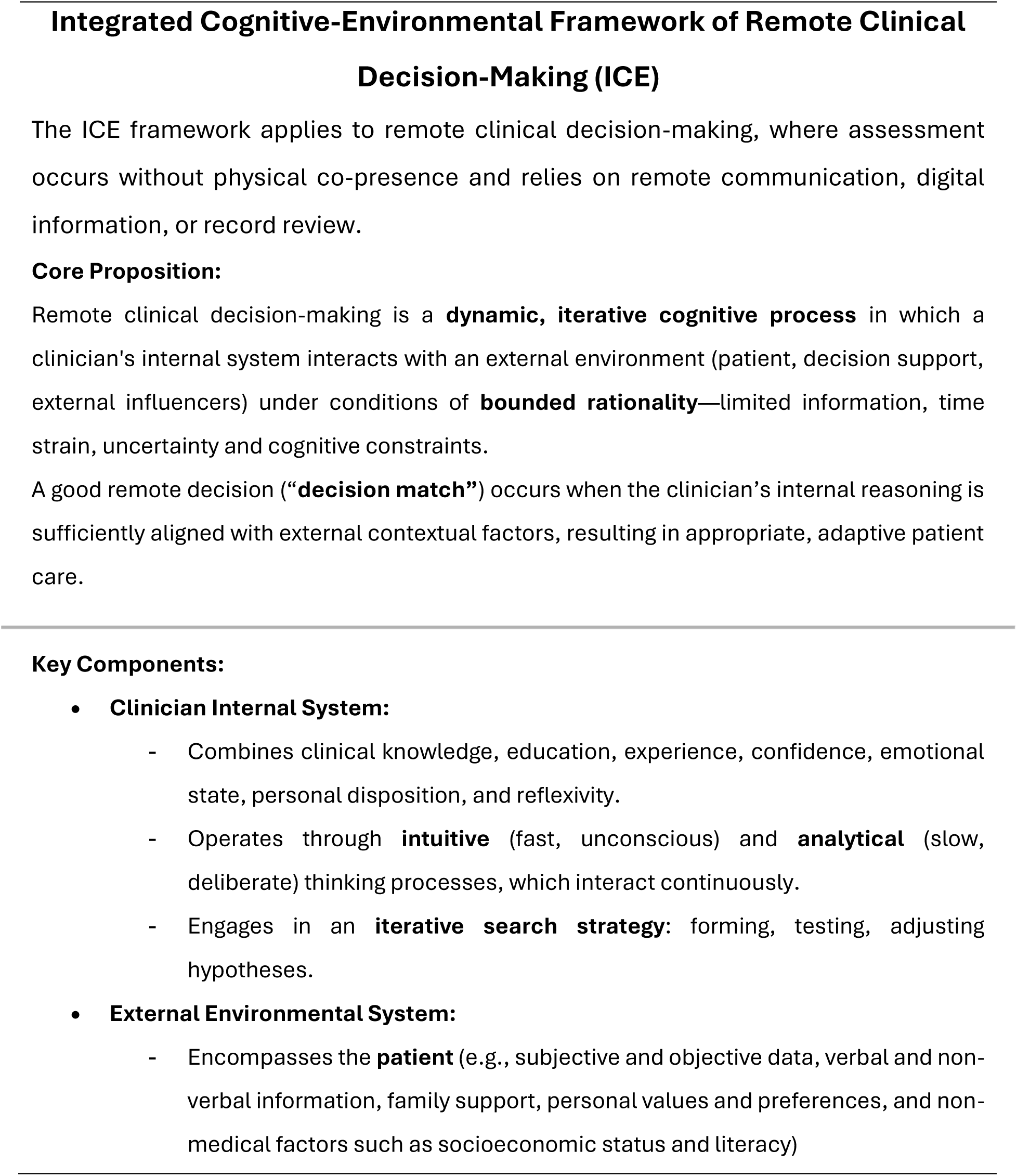
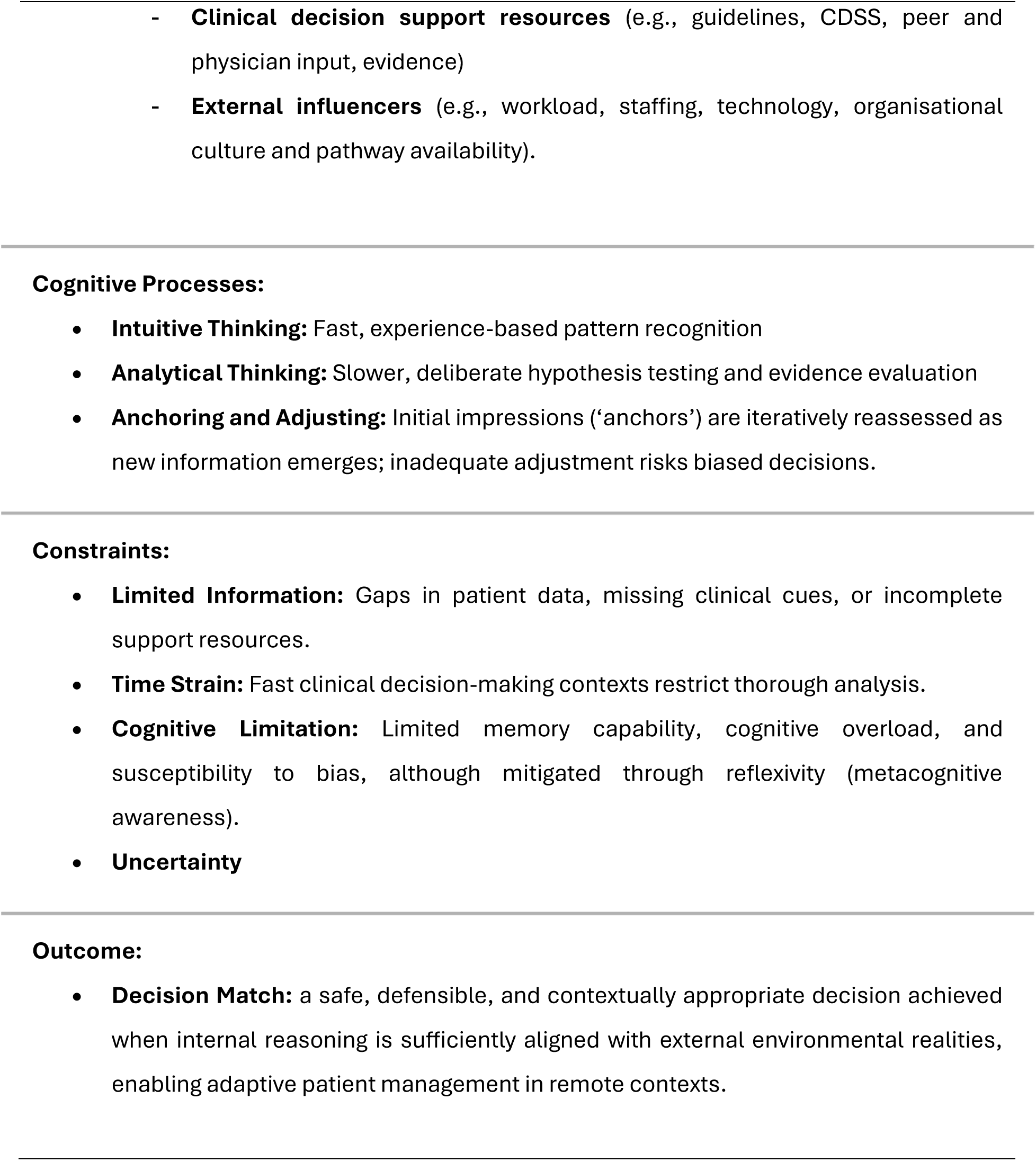

## Appendix 10

### Key Concepts and Definitions in ICE Framework

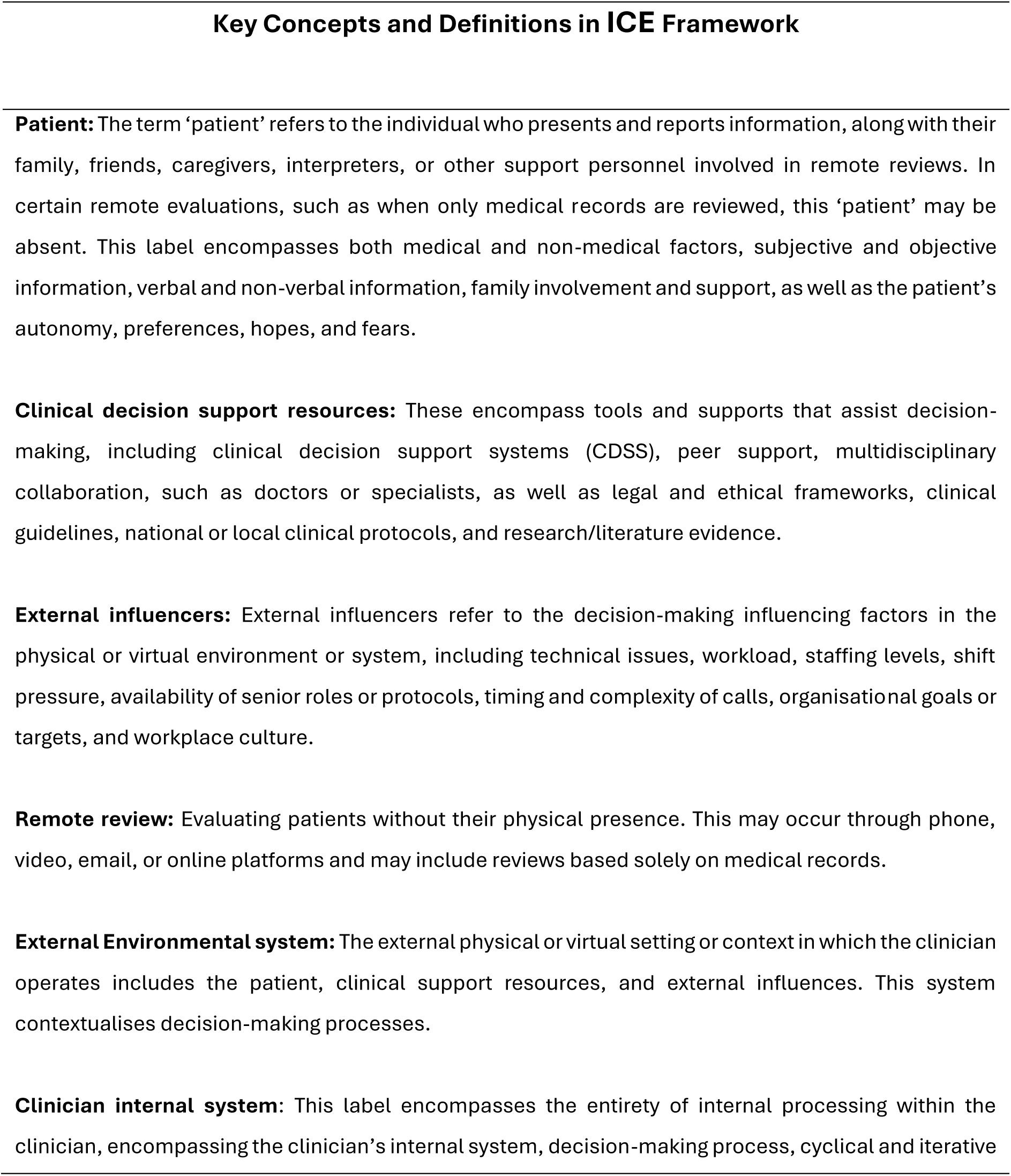
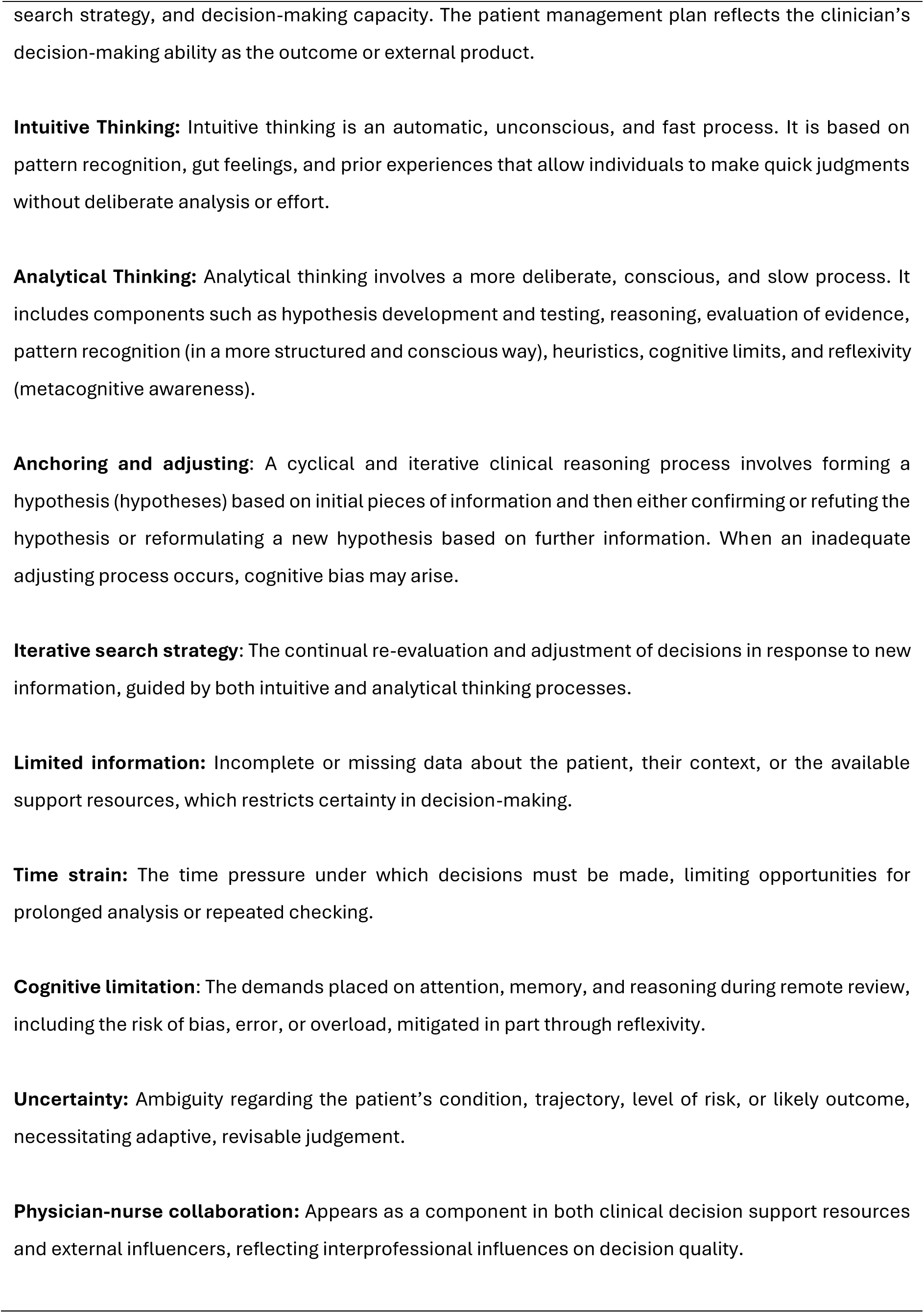
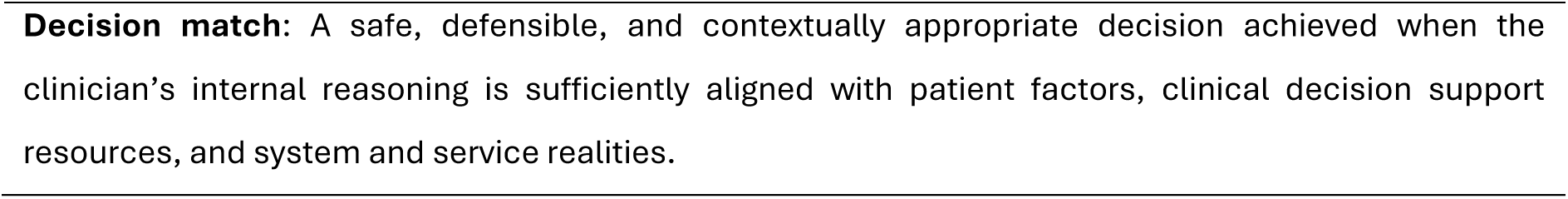

## Appendix 11

### Table Mixed-method evidence supporting the ICE framework

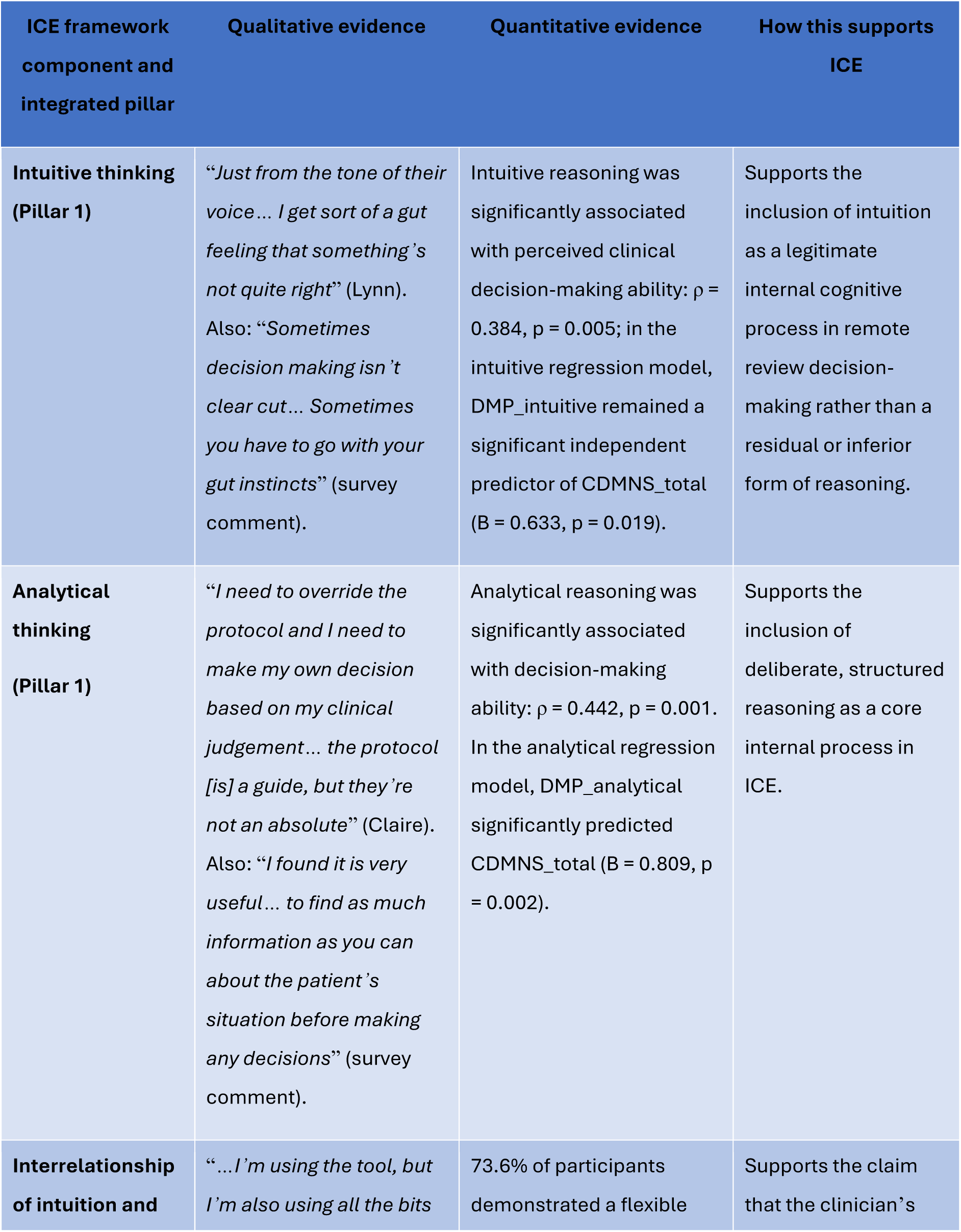
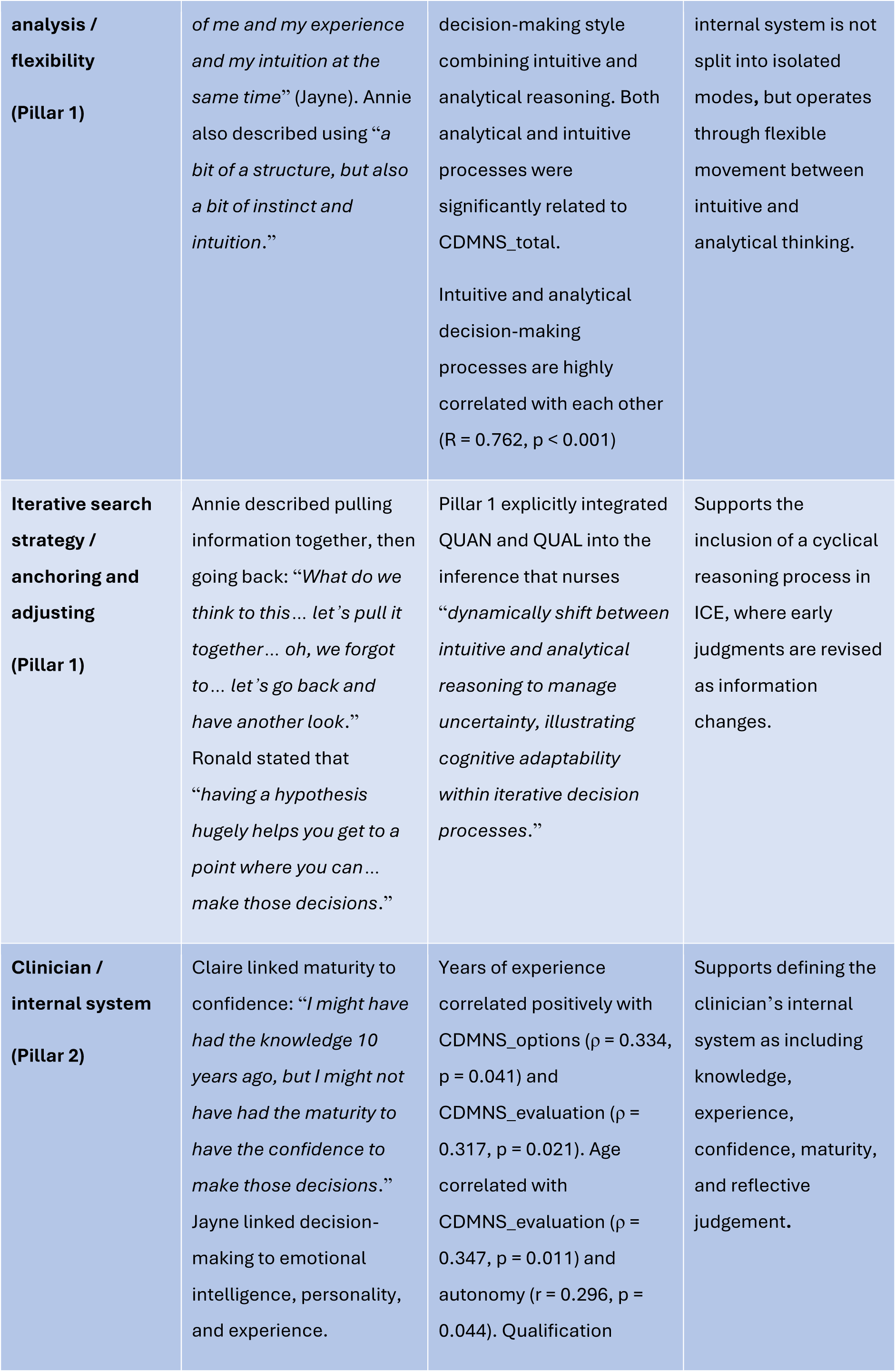
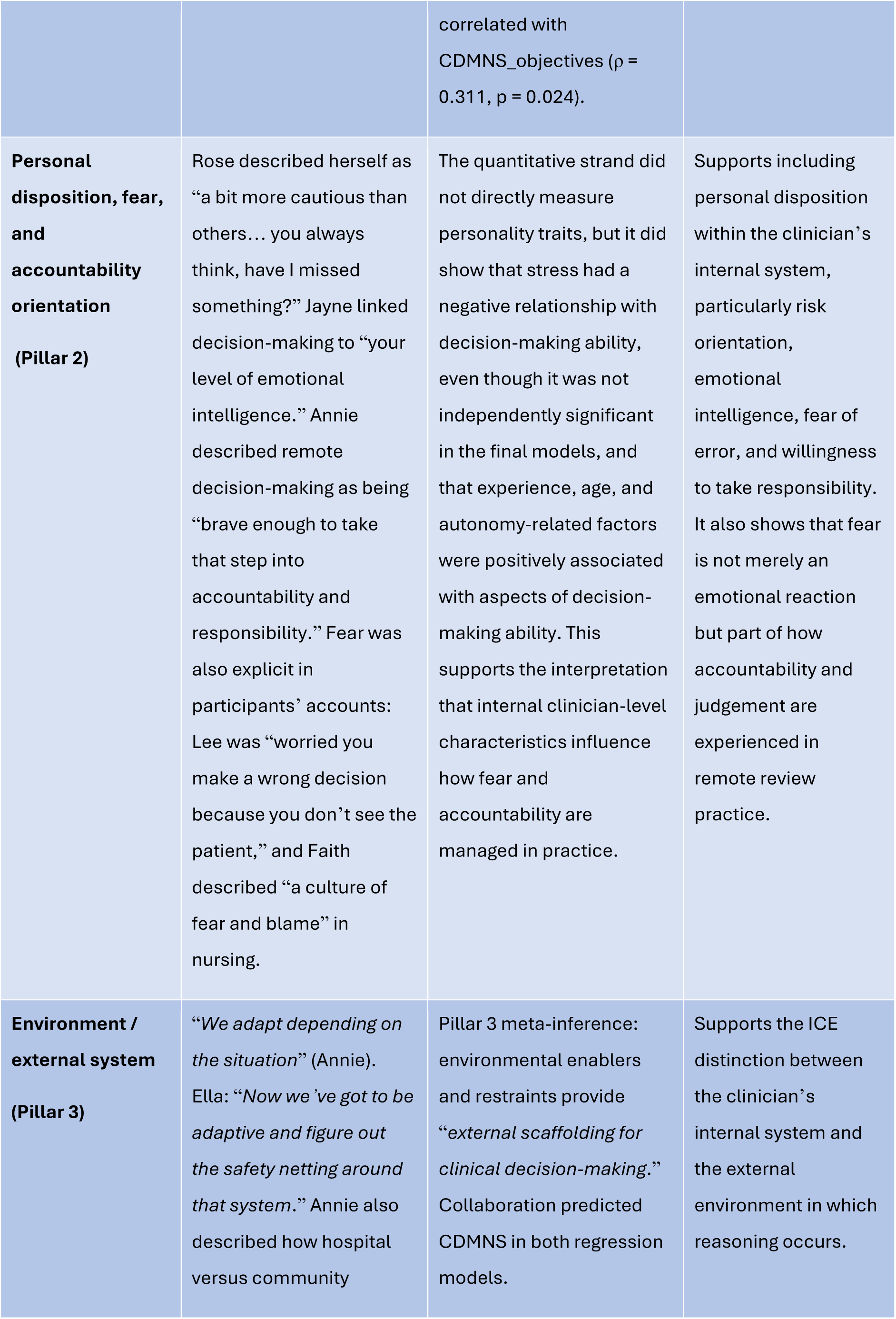
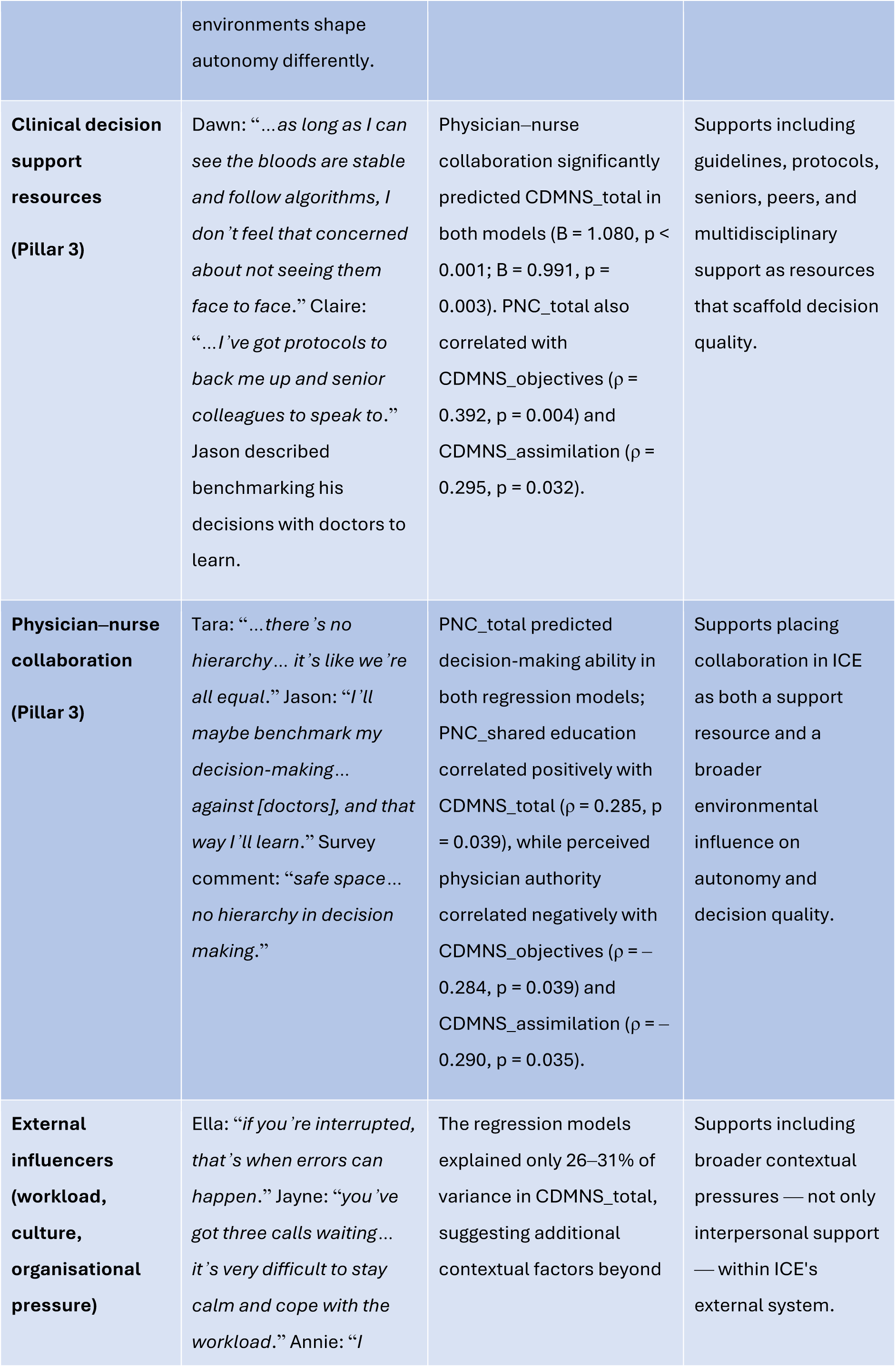
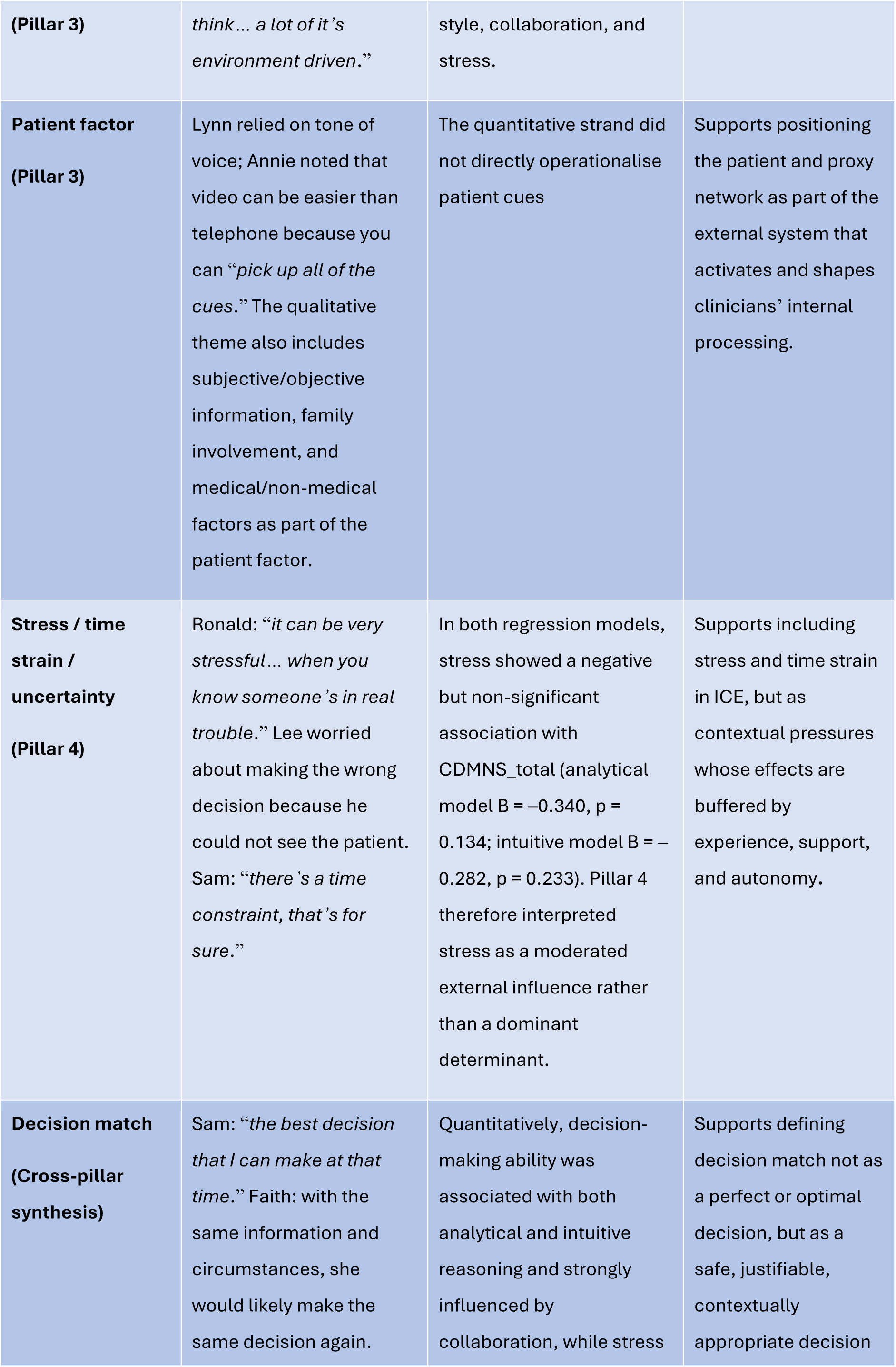
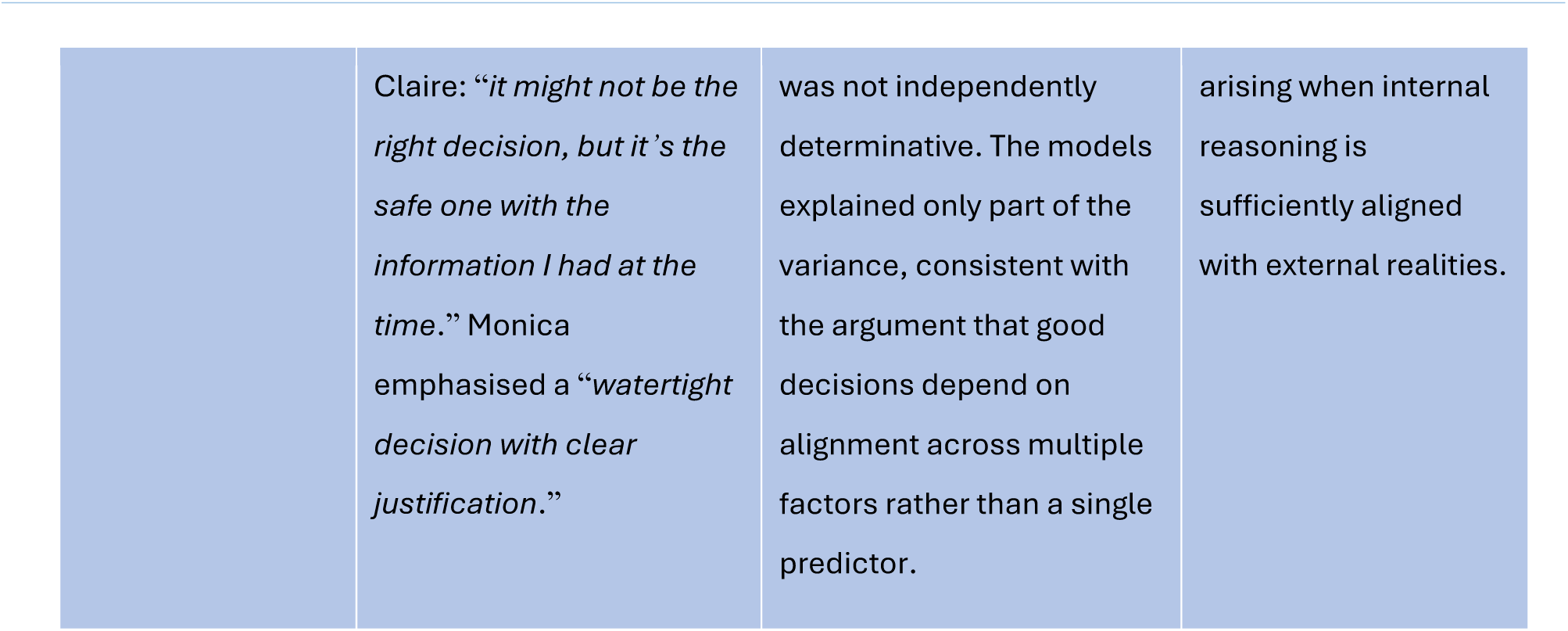

### Mixed Methods Reporting in Rehabilitation & Health Sciences (MMR-RHS)

| Section | Checklist item | Y/N | Page(s) | Comments |
| --- | --- | --- | --- | --- |
| Title | Concise describes the topic of the study identifying the study as mixed methods | Y | 1 | Title includes the topic and explicitly states 'Convergent Mixed-methods Study'. |
| Abstract | Summarizes key elements using journal-specific abstract format | Y | 2 | Abstract includes background, aim, methods, results, conclusion, and impact. |
| Introduction | Includes literature review on the topic of interest (quantitative, qualitative, and mixed) | Y | 3–4 | Background literature and theoretical literature are summarised; mixed-methods reporting standard is also identified. |
| Introduction | Identifies gap that justifies the need for mixed methods approach | Y | 3–4 | Gap is stated as limited empirical evidence on how nurses make remote clinical decisions and what shapes that process. |
| Introduction | Clearly states overarching goal of the study that supports a mixed methods approach | Y | 4–5 | Overall aim and objectives cover both measurable associations and lived experiences/perceptions. |
| Introduction | States the rationale for using mixed methods research | Y | 4–5 | Rationale is implicit in the need to examine both relationships between variables and nurses’ experiences |
| Introduction | Clearly identifies discrete aim(s) for qualitative and quantitative components | Y | 4–5 | Objectives distinguish decision-making processes/influencers/associations (quantitative) and experiences/perceptions (qualitative). |
| Introduction | Aims align with corresponding component methods | Y | 8–15 | Quantitative survey and qualitative interviews map onto the stated objectives. |
| Introduction | Provides statement of significance and potential impact | Y | 2, 4, 46–49 | Significance appears in the introduction, abstract impact statement, and implications section. |
| Methods – Design | Clearly describes the mixed methods design used to accomplish the overarching goal | Y | 8 | Convergent mixed-methods design is named and described. |
| Methods – Design | Emphasis noted (sequential or concurrent QUAL/QUAN) | Y | 8 | Convergent emphasis is stated. |
| Methods – Design | Visual display of overall design highlighting integration | Y | 8 | Figure 3 presents the study flow chart. |
| Methods | Describes and supports the qualitative and quantitative methodologies used to accomplish the discrete aims | Y | 8, 11–14 | Descriptive cross-sectional quantitative component and generic qualitative method are described. |
| Methods | States researcher(s) background and contributions to project (e.g. content or methods expertise, relationships to participants) | Y | 9 | Researcher relationship is addressed through exclusion of the researcher's OUH virtual ward colleagues from interviews. |
| Methods | Identifies setting (e.g. hospital system, geographical location) | Y | 9 | Three hospitals in three UK regions; recruitment also through Facebook, X, and LinkedIn. |
| Methods – Subjects/Participants | Sampling and recruitment | Y | 9–10 | Sampling, sampling technique, and recruitment processes are clearly described. |
| Methods – Subjects/Participants | Inclusion/Exclusion criteria | Y | 9 | Inclusion and exclusion criteria are listed. |
| Methods – Subjects/Participants | Ethical considerations (consent process, researcher relationship with participants) | Y | 10, 15 | Electronic consent, interview consent, ethics approvals, and colleague exclusion are described. |
| Methods – Data collection | Pilot study (if applicable) | N/A |  |  |
| Methods – Data collection | Instrumentation (validity, reliability) | Y | 11–13 | Measures are described with sources and reliability statistics. |
| Methods – Data collection | Implementation matrix (e.g. data source, timeline, type, anticipated outcomes) | N/A |  |  |
| Methods – Data analysis | Qualitative analysis procedures | Y | 14 | Framework analysis steps are clearly described. |
| Methods – Data analysis | Quantitative analysis procedures | Y | 13–14 | Descriptive statistics, group comparisons, correlations, regression, and imputation are described. |
| Methods – Data analysis | Mixed methods analysis / integration procedures | Y | 14 | Pillar-building and theory-building integration are described. |
| Methods – Methodological rigor | Qualitative rigor / trustworthiness | Y | 15 | Credibility, dependability, and confirmability are referenced. |
| Methods – Methodological rigor | Quantitative rigor | Y | 13, 15 | Validated instruments, standardised data collection, internal consistency, and transparent statistics are reported. |
| Methods – Methodological rigor | Mixed methods validity / legitimacy | Y | 14–15 | Integration procedures are described |
| Results / Findings | Clearly presents qualitative findings (includes data exemplars) | Y | 24–35 | Themes, subthemes, and verbatim participant quotations are presented. |
| Results / Findings | Clearly presents quantitative findings | Y | 16–24 | Demographics, descriptive statistics, associations, and regression models are reported. |
| Results / Findings | Provides integrated findings / overall study results | Y | 35–43 | Integrated pillars and theory-driven interpretation are presented. |
| Results / Findings | Provides joint display | Y | 14, 35–43 | Integration through joint display tables is described; appendices are referenced for further detail. |
| Discussion | Incorporates discussion on implications of integrated findings | Y | 44-48 | Discussion and implications sections interpret the integrated findings. |
| Discussion | Provides synthesis and interpretation in context of existing literature and theoretical / conceptual framework | Y | 44-48 | Findings are discussed in relation to bounded rationality, ecological rationality, DPT, CCT, and prior literature. |
| Discussion | Includes subsection of limitations | Y | 48 | Limitations are explicitly headed and discussed. |

© 2023 Tovin M. & Wormley M.

